# A phase II multi-center, open-label, randomized, parallel-group, superiority study to compare the acute toxicity of Hypofractionated dOse Painted External radiotherapy versus conventional external radiotherapy in patients with carcinoma cervix stage I - IIB. (HOPE)

**DOI:** 10.64898/2026.07.24.26358844

**Authors:** Santam Chakraborty, Tapesh Bhattacharyya, Bhavana Rai, Srinivasa GY, Shirley Lewis, Ankita Mallick, Pritanjali Singh, Ram Madhavan, Geetha Muttath, Tejshri, Telkhade, Yusuf Malik, Rajendra Prasad Patel, Sougata Maity, HOPE Trialists Group

## Abstract

**Introduction:** The HOPE trial is a Phase II, multi-center, open-label, randomized, parallel-group superiority study comparing acute toxicity of hypofractionated dose-painted external radiotherapy versus conventional external radiotherapy in patients with carcinoma cervix stages I-IIB (FIGO 2018). The trial will be conducted in India with enrollment commencing July 2026.

**Methods and analysis:** A total of 300 patients will be randomized in a 1:1 ratio to experimental or control arms. The experimental arm employs dose-painted interval-compressed concurrent chemoradiation utilizing volumetric modulated arc therapy (VMAT), delivering 42.5 Gy in 20 fractions over 4 weeks to the primary clinical target volume and 36 Gy in 20 fractions over 4 weeks to elective pelvic nodal volumes (external iliac, internal iliac, presacral, and common iliac). Concurrent chemotherapy consists of weekly cisplatin 50 mg/m². The control arm delivers conventional concurrent chemoradiation with VMAT at 45 Gy in 25 fractions over 5 weeks to both primary and elective volumes, with weekly cisplatin 40 mg/m². Both arms receive image-guided brachytherapy to achieve a total equivalent dose of at least 80 Gy₁₀ to the high-risk clinical target volume.

The primary endpoint is the proportion of patients experiencing grade ≥2 gastrointestinal toxicity (diarrhea, vomiting, nausea) per CTCAE 6.0 criteria. Secondary endpoints include three-year disease-free survival, quality of life, and late toxicity.

**Ethics and dissemination:** The study has been approved by the institutional review board of Tata Medical center. Additional centers will have independent IRB approval after adequate extramural funding has been secured. The results will be disseminated in peer reviewed publications in journals and presented in academic meetings. The study will be conducted as per the applicable laws and regulations of the Indian regulator authorities.

**Registration details:** Indian Clinical Trial Registry (CTRI) CTRI/2026/05/111292

**Strengths and Limitations:**

- First trial of its kind evaluating simultaneous dose de-escalation with hypofractionation with adequate sample size to evaluate impact of toxicity reduction due to bowel and bone marrow dose reduction
- Cumulative dose of cisplatin as well as therapeutic dose fractionation (1.8 Gy) maintained - pure dose de-escalation being evaluated.
- Rigorous multi-step multi-center quality assurance planned
- Limited to accrual in Indian patients.
- Requirement of Intensity Modulated radiotherapy for external beam radiotherapy delivery.

## Introduction

Cervical carcinoma is a major public health concern and is the second most common cancer affecting women in India, with an estimated annual incidence rate of 11.4 per 100,000 [1]. For patients with locally advanced disease, the standard treatment is external-beam radiotherapy (EBRT) to a dose of 40-50 Gy in 1.8-2 Gy fractions over 23-25 fractions, along with concurrent chemotherapy (CCRT) with Cisplatin (40 mg/m2 weekly for 5 cycles) [2,3]. This is followed by intracavitary brachytherapy with a targeted cumulative high-risk clinical target volume (HRCTV) dose of 80-90 Gy [4].

Traditionally, cervical cancer is considered to have an alpha:beta ratio (signifying sensitivity to fraction size) values between 10 - 12 and these have been traditionally used for radiobiological dose calculations. However a recent review compiled the results of alpha:beta ratio values for 98 cervical cancer cell lines from 37 papers [5]. They found that the published results were best fitted by a log-normal distribution with the most probable value of 4.3 Gy. The average value for cervical cancer was 8.05 Gy. Both of these values indicate a higher fraction size sensitivity, which may indicate a role for evaluation of hypofractionated radiotherapy.

Two randomized trials have evaluated hypofractionated radiotherapy with modern radiation techniques. Safaei et al reported a randomized controlled trial (n = 59) comparing the efficacy of hypofractionated radiotherapy to conventionally fractionated radiotherapy (45 Gy / 25 fractions / 5 weeks) [6]. In the hypofractionated radiotherapy arm, a dose of 40 Gy in 15 fractions (three weeks) with weekly Cisplatin to a dose 40 mg/m^2^ (three cycles) was delivered. While the trial demonstrated a similar response in the two arms at three months, grade 3 acute toxicity was higher in patients receiving hypofractionated radiotherapy. Dankulchai et al. reported the results of the HYPOCx-iRex Trial (n = 41) which compared a hypofractionated radiotherapy regimen of 44 Gy in 20 fractions (4 weeks) to a conventional regimen (45 Gy / 25 fractions / 5 weeks) [7]. Concurrent chemotherapy was administered at a dose of 40 mg/m^2^ for 5 cycles in both arms. Grade 3 acute toxicity (CTCAE) was higher in the hypofractionated radiotherapy (43%) as compared to the conventional fractionation (32%). Tumor control was similar in both arms though para-aortic nodal control was improved in the hypofractionated radiotherapy arm.

The results of these two trials demonstrate that whilst the outcomes were comparable, acute toxicity was substantially increased. Additionally, in both trials, the dose of concurrent chemotherapy was compromised. In the trial by Safaei et al, only three cycles were delivered (reducing the total cumulative dose) whilst in the HYPOCx-iRex trial one cycle was administered after the completion of external beam radiotherapy (again reducing the cumulative dose delivered with external beam radiotherapy). Thus there is a need for an alternative method for delivering hypofractionated radiotherapy which does not result in increased toxicity whilst ensuring that cumulative dose of concurrent chemotherapy is not compromised.

The incidence of pelvic nodal failure (PNF) in patients with pelvic node-negative disease at presentation is low. In a study from Japan by Kobayashi et al., the pelvic nodal relapse rate after treatment in patients without pelvic lymph nodes at diagnosis (n = 89) was 1 recurrence in the pelvic node at the first relapse [8]. Of 48 patients with pelvic node-positive disease, 6 developed pelvic nodal relapse. However, in this cohort, only 12 patients received EBRT boost to the pelvic lymph nodes. Among the patients with pelvic node-positive disease, all patients had recurrence in an initially involved node [8]. In a report of pelvic nodal relapse from the EMBRACE I study, among the 641 patients with initially pelvic node-negative disease, a total of 23 patients (3.58%) had pelvic nodal relapse [9]. Among the patients with initial pelvic or para-aortic node-positive disease (n = 697), pelvic nodal relapse was observed in 61 patients (8.75%). Isolated pelvic nodal failure in the absence of other sites of failure was observed in 2% of the patients with pelvic node-negative disease and 3% of patients with node-positive disease [9]. The authors note that the upper border of the pelvic radiotherapy field was not strictly defined, implying that the common iliac chain may not have been adequately covered. In the cohort study conducted at Tata Medical Center, among patients staged with FDG PET/CT, the actuarial cumulative incidence of pelvic nodal failure was 3% at 5 years [10]. We have also audited the outcomes of patients with cervical carcinoma treated between 2019 and 2022 at Tata Medical Center (unpublished data). In this audit, a total of 97 patients with stage I-II squamous cell carcinomas were identified. Among these patients, one pelvic failure (cumulative incidence : 1.1%) was identified at a median follow-up of 45 months.

Given the low incidence of pelvic nodal relapse in patients with node negative cervical cancer, deescalation of dose to elective nodal regions can potentially reduce toxicities. This is particularly true as unlike other Human Papilloma Virus (HPV) associated cancer (anal and head neck cancer) elective dose deescalation has not been evaluated in cervical cancer. Two randomized trials enrolling a total of 500 head neck cancer patients have demonstrated a reduced dose to the elective nodal regions did not result in significant increase in recurrence [11,12]. The equivalent dose at 2 Gy (EQD2) was around 40 - 40.5 Gy_10_ in these trials. It is noteworthy that these results were obtained despite patients having node positive disease and not all patients were HPV positive. Recently, Tsai et al reported on the outcomes of a cohort of HPV+ve oropharyngeal cancer (n = 276) where an elective nodal EBRT dose of 30 Gy (in 15 fractions, EQD2 = 30 Gy) was used. At a median follow-up of 26 months, 8 patients experienced locoregional recurrence, all at the site of primary or nodal gross disease [13]. This translated into a 2-year locoregional control of 97%. In case of anal canal cancer, Lepinoy et al presented outcomes of 142 patients with anal canal cancer treated with an elective inguinal and pelvic nodal radiotherapy dose of 36 Gy followed by a boost to involved nodes and gross disease [14]. These patients had an inguinal control rate of 98.5% and a nodal control of 96%.

This study will evaluate a dose painting approach incorporating a dose reduction for the elective nodal volume (36 Gy in 20 fractions at 1.8 Gy per fraction) whilst simultaneously delivering a moderately hypofractionated dose to the primary disease (42.5 Gy in 20 fractions at 2.12 Gy per fraction) to reduce the overall treatment time. The primary dose is chosen to be isoeffective to the standard 45 Gy in 25 fraction In order to ensure that the cumulative dose of 200 mg/m^2^ cisplatin is delivered in both arms, a dose of 50 mg/m^2^ weekly cisplatin will be administered during radiation per cycle.

Additionally, this regimen allows a dose of 1.8 Gy to be delivered to the elective nodal volume. A lower dose per fraction would be required if a five week regimen is adopted which may be detrimental. In essence an experimental arm with dose deescalation over 5 weeks would involve deescalation of both the total dose and and the dose per fraction. The current regimen is a deescalation only for the total dose and not the dose per fraction. It is noteworthy that for most squamous cell carcinomas, the dose response curve between the dose per fraction region of 1.4 - 1.8 Gy shows a very steep reduction in the response probability below 1.8 [15]. Radiobiological modelling of the reduction in tumor control probability (TCP) due to elective dose de-escalation has been done using the methodology proposed by Moos et al [16] (see full protocol for details). Predicted loss of the TCP with the proposed dose de-escalation exceeds 3 % only when the probability of elective nodal failure is 9% with the standard regimen. However as shown in the literature review this rate is realistically less than 5% and the predicted loss of TCP is 1.74% or less when this is the case.

## Methods and analysis

### Objectives

The primary objective of the study is to compare the proportion of grade 2 or higher acute CTCAE 6.0 defined gastrointestinal (during and upto 6 weeks post treatment) between patients undergoing dose-painted hypofractionated concurrent chemoradiation (HDRT) versus conventional concurrent chemoradiation (CRT). The key gastrointestinal toxicity endpoints will include nausea, vomiting, and diarrhea.

#### Estimand

This outcome (gastrointestinal toxicity) will be calculated for the entire cohort of patients who have been randomized. The comparison will be between the two arms, and in the event of a cross over, the patient will be analyzed as randomized. The risk ratio will be provided along with 95% confidence intervals of the same. In the event of an intercurrent event which results in treatment discontinuation or interruption, the maximum observed grade of the event observed till the observation period will be used. If one of the defined gastrointestinal toxicity leads to the treatment discontinuation or interruption then the grade of the toxicity used will be considered as grade 4. Thus a composite strategy will be used for handling the intercurrent events.

##### Secondary Objectives

1. **Disease Free Survival:** To compare the 3 year disease free survival between patients receiving HDRT vs CRT. The endpoint of disease free survival is defined as the interval of time between randomization to disease recurrence, second metastases or death.
2. **Quality of Life:** To compare the global patient reported quality of life, 15 item summary score, and symptom experience between patients receiving HDRT vs CRT. The key endpoints of interest will include the mean global health related quality of life, 15 item summary score, and the symptom experience score using EORTC QLQ C30 and CX24 questionnaires respectively.
3. **Late Toxicity:** To compare the incidence of any grade late CTCAE 6.0 toxicity between patients undergoing HDRT versus CRT. Late toxicity endpoints of interest will include fatigue, abdominal pain, proctitis, cystitis, vaginal toxicity, bowel toxicity and bone fractures.

The estimands for these objectives are shown in Table 1.

**Table 1:**
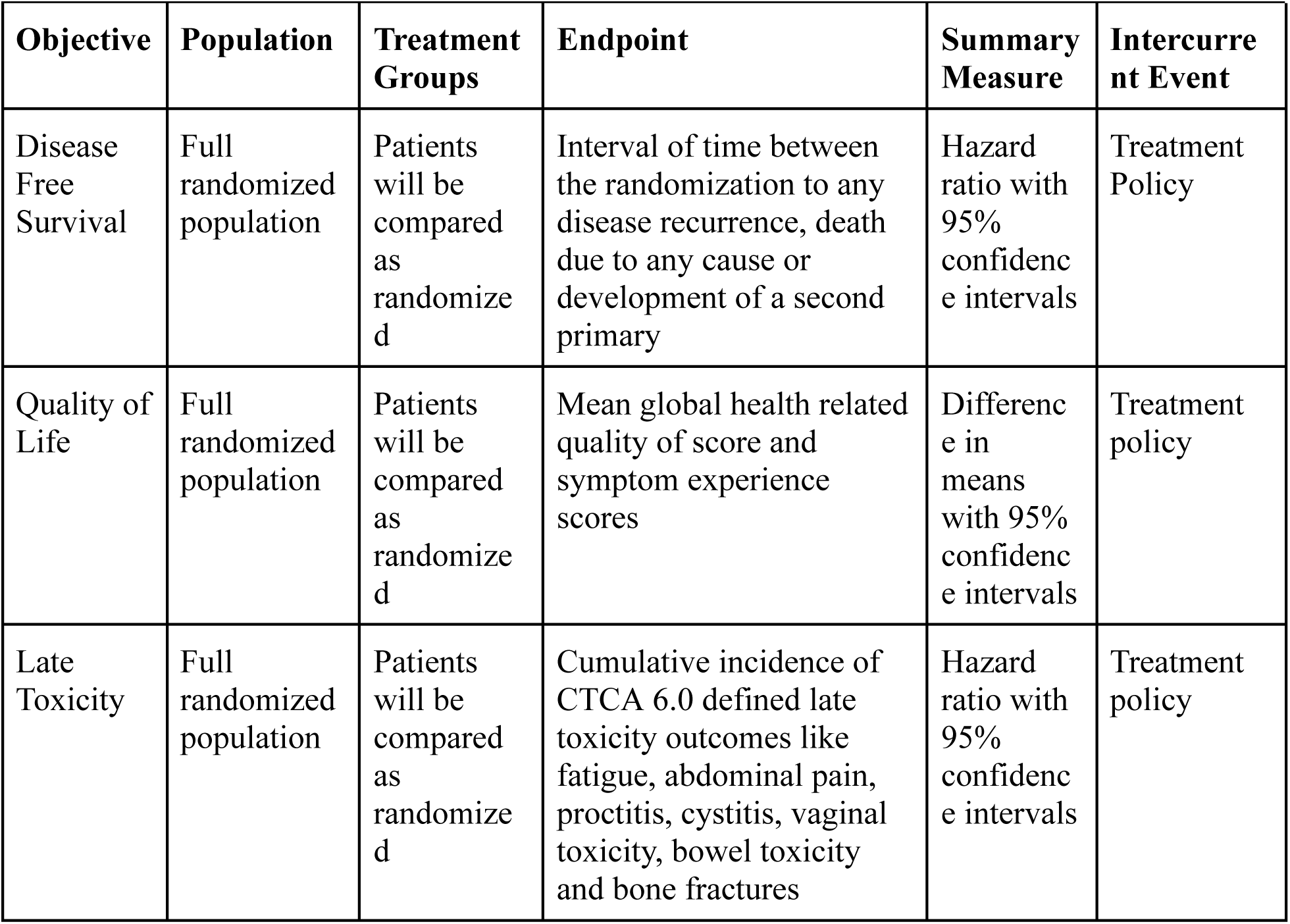
Estimands for secondary objectives. Treatment policy handling of intercurrent events implies that the occurrence of intercurrent events which result in treatment discontinuation or interruptions will not result in exclusion of the patients from the analysis of the endpoint. As late toxicity related events occur after completion of the treatment hence they will be handled as per treatment policy strategy.

###### Trial Run In Objectives

An initial pilot run in phase is planned for this trial where we will evaluate the deliverability of HDRT. The primary objective will be to demonstrate that concurrent chemoradiation is deliverable after chemotherapy. The regimen would be considered deliverable if 16 or more of the 20 planned patients complete the planned treatment without major protocol deviations. If 15 or fewer of these 20 patients do not complete the planned chemotherapy and chemoradiation regimen, then termination of the study will be considered at this time point. Although this part of the study will be a part of the main randomization, evaluation of the protocol adherence and completion will be done only in the HDRT arm.

### Trial Design

The trial design is an open label, parallel group, two arm, randomized controlled trial. An initial trial run-in phase will be included in the design to establish that HDRT can be delivered safely in these patients. Patients enrolled in the run-in phase will be included in the main analysis for toxicity and outcomes.

Stratified randomization will be performed centrally in a 1:1 ratio. Stratification variables will include:

1. Receipt of neoadjuvant chemotherapy following the INTERLACE protocol
2. Primary tumor size : 4 cm or less and more than 4 cm

### Study Setting

The study is designed as a multi-center trial conducted in tertiary cancer centers which are enrolling patients in the ongoing PROPARA trial [17]. Additional trial sites which are not enrolling in the PROPARA trial will also be eligible after radiotherapy trial quality assurance. All trial sites must have access to image guidance in the form of volumetric cone beam imaging and image guided brachytherapy. Use of intensity modulated radiotherapy is mandatory for this trial.

### Eligibility Criteria

#### Inclusion Criteria

All the following criteria must be met to include the patient in the trial.

1. Age between 18 - 70 years
2. Eastern Cooperative Oncology Group (ECOG) Performance status 0 - 2.
3. Biopsy proven squamous cell carcinoma of cervix. HPV status determination using p16 IHC is not mandatory but recommended.
4. FIGO 2018 stage I - IIB disease with staging done using MRI pelvis and FDG PET CT scan. Any visualized lymph node must meet all of the following criteria to be deemed non-pathological:

a. Short axis diameter: < 10 mm
b. Homogenous Texture
c. Smooth Border
d. Any shape with fatty hilum OR bean like / oval shape without fatty hilum.
e. Absence of significant uptake on FDG PET CT (only for patients with no significant contraindication or hypersensitivity which precludes FDG PET/CT)
5. Adequate end organ function to permit delivery of cisplatin based concurrent chemoradiation:

a. Adequate bone marrow function (white blood cells ≥3·0×10⁹/L, platelets ≥100×10⁹/L)
b. Liver function (bilirubin ≤1·5×upper normal limit [UNL], aspartate aminotransferase and alanine aminotransferase ≤2·5×UNL)
c. Kidney function (creatinine clearance >60 mL per min calculated according to Cockroft and Gault or > 50 mL per min DTPA)
6. Women of childbearing age must have a pregnancy test (serum or urine) within 7 days before enrollment. The result is negative, and they are willing to use appropriate contraceptive methods during the test.

#### Exclusion Criteria

None of the criteria noted below must be present.

1. Presence of pathologically proven or radiologically suspected pelvic or para-aortic lymphadenopathy. Radiologically suspicious nodes will include any lymph node with:

a. Short axis diameter ≥ 10 mm
b. Heterogenous Texture
c. Irregular Border
d. Any shape with loss of fatty hilum OR rounded shape without fatty hilum.
e. Presence of significant uptake on FDG PET CT (only for patients with no significant contraindication or hypersensitivity which precludes FDG PET/CT)
2. Presence of significant uterine body involvement defined as involvement beyond the lower uterine segment.
3. Presence of significant uterine fibroids or intrauterine collection that is likely to result in significant portion of bowel receiving hypofractionated radiotherapy
4. Patients who have received prior pelvic radiotherapy
5. Patients with synchronous malignancies in the abdomino-pelvic region which are likely to require radiotherapy
6. Patients with prior history of malignancy excluding cervical intra-epithelial neoplasia or basal cell carcinoma of the skin.
7. Liver cirrhosis, decompensated liver disease, chronic renal insufficiency, and renal failure
8. Presence of uncontrolled or severe comorbidities which preclude safe concurrent chemoradiation e.g Myocardial infarction, severe arrhythmia, and grade 2 or more congestive heart failure (NYHA classification).
9. Prior history of renal transplantation
10. Horseshoe kidney or single kidney
11. History of severe allergic reaction to platinum-containing chemotherapy drugs
12. Patients with a history of autoimmune diseases are known to increase the risk of late radiation induced adverse effects especially Systemic Lupus Erythematosus.
13. Patients with accompanying diseases or other special circumstances that seriously endanger the safety of patients or affect the completion of the study

Please refer to the main protocol for details on the registration, withdrawal criteria, patient transfer and trial center requirements.

### Intervention and Comparator

#### Interventions

1. **Control Arm (CRT) :** Patients will undergo radiotherapy to the whole pelvis (primary and elective clinical target volumes) to a dose of 45 Gy in 25 fractions (1.8 Gy per fraction) delivered over a period of five weeks along with concurrent chemotherapy using Cisplatin at a dose of 40 mg/m^2^ weekly (capped at a maximum of 70 mg per cycle).
2. **Experimental Arm (HDRT) :** Patients will under radiotherapy to a dose of 42.5 Gy in 20 fractions (2.12 Gy per fraction) to the primary clinical target volume and a dose of 36 Gy in 20 fractions (1.8 Gy per fraction) to the elective nodal clinical target volume delivered over a period of 4 weeks. Concurrent cisplatin will be administered at a dose of 50 mg/m^2^ weekly (capped at a maximum of 80 mg per cycle).

After completion of concurrent chemoradiation, patients in both arms will undergo brachytherapy using intracavitary or combined intracavitary-interstitial brachytherapy. The linear quadratic equivalent dose (LQED2) at 2 Gy to the high-risk clinical target volume (HR CTV) D90 should be more than ≥ 80 Gy depending on HRCTV volume. Alpha beta ratio for calculation of LQED2 for the high risk CTV would be 10.

Patients in both arms will undergo image guidance in the form of volumetric imaging using onboard cone-beam or megavoltage fan beam CT scans. Daily image guidance is recommended but at least 3 fractions per week should have volumetric image guidance. Similarly use of image guided brachytherapy is recommended with pre-brachytherapy MRI or MRI done with an applicator in situ. At a minimum CT guidance should be available for brachytherapy planning.

#### External Beam radiotherapy

External beam radiotherapy will be delivered using intensity modulated radiotherapy. The primary CTV (CTVp) will encompass the gross disease and the entire cervix and uterus. The vaginal extent of the disease will determine the length of vagina to be included in the primary CTV. Typically at least 2 cm of the normal vagina beyond the clinically or radiologically demonstrated disease should be included in the CTV. CTV primary should include parametrium which will extend superiorly up to the peritoneal reflection (guided by the presence of the round ligament or the sigmoid colon). Anteriorly the boundary will be the posterior wall of the bladder or the posterior border of the external iliac vessel. Laterally it will extend to the medial edge of the internal iliac vessels. Inferiorly the volume will be bounded by the urogenital diaphragm. The elective nodal CTV (CTVn_Pelvic) will include the common iliac, external iliac, internal iliac, presacral, and obturator group of lymph nodes. The constraints to be followed for treatment planning are shown in Table 2. For further details of the immobilization, pretreatment imaging, target volume delineation, planning, plan quality assurance and handling of treatment interruptions please refer to the full protocol.

**Table 2:**
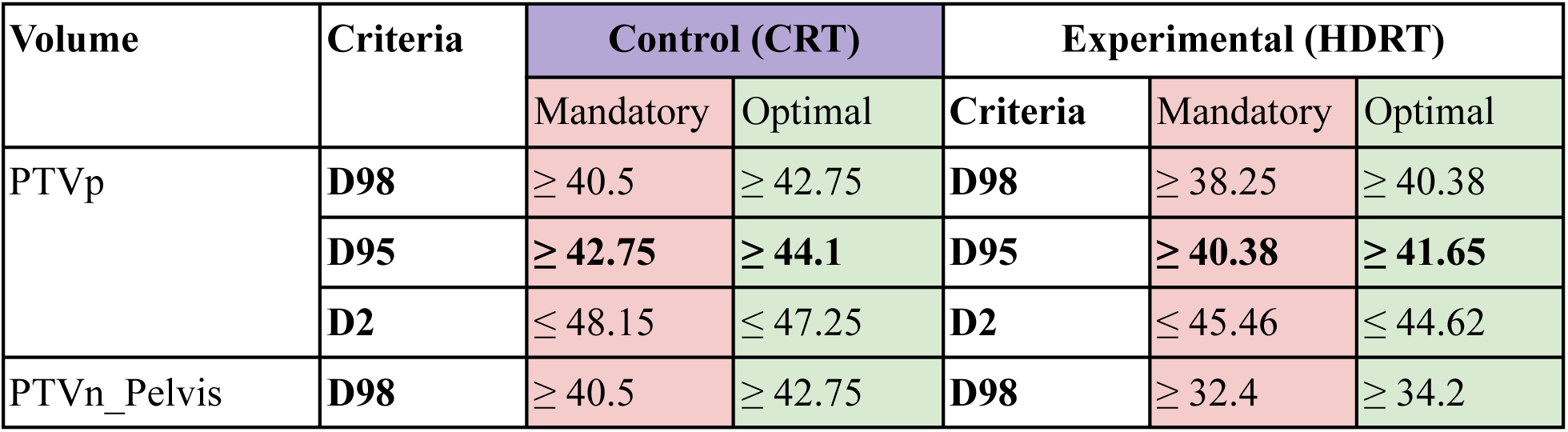
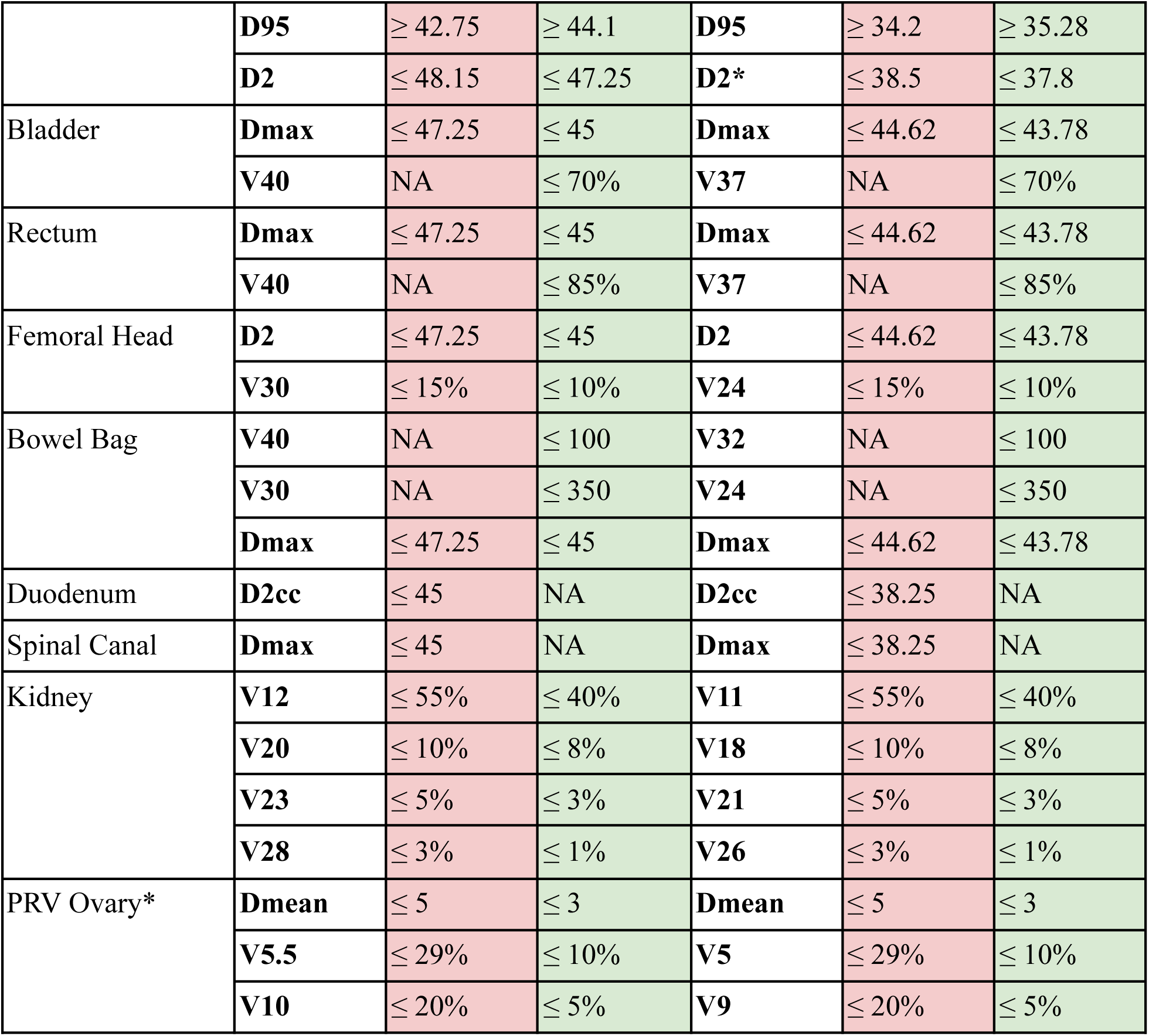
Showing the dose constraints to be followed for the trial. Mandatory constraints should be met in all cases. * D2 for PTVn_Pelvis for the HDRT arm to be evaluated for a planning volume which is not overlapping the PTVp with an additional margin of 3 mm. ** PRV Ovaries are required only if oophoropexy has been performed. Vx =Volume receiving x dose in Gy, Dx = Minimum dose to x volume. For CRT arm plan evaluation for the PTVp and PTVn_Pelvis can be done as a singular structure.

| Volume | Criteria | Control (CRT) |  | Experimental (HDRT) |  |  |
| --- | --- | --- | --- | --- | --- | --- |
|  |  | Mandatory | Optimal | Criteria | Mandatory | Optimal |
| PTVp | **D98** | $\geq 40.5$ | $\geq 42.75$ | **D98** | $\geq 38.25$ | $\geq 40.38$ |
| | **D95** | $\geq 42.75$ | $\geq 44.1$ | **D95** | $\geq 40.38$ | $\geq 41.65$ |
| | **D2** | $\leq 48.15$ | $\leq 47.25$ | **D2** | $\leq 45.46$ | $\leq 44.62$ |
| PTVn_Pelvis | **D98** | $\geq 40.5$ | $\geq 42.75$ | **D98** | $\geq 32.4$ | $\geq 34.2$ |
| | **D95** | $\geq 42.75$ | $\geq 44.1$ | **D95** | $\geq 34.2$ | $\geq 35.28$ |
| | **D2** | $\leq 48.15$ | $\leq 47.25$ | **D2\*** | $\leq 38.5$ | $\leq 37.8$ |
| Bladder | **Dmax** | $\leq 47.25$ | $\leq 45$ | **Dmax** | $\leq 44.62$ | $\leq 43.78$ |
| | **V40** | NA | $\leq 70\%$ | **V37** | NA | $\leq 70\%$ |
| Rectum | **Dmax** | $\leq 47.25$ | $\leq 45$ | **Dmax** | $\leq 44.62$ | $\leq 43.78$ |
| | **V40** | NA | $\leq 85\%$ | **V37** | NA | $\leq 85\%$ |
| Femoral Head | **D2** | $\leq 47.25$ | $\leq 45$ | **D2** | $\leq 44.62$ | $\leq 43.78$ |
| | **V30** | $\leq 15\%$ | $\leq 10\%$ | **V24** | $\leq 15\%$ | $\leq 10\%$ |
| Bowel Bag | **V40** | NA | $\leq 100$ | **V32** | NA | $\leq 100$ |
| | **V30** | NA | $\leq 350$ | **V24** | NA | $\leq 350$ |
| | **Dmax** | $\leq 47.25$ | $\leq 45$ | **Dmax** | $\leq 44.62$ | $\leq 43.78$ |
| Duodenum | **D2cc** | $\leq 45$ | NA | **D2cc** | $\leq 38.25$ | NA |
| Spinal Canal | **Dmax** | $\leq 45$ | NA | **Dmax** | $\leq 38.25$ | NA |
| Kidney | **V12** | $\leq 55\%$ | $\leq 40\%$ | **V11** | $\leq 55\%$ | $\leq 40\%$ |
| | **V20** | $\leq 10\%$ | $\leq 8\%$ | **V18** | $\leq 10\%$ | $\leq 8\%$ |
| | **V23** | $\leq 5\%$ | $\leq 3\%$ | **V21** | $\leq 5\%$ | $\leq 3\%$ |
| | **V28** | $\leq 3\%$ | $\leq 1\%$ | **V26** | $\leq 3\%$ | $\leq 1\%$ |
| PRV Ovary* | **Dmean** | $\leq 5$ | $\leq 3$ | **Dmean** | $\leq 5$ | $\leq 3$ |
| | **V5.5** | $\leq 29\%$ | $\leq 10\%$ | **V5** | $\leq 29\%$ | $\leq 10\%$ |
| | **V10** | $\leq 20\%$ | $\leq 5\%$ | **V9** | $\leq 20\%$ | $\leq 5\%$ |

For details on expected adverse effects, handling of treatment interruptions, treatment modifications and concomitant therapy please refer to the full protocol.

### Outcomes

#### Primary Outcome

The primary outcome of interest is the incidence of acute toxicity defined using the CTCAE 6.0 grading system. Acute toxicity is defined as an adverse effect occurring during the course of radiotherapy and upto 6 weeks after completion of treatment. The maximum grade of each toxicity endpoint the patient experiences will be evaluated. The following the key gastrointestinal physician defined toxicity endpoints will be evaluable for this outcome (See appendix II of the full protocol for the definition of grades): diarrhea, vomiting, nausea and abdominal pain. Proportion of patients experiencing any grade 2 or higher gastrointestinal toxicity will be compared between the two arms.

#### Secondary Outcomes

1. **Disease free survival:** This will be defined as the interval between the date of randomization to the documented date of death, any recurrence or a second malignancy (whichever event is detected earliest). Patients who are alive without disease will be censored at the last date of follow up. The actuarial estimate of disease free survival in both arms will be estimated along with 95% confidence intervals.
2. **Patient reported outcome:** Patient reported outcomes will be evaluated using the EORTC QLQ C30 and CX24 questionnaires which will be administered at baseline and subsequently at each week during radiotherapy and at each scheduled follow up visit. The three scales of interest for this endpoint are the global quality of life, C30 summary score (defined by Glessinger et al) and the symptom experience score. These will be calculated from responses using guidance provided by the EORTC group. Mean scores and 95% confidence intervals of mean will be compared between groups at 6 months, 12 months, 24 months and 36 months.
3. **Late Toxicity:** The cumulative incidence and 95% confidence intervals of each of the defined late toxicity endpoints (see Appendix II) will be compared between the two arms.

### Participant Timelines

The following table shows the participant timelines at randomization. Baseline screening and eligibility evaluation will be performed before starting chemoradiation.

**Table 3:**
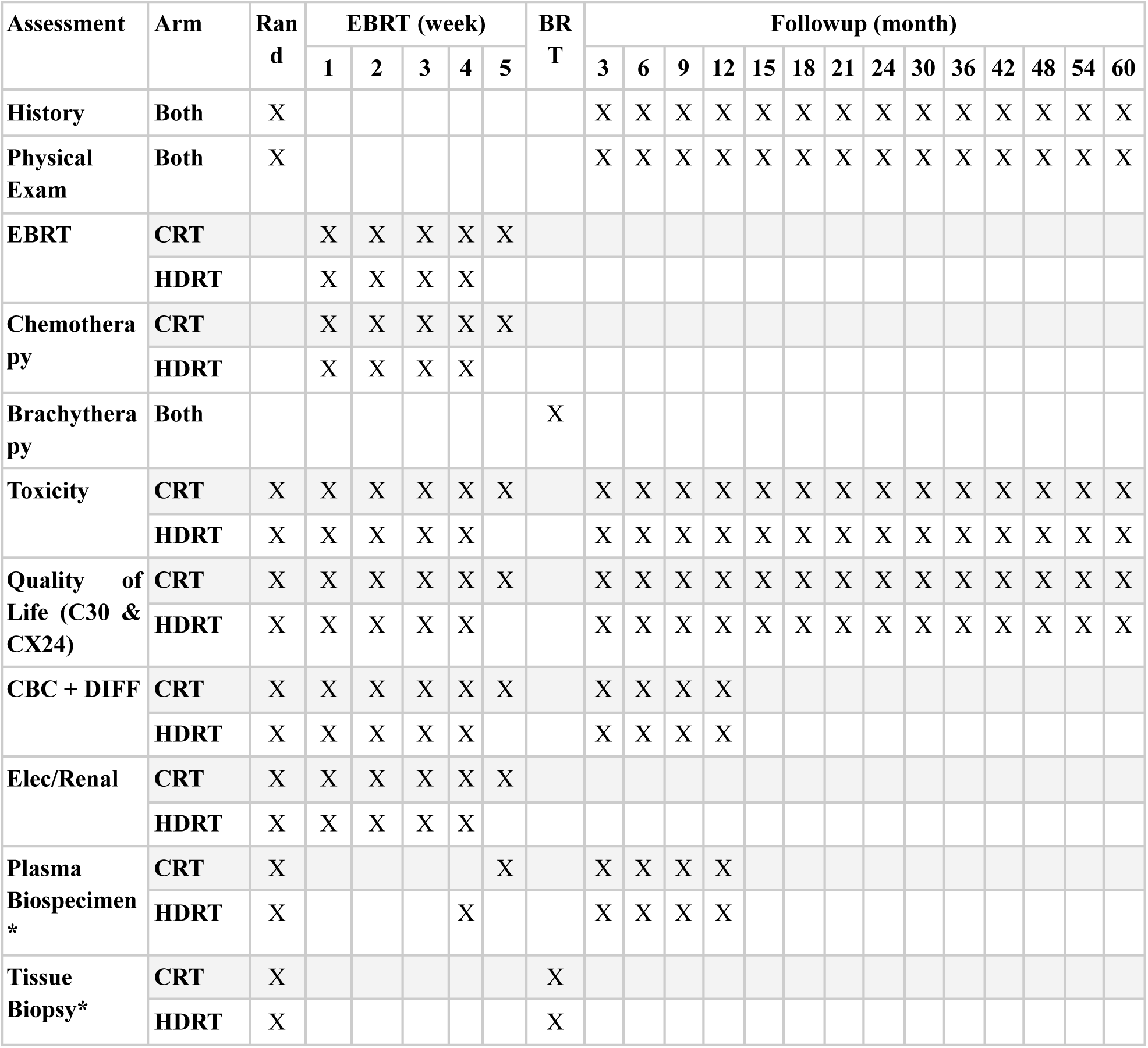
Schedule of Assessments in each arm. * Plasma and tissue biospecimen collection in patients who consent for biobanking only. CRT = Conventional Radiotherapy, HDRT = Hypofractionated Dose Painted Radiotherapy, EBRT = External Beam Radiotherapy, C30 = EORTC QLQ C30, CX24 = EORTC CX24, CBC+DIFF = Complete hemogram with differential counts, Elec/Renal = Electrolytes and renal panel (Serum sodium, potassium, calcium, magnesium, creatinine), BRT = Brachytherapy. Note that chemotherapy will be delivered with concurrent cisplatin at a dose 40 mg/m2 weekly in the CRT arm and 50 mg/m2 in the HDRT arm. Brachytherapy schedule is tailored to deliver a cumulative EQD2 of 80 - 90 Gy_10_ to the high risk clinical target volume.

### Trial Quality Assurance

#### Overview

A multi-layer quality assurance protocol has been planned for the HOPE trial. This will start with site initialization and commissioning and will continue prospectively with virtual multi-center peer reviews for target volume delineation and online image guidance. Additionally as described in the section Radiotherapy Quality Assurance, each plan will undergo individual dose verification before delivery.

#### Site Initialization and Commissioning

Each of the sites will be sent a dummy dataset of three patients with node negative cervical cancer (deidentified). They will be requested to perform segmentation of the target volumes and the organs at risk on this dataset. Each dataset will have the required clinical and diagnostic radiological information also available. The segmentation performed will be reviewed in a virtual mulit-center peer review session where the segmentations will be evaluated with respect to the protocol. Specific attention would be given to ensure that the primary target volume is delineated in the manner expected. The investigators at the center will not only be a part of these peer review meetings but would be actively encouraged to evaluate other center’s contours in an open and transparent fashion. After the review, each center will be provided with a summary of the key modifications needed which they will be making. The revised version of the segmentation would be reviewed. Quantitative spatial overlap metrics for these segments will also be computed like Dice Similarity Index, Hausdorff distance and added path length against the STAPLE contour computed from the modified contours. The results of these quantitative analysis will be communicated to the individual centers also.

The center will also participate in a dedicated planning exercise where dummy DICOM datasets for three other patients will be provided for treatment planning as per the HOPE trial protocol. Centers will be free to use the IMRT planning technique they would prefer but use of IMRT is mandatory in the study. In order to ensure comparability, the PTVs will be provided to the center for planning. Plans will be evaluated against the pre-specified protocol defined constraints and also evaluated in a multi-center virtual meeting with expert physics planners from each center. This would allow easy exchange of knowledge and planning techniques as different centers have access to different types of machines and treatment planning systems. An iterative feedback driven process will be followed for this step also where the modifications requested for the plans will be implemented, reviewed and then formalised by the institution.

Once the plans are deemed acceptable, then delivery quality assurance of each of these plans will be performed by the center. Both absolute and relative dose measurements are required and absolute dose measurement should be done using a licensed and calibrated ionization chamber. The calibration documentation will be reviewed. Once this step is completed, the site will be ready for recruitment in the study.

#### Prospective Peer Review

A prospective multi-center virtual peer review will be organized as is being currently done for the PROPARA trial. This process will be done for each new case recruited in each center for at least the first 15 cases. Each center will be performing the segmentation for the case based on the protocol specified guidelines, and presenting these in a virtual meeting attended by the team of investigators from the trial. We have found that these meetings result in identification of major modifications in the clinical target volumes in 80% of the cases. In addition to the impact on the quality, there is immense educational and training value in these peer review sessions. Peer review meetings will focus on the case selection, review of the imaging and clinical findings as well as evaluate the target volume and organ at risk delineation.

In addition to new cases, these meetings will also be used to review the approved plan for these cases and discuss any difficult planning issues encountered for treatment planning.

#### Multicenter IGRT review

A novel multi-center virtual IGRT review consisting of a team of radiographers from each center will also be implemented as a part of this research protocol. This peer review session will evaluate the on-board volumetric image guidance results for the trial patients focussing on sessions where significant matching issues were identified. We expect that these sessions will have immense value in harmonizing IGRT practices across centers as we have observed with target volume delineation.

### Sample Size

#### Justification for the outcome measures

The primary outcome measure is the acute toxicity specifically grade 2 or higher CTCAE 6.0 defined acute diarrhea, vomiting and nausea. This endpoint has been selected as the primary endpoint because of its relevance to the patient quality of life, as well healthcare resource utilization in the form requirement for treatment interruptions, admissions, parenteral drug administration as well as downstream secondary toxicities like electrolyte imbalance, weight loss, and malnutrition. Dose reduction to the elective pelvic lymph nodes is likely to mostly impact this toxicity endpoint. Additionally, in the trials of cervical cancer where moderate hypofractionation has been employed, acute GI toxicity has been reported to be numerically higher (though none of these studies involved simultaneous dose deescalation to elective nodal volumes).

#### Primary Outcome

The primary outcome measure is to compare the proportion of patients with CTCAE v 6.0 grade 2 or more gastrointestinal toxicity (any of diarrhea, vomiting, nausea) in the two arms. Based on the results of the trials mentioned in the review of literature we can see a reduction in the bowel dose results in a corresponding reduction in the risk of acute grade 2 or higher gastrointestinal toxicity. Based on the results of the literature review cited above, we estimate the rate of grade 2 or higher gastrointestinal toxicity during concurrent chemoradiation will range between 35 - 40% in the control arm.

In-silico planning results of this dose schedule performed at Tata Medical Center suggest that the bowel volume receiving 30 - 45 Gy_10_ can be reduced by 45 - 90% with the new regimen. We therefore expect that the risk of toxicity can be reduced by 40% - 50% further. This translates to an absolute reduction of 14% - 20%.

We assume that the event rate (Grade 2 or higher Gastrointestinal toxicity) will be observed in 35% of the patients in the CRT arm and 21% of the patients in the HDRT arm corresponding to an absolute risk reduction of 14% (relative reduction of 40%).

A group sequential design has been planned with 3 interim analyses planned at 25%, 50% and 75% of the information fractions. Using an asymmetric two-sided group sequential design with non-binding futility bound, 4 analyses, the target sample size is 274, for 80% power, and 5% (1-sided) Type I error. Efficacy bounds derived using a Hwang-Shih-DeCani spending function with gamma = -8. Futility bounds derived using a Hwang-Shih-DeCani spending function with gamma = -2. The sample size was calculated using the gsDesign software version 3.8.0 [18].

The tabular summary of the interim analyses with Z values for efficacy and futility bounds for each interim analysis are described in the table below. The final sample size is rounded to 300 patients to account for dropouts.

**Table 4:**
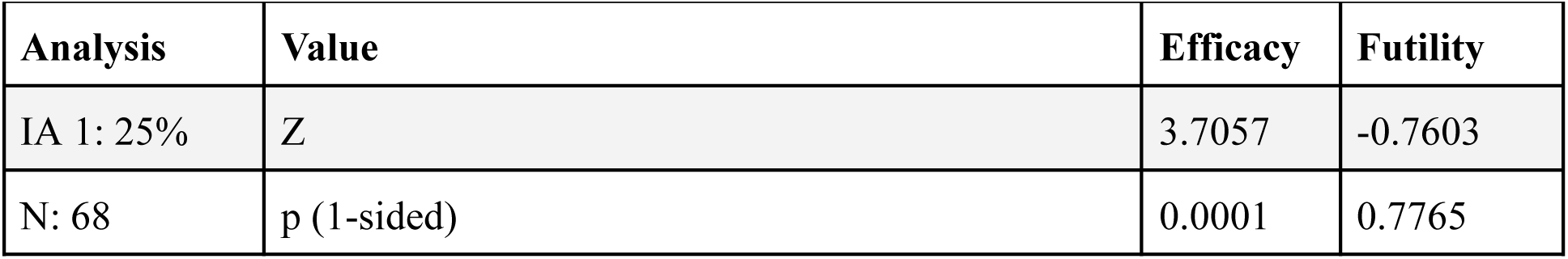
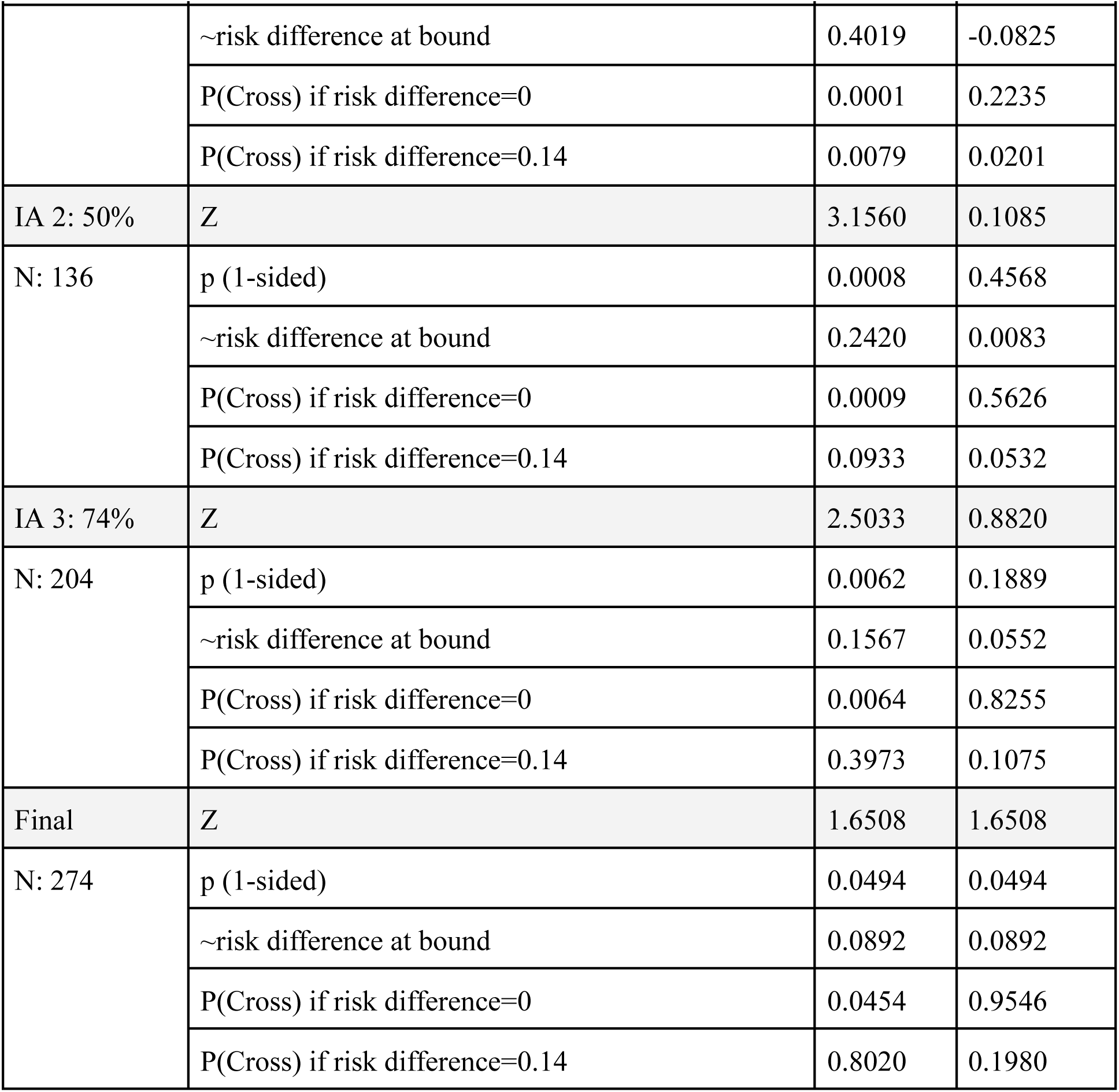
IA is the percent of statistical information (planned sample size) at interim analysis. N = rounded sample size. The nominal p-values indicative of superiority and futility at each time point are shown in the table. The p(Cross) refers to the cumulative probability of cross a bound at or before the given analysis under the null hypothesis.

### Randomization

#### Sequence Generation

The randomization sequence will be generated by an independent statistician who will then put the sequence in the REDCap database. Once added the sequence cannot be changed or modified. The randomisation schedule will be created using computer-generated random numbers before the first participant has been recruited, in a one-to-one ratio.

#### Type of Randomization

Randomization will be performed after the target volume delineation has been reviewed and approved prior to treatment planning. This will be done to minimize bias arising out of differences in the target volume delineation. Randomization will be done in a 1:1 (equal) ratio in one of the two treatment arms stratified by the receipt of neoadjuvant chemotherapy the primary tumor size using permuted blocks of random sizes.

#### Allocation Concealment

Randomization will be centrally and will be only performed after target volume delineation has been reviewed and approved prior to treatment planning. Allocation to the randomized arm will only be revealed before the treatment prescription is given to minimize bias. This will ensure that the target volume delineation and patient selection will not be influenced by the choice of the treatment arm.

#### Implementation

Once the screening is completed and the patient has given consent for the study, the random allocation will be done. The allocation will be done using the REDCap randomization module which will be available at the institutional review board of the Tata Medical Center and will be communicated to the Principal Investigator (PI) or Co-PI over the phone. The randomization module will not be available with the PI / Co-PI to ensure that allocation concealment is maintained. Additionally the trial personnel can only click the randomize button to randomize the patient and have no prior knowledge of the randomization sequence. Access to the randomization sequence will be restricted to the trial statistician who has generated and uploaded the randomization sequence. REDCap automatically logs all randomization related actions and fixes the randomization once performed.

### Data Collection and Methods

Assessments will be administered during the routine follow up visits and will be administered by the assessing doctors and nurses during the treatment and follow up. Patient reported outcome measures (quality of life) assessments will be completed by the patients themselves. For patients who are illiterate, quality of life assessments can be completed by verbally asking the patient about the questions. Paper based case record forms will be obtained from the electronic REDCap data management system in which the centralized study database will be created.

Data will be collected using case record forms designed and hosted on an electronic research data capture tool - REDCap hosted at Tata Medical Center, Kolkata. All participating institutes will also have access to the same REDCap database. REDCap (Research Electronic Data Capture) is a secure, web-based software platform designed to support data capture for research studies, providing 1) an intuitive interface for validated data capture; 2) audit trails for tracking data manipulation and export procedures; 3) automated export procedures for seamless data downloads to common statistical packages; and 4) procedures for data integration and interoperability with external sources [19,20]. In the clinic, electronic data may be captured directly on the electronic CRFs or may be transcribed from paper CRFs completed in the clinic. In case the latter is done, paper records should be saved in a study specific participant file. If patient reported outcomes are collected electronically in a system separate from REDCap, a mechanism must be in place which allows import of the patient reported data in the REDCap database. Please refer to the full protocol for the details on the assessments performed at different timepoints in the trial.

### Data Management

Access to the data will be available to the principal investigator of the study as well as to the co-investigators. The data quality and integrity will be checked at quarterly intervals using the data quality checking system available in Redcap. Patient identifiers will be noted as such in the Redcap database and the same will be used for ensuring that any data exports contain de-identified data only. Institutes will have access to the data of their own institute during the trial phase. Once the trial is completed the main database will be checked for data consistency and quality and then closed for analysis.

### Data Analysis

#### Primary Outcome Analysis

The primary outcome (proportion of patients with acute grade 2 or higher gastrointestinal toxicity - diarrhea, vomiting and nausea) will be compared between the two arms using a log binomial model with randomization arm as the independent variable. The model will be adjusted for the stratification variables - receipt of neoadjuvant chemotherapy (yes versus no) and tumor size (4 cm or less versus higher) as well as for the prognostic baseline variables: age, hemoglobin, and body mass index. Continuous variables like age, hemoglobin and body mass index will be modelled with polynomial expansion to allow for nonlinear relationships. An interaction between age and body mass index will also be modelled. The log binomial model provides us with relative risk estimates which will be reported with 95% confidence intervals. In the event of a convergence failure for the log binomial model, an alternative modelling approach proposed by Zhou et al involving the use of a modified poisson regression with robust error variance will be utilized [21]. For this model the maximum grade of observed toxicity will be taken.

In addition to using the maximum grade method, an ordinal state transition model will also be used to model the weekwise acute toxicities during treatment (during EBRT). If more than one observation is present in a given week, then the maximum grade of the toxicity for the given week will be considered. The analytical methodology will be adapted from Rohde et al [22]. The ordinal state transition models will be developed for the following key toxicity endpoints where the states will be defined using the CTCAE 6.0 grading criteria (Appendix III).

1. Diarrhea
2. Nausea
3. Vomiting
4. Abdominal Pain
5. Non Infective Cystitis
6. Proctitis
7. Neutropenia
8. Thrombocytopenia

State occupancy and state transition probabilities will be computed and visualized empirically as well as from model derived estimates. In the event of a death or treatment discontinuation, the last grade of toxicity will be carried forward. Note that the treatment discontinuation refers to discontinuation of external beam radiotherapy not concurrent chemotherapy. In case of death related to toxicity the grade will be considered as 5. The correlation structure in the data will be evaluated by computing the Spearman correlation between outcomes for each pair of study weeks. The independent variables for the model will include those specified above. The ordinal transition model has two parts - a cumulative probability model for the ordinal data and a transition model for the longitudinal data. A logit link function will be used for cumulative probability models. The transition model will include the week of treatment as a covariate. The derived quantities from such models will be:

1. State occupancy probabilities: A state occupancy probability is the probability that a subject is having a certain grade of toxicity at a certain week of treatment in either arm.
2. Mean time in the state: This is the average time a subject will spend with a certain grade of toxicity.
3. Days difference: This will be the difference in the average number of days with a lower toxicity in the two arms.

#### Secondary Endpoints

##### Disease Free Survival

The disease free survival (interval of time between randomization to that of death, recurrence or second malignancy) will be compared between the two arms using a cox proportional hazards model. The randomization arm, and the stratification variables will be covariates used for this analysis. Patients will be censored at the last date of follow up. As the trial is not powered for detecting a non-inferiority in this outcome, subgroup analyses will not be attempted for this endpoint.

In addition to disease free survival, the cumulative incidences of local, pelvic nodal, para-aortic nodal, locoregional and distant relapse will also be compared between the two arms. Additionally, the overall survival will also be compared and reported. Cumulative incidences of local, pelvic nodal, para-aortic nodal, locoregional and distant recurrences will be compared between two arms using a Fine Gray model with death as the competing event. Local recurrence is defined as any recurrence observed in the cervix, uterus or the adjacent vaginal mucosa. Pelvic nodal recurrence is defined as any recurrence observed in the common iliac, internal iliac, external iliac, obturator, presacral, and parametrial lymph nodes. Paraaortic nodal recurrence is defined as any recurrence observed in para-aortic (retroperitoneal) nodal chain. Note that inguinal nodal, distant vaginal and retrocrural recurrences will be classified as distant recurrences.

##### Quality of Life

Scores will be computed for the global quality of life, 15 item summary score (as proposed by Giesinger et al [23]), and symptom experience scale from the EORTC QLQ C30 and CX24 questionnaires following the prescribed procedure. These scores will be aggregated at 6, 12, 24 and 36 months and represented by medians and interquartile range. Comparison of the mean scores at each time point will be performed using a longitudinal mixed model with randomization group and stratification variables as well as the age as fixed predictors, and time as the random predictor. Model derived estimates at each time point will be visualized and reported in tabular summaries. Additional analyses will focus on the individual symptom and functional scales following the same methodology as outlined.

In addition to this a Persistence of Late Substantial Patient Reported Symptoms (LAPERS) analysis will also be conducted. For this analysis patients with a minimum follow up of 6 months will be included. Patient reported outcomes from 6 months onward will be filtered. A symptom will be regarded as substantial for response categories of 3 or 4 and persistent if the median score of the symptom in the late follow up (beyond 6 months) is ≥ 2.5 (indicating that the substantial symptoms are present in at least half of the late follow up visits). Baseline condition will be defined as the minimum of the scale score from baseline to until 6 months (early follow up period). Progression of a symptom will be defined as a median score at late follow up more than the minimum baseline score. The difference in LAPERS in between each arm will be reported.

##### Late Toxicity

The cumulative incidence of late toxicity related endpoints will be compared between two arms using a Kaplan Meier method and cox proportional hazards model. The first documented incidence of late toxicity will be used for computation of the interval between randomization to that of the toxicity. Multivariable cox proportional hazards models will include the randomization group and stratification variables as covariates. The cumulative incidence of any grade late toxicity as well as moderate to severe late toxicities (Grade 3 or higher) will be presented. Sensitivity analysis will also be conducted by including patients with at least 6 months of documented follow up for these endpoints.

#### Analysis Population

An intention to treat analysis methodology will be adopted. This population includes all randomised participants regardless of whether they were later found to be ineligible, did not adhere to the protocol or were never treated. Additionally a sensitivity analysis of the acute toxicity will be reported for the as treated population of all randomised participants classified according to the actual treatment received.

#### Missing Data

The sample size for the primary outcome has been inflated to account for a 9.5% dropout rate. This rate is similar to the dropout observed during chemoradiation in major randomized controlled trials. Missing data for the primary endpoint may arise due to incomplete treatment or incomplete evaluations. If the patient has missed their treatment, then the last observed grade of toxicity will be used for the primary analysis and will be carried forward for the additional analysis using ordinal state transition model. In case of missed evaluation, a sensitivity analysis dataset will be created which will impute the maximum of the grade of toxicity observed before and / or after the missing observation.

For the evaluation of disease free survival and other disease related endpoints, missing data can arise from loss to follow up. In such a situation, the investigators will attempt to contact the patient using email, telephone and postal contact methods to determine the last status. If the same cannot be elicited then, the last known status will be used. However if the patient has a known recurrence prior to loss to follow up, then as a part of a sensitivity analysis, the overall survival will be computed using the date of last follow up as the date of death. Similarly the cumulative incidences of the local, pelvic nodal, para-aortic nodal, and locoregional recurrences will be estimated by considering that these patients have died as a part of a sensitivity analysis.

### Patient and Public Involvement

No specific patient and public involvement was there for the drafting of the protocol.

## Ethics and dissemination

### Research Ethics Approval

This study will be conducted according to the Note for Guidance on Good Clinical Practice provided by the Central Drugs Standard Control Organization (CDSCO) and in compliance with applicable laws and regulations. The study will be performed in accordance with the principles laid down by the World Medical Assembly in the Declaration of Helsinki 2004. The investigator shall comply with the protocol, except when a protocol deviation is required to eliminate immediate hazard to a subject. In this circumstance the local IRB and the trial CTU should be notified immediately.

### Protocol Amendments

Changes and amendments to the protocol can only be made by the Trial Management Committee. Approval of amendments by the Institutional (IRB) is required prior to their implementation. In some instances, an amendment may require a change to a consent form. The Investigator must receive approval/advice of the revised consent form prior to implementation of the change. The investigator should not implement any changes to, or deviations from, the protocol except where necessary to eliminate immediate hazard(s) to trial subject(s).

### Consent

All patients will be provided with a patient information leaflet which will detail the study purpose, procedures, harms and expected benefits. Written informed consent will be obtained and signed by an independent witness as well as the PI or Co-PI or designated authorized person. The date, time and the name of the person obtaining the consent will be recorded in the medical records. Patients can withdraw consent at any time without ascribing a reason for the same. For patients who withdraw their consent for the trial, data up till the point of consent withdrawal will be retained but no further data will be collected.

### Confidentiality

All study-related information will be stored securely on a trial specific REDCap database which has a password-protected access system and access to the information will be limited to only those that are part of the trial team. All the participant information that is stored in physical format will be in locked file cabinets in areas with limited access. The study data will be accessible to the local IRB for auditing, state and national regulatory bodies as necessary.

### Dissemination Policy

The Trial Management Committee will appoint a Writing Committee to draft manuscript(s) based on the trial data. Manuscript(s) will be submitted to peer-reviewed journal(s). The Writing Committee will develop a publication plan, including authorship, target journals and expected dates of publication. All publications must be circulated to the trial management committee and receive approval prior to submission. Note that for the purpose of approval an email approval is recommended but not mandatory. Additionally trial results will be reported at the clinical trial registry of India as per the extant policies and regulations.

All publications and presentations will be submitted on behalf of an author group. A trial author group composed of all members of the trial management committee as well as any additional contributors will have oversight over each publication. All members in the author group will be considered to have equal authorship in publication and names in the author group will be kept in the alphabetical order using the first name and last name. One of the authors will be the designated corresponding author.

## Supporting information

Main Protocol

## Data Availability

All data produced in the present study are available upon reasonable request to the authors

## Author Contributions

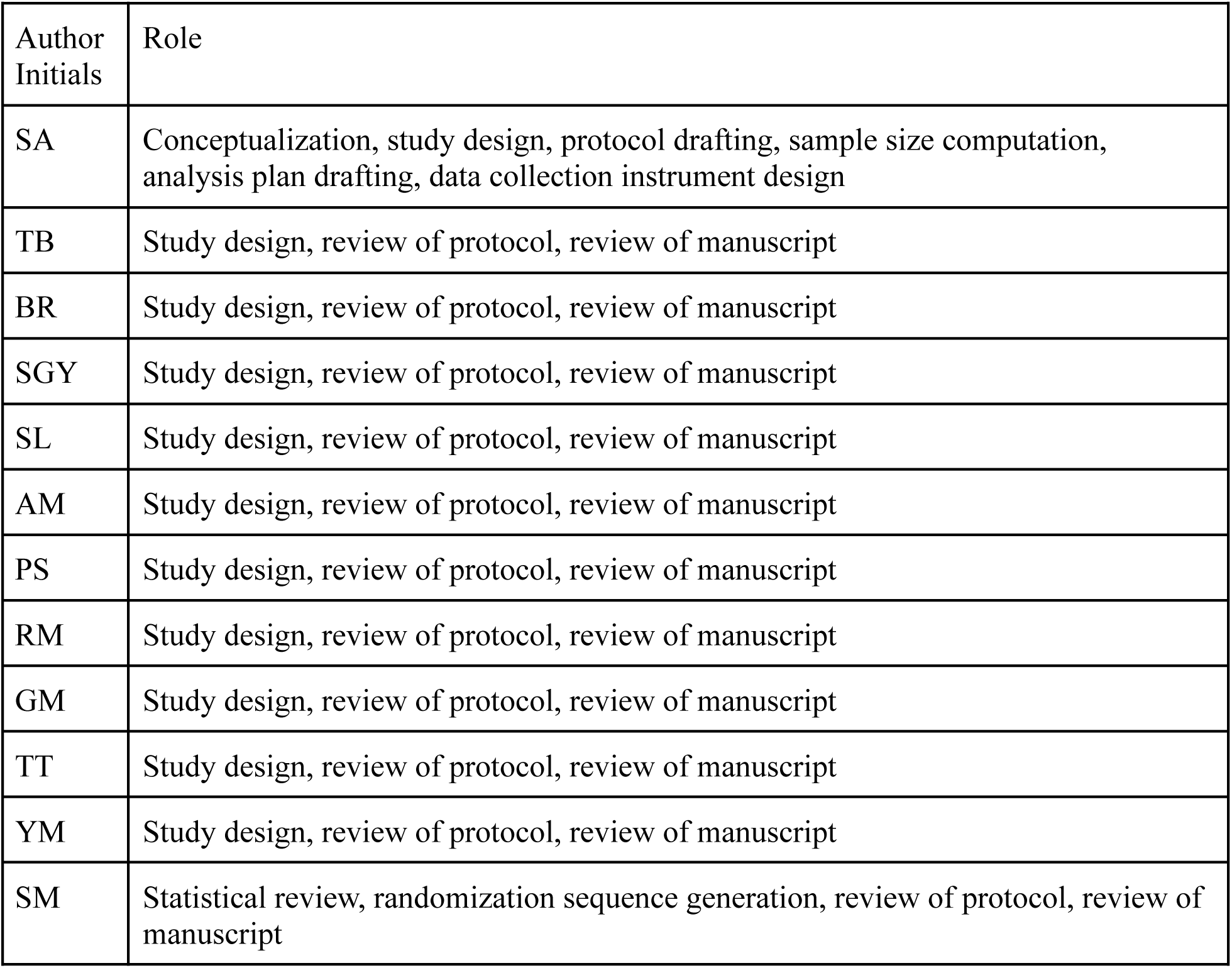

All authors agreed to the manuscript submission. Author SA is the guarantor.

## Full References

[1] National Cancer Registry Programme Investigator Group, Mathur P, Sathishkumar K, Das P, Santhappan S, Sankarapillai J, et al. Cancer incidence and mortality across 43 cancer registries in India. JAMA Netw Open 2025;8:e2527805.

[2] Pötter R, Tanderup K, Schmid MP, Jürgenliemk-Schulz I, Haie-Meder C, Fokdal LU, et al. MRI-guided adaptive brachytherapy in locally advanced cervical cancer (EMBRACE-I): a multicentre prospective cohort study. Lancet Oncol 2021;22:538–47.

[3] De Leeuw AAC, Nout RA, Van Leeuwen RGH, Mans A, Verhoef LG, Jürgenliemk-Schulz IM. Implementation of state-of-the-art (chemo)radiation for advanced cervix cancer in the Netherlands: A quality improvement program. Technical Innovations & Patient Support in Radiation Oncology 2019;9:1–7.

[4] Lindegaard JC, Kirisits C, Schmid MP, Wulff CN, Steen SG, Kristoffersen KB, et al. Impact of Patient Selection on Real-World Outcomes by Using the EMBRACE-II Treatment Protocol in Locally Advanced Cervical Cancer. Int J Radiat Oncol Biol Phys 2025;123:669–80.

[5] Thayer-Freeman C, Washington B, Fabian D, Cheek D, Clair WS, Bernard M, et al. In vitro α/β ratio variations in cervical cancer, with consequent effects on equivalent dose in 2 Gy fraction in high-dose-rate brachytherapy. Adv Radiat Oncol 2025;10:101725.

[6] Maddah Safaei A, Esmati E, Gomar M, Akhavan S, Sheikh Hasani S, Malekzadeh Moghani M, et al. Hypofractionated versus standard chemoradiotherapy in the definitive treatment of uterine cervix cancer: interim results of a randomized controlled clinical trial. J Cancer Res Clin Oncol 2024;150:20.

[7] Dankulchai P, Prasartseree T, Sittiwong W, Thephamongkhol K, Nakkrasae P. Early results of hypofractionated chemoradiation in cervical cancer with 44 Gy/ 20 F vs 45 Gy/ 25 F: A phase II, open-label, randomised controlled trial (HYPOCx-iRex trial). Clin Oncol (R Coll Radiol) 2025;46:103907.

[8] Kobayashi R, Yamashita H, Okuma K, Ohtomo K, Nakagawa K. Details of recurrence sites after definitive radiation therapy for cervical cancer. J Gynecol Oncol 2016;27:e16.

[9] Nomden CN, Pötter R, de Leeuw AAC, Tanderup K, Lindegaard JC, Schmid MP, et al. Nodal failure after chemo-radiation and MRI guided brachytherapy in cervical cancer: Patterns of failure in the EMBRACE study cohort. Radiother Oncol 2019;134:185–90.

[10] Achari RB, Chakraborty S, Ray S, Mahata A, Mandal S, Das J, et al. 18F-fluorodeoxyglucose PET-CT-guided pelvic chemoradiation therapy using helical tomotherapy for locally advanced carcinoma cervix without para-aortic nodal disease: Clinical and patient-reported outcomes from a prospective phase 2 study. J Med Imaging Radiat Oncol 2024;68:624–34.

[11] van den Bosch S, Cox MC, Doornaert PAH, Hoebers FJP, Kreike B, Vergeer MR, et al. Dose de-escalation of elective neck irradiation in head and neck cancer: A secondary analysis of acute toxicity findings from the randomized controlled UPGRADE-RT trial. Radiother Oncol 2025;213:111196.

[12] Nuyts S, Lambrecht M, Duprez F, Daisne J-F, Van Gestel D, Van den Weyngaert D, et al. Reduction of the dose to the elective neck in head and neck squamous cell carcinoma, a randomized clinical trial using intensity modulated radiotherapy (IMRT). Dosimetrical analysis and effect on acute toxicity. Radiother Oncol 2013;109:323–9.

[13] Tsai CJ, McBride SM, Riaz N, Kang JJ, Spielsinger DJ, Waldenberg T, et al. Evaluation of substantial reduction in elective radiotherapy dose and field in patients with human Papillomavirus-associated oropharyngeal carcinoma treated with definitive chemoradiotherapy. JAMA Oncol 2022;8:364–72.

[14] Lépinoy A, Lescut N, Puyraveau M, Caubet M, Boustani J, Lakkis Z, et al. Evaluation of a 36 Gy elective node irradiation dose in anal cancer. Radiother Oncol 2015;116:197–201.

[15] Herring DF. The degree of precision required in the radiation dose delivered in cancer radiotherapy. Brit J Radiol 1971:51–8.

[16] Moos K, Baldinger M, Perez Haas Y, Ludwig R, Looman E, Balermpas P, et al. A tumor control probability model for elective nodal irradiation to balance toxicity and regional tumor control in treatment plan optimization for head-and-neck squamous cell carcinoma. Phys Med Biol 2026;71:035022.

[17] PROPARA Trialist Group. Prophylactic para-aortic irradiation vs pelvic radiotherapy in pelvic node-positive carcinoma cervix in the setting of concurrent chemoradiation: a phase II open-label multi centric randomized controlled trial (PRO-PARA). Trials 2026;27:195.

[18] Anderson K. gsDesign: Group Sequential Design. 2026.

[19] Harris PA, Taylor R, Thielke R, Payne J, Gonzalez N, Conde JG. Research electronic data capture (REDCap)--a metadata-driven methodology and workflow process for providing translational research informatics support. J Biomed Inform 2009;42:377–81.

[20] Harris PA, Taylor R, Minor BL, Elliott V, Fernandez M, O’Neal L, et al. The REDCap consortium: Building an international community of software platform partners. J Biomed Inform 2019;95:103208.

[21] Zou G. A modified poisson regression approach to prospective studies with binary data. Am J Epidemiol 2004;159:702–6.

[22] Rohde MD, French B, Stewart TG, Harrell FE Jr. Bayesian transition models for ordinal longitudinal outcomes. Stat Med 2024. 10.1002/sim.10133.

[23] Giesinger JM, Kieffer JM, Fayers PM, Groenvold M, Petersen MA, Scott NW, et al. Replication and validation of higher order models demonstrated that a summary score for the EORTC QLQ-C30 is robust. J Clin Epidemiol 2016;69:79–88.

