## Supplementary material for "A phase II multi-center, open-label, randomized, parallel-group, superiority study to compare the acute toxicity of Hypofractionated dOse Painted External radiotherapy versus conventional external radiotherapy in patients with carcinoma cervix stage I - IIB. (HOPE)": Main Protocol

**CTRI Registration Number** : CTRI/2026/05/111292

**Title** 1
**Trial Registration Numbers** 6
**Trial Registration Dataset** 7
**Protocol Version** 10
**Protocol Summary** 11
**Funding** 12
**Roles and Responsibilities** 13
Protocol Contributors 13
Trial Sponsors 13
Role of study sponsor and funders 13
Coordinating Centre 13
Committees 13
**Access to Trial Protocol** 13
**Data Sharing** 13
**Background** 14
Epidemiology of Node-Negative Cervical Cancer 14
Pelvic Node Failure in N0 14
Elective Regional Nodal Dose Deescalation 15
Head Neck Cancers 15
Anal Canal Cancer 16
Radiobiological Basis of Radiosensitivity 16
Clinical outcomes of HPV Negative cervical cancer 16
Mechanism of Radiosensitivity in HPV positive cancers 17
Differences between head and neck and cervical cancers 17
Impact of pelvic radiation dose on adverse effects 18
Acute Gastrointestinal adverse effects 18
Acute Haematological Toxicity 19
Fraction size sensitivity in cervical cancer cell lines 20
Randomized trials on Hypofractionation in Cervix 20
Rationale for the current study 20
**Objectives** 22
Trial Run In Period 22
Overall Study Objectives 22
Primary Objective 22
Estimand 22
Secondary Objectives 22
Estimands 23

Translational Objectives 23
**Trial Design** **24**
**Study Setting** **25**
**Eligibility Criteria** **25**
Inclusion criteria 25
Exclusion Criteria 26
Registration 26
Withdrawal Criteria 27
Patient Transfer 27
Trial Center Requirements 27
**Intervention and Comparator** **28**
Summary 28
Interventions 28
Pre-treatment Evaluation 28
Neoadjuvant Chemotherapy 29
External Beam Radiotherapy 29
Timing 29
Equipment 29
Localization, Simulation and positioning 30
Target volume delineation 30
Dose fractionation 32
Dose Specification 32
Treatment Planning 33
Beam Energy 34
Plan Quality Assurance 34
Treatment Delivery 35
Verification Imaging 35
On treatment review 35
Treatment scheduling and gaps 35
Brachytherapy 35
Concurrent Chemotherapy 36
Overall treatment time 36
Treatment therapy Adverse events 36
Expected 37
Common 37
Less Common 38
Treatment Modifications 38

External Beam Radiotherapy 38
Concurrent Chemotherapy 39
Brachytherapy 39
Concomitant Therapy 40
**Outcome 40**
Trial Run In 40
Primary Outcome 40
Secondary Outcomes 41
Translational Outcomes 41
**Participant Timeline 43**
**Sample Size 44**
Justification for the outcome measures 44
Primary Outcome 44
**Recruitment 47**
**Randomization 47**
Sequence Generation 47
Type of Randomization 47
**Allocation Concealment 47**
Implementation 47
**Blinding 48**
**Data Collection and Methods 48**
Screening 48
Baseline Assessments 49
On treatment Assessment 49
Quality of Life Assessment 49
Follow up Assessment 49
End of Study Visit 50
Post Study Closure 50
Consent withdrawal 50
**Data Management 50**
**Statistical Methods 51**
Outcome Comparison 51
Primary Outcome Analysis 51
Secondary Endpoints 52
Disease Free Survival 52
Quality of Life 53
Late Toxicity 53
Analysis Population 53

Missing Data 53
Additional Analyses 54
**Data Monitoring 54**
**Safety Reporting 54**
Assessment of Safety 54
Definitions 54
Adverse Events 54
Serious Adverse Events 55
Suspected Unexpected Serious Adverse Reaction (SUSAR) 55
Reporting SAE 56
**Auditing 56**
**Research Ethics Approval 56**
**Protocol Amendments 56**
**Consent 57**
**Confidentiality 57**
**Declaration of Interests 57**
**Access to Data 57**
**Ancillary and Post Trial Care 57**
**Dissemination Policy 57**
**Clinical Study Report 58**
**References 58**
**Appendix I: Biological Materials 64**
Biobanking Methodology 64
Whole Blood and Plasma Banking 64
Tissue Banking 64
**Appendix II: Definition of Acute Toxicity related endpoints 66**

#### Trial Registration Numbers

Indian Clinical Trial Registry (CTRI) CTRI/2026/05/111292

#### Trial Registration Dataset

| | |
|---|---|
| Primary Registry and Trial Identifying Number | Indian Clinical Trial Registry (CTRI) CTRI/2026/05/111292 |
| Date of Registration in Primary Registry | 2026 |
| Secondary Identifying Numbers | |
| Source(s) of Monetary or Material Support | Extramural funding will be sought |
| Primary Sponsor | Tata Medical Center Kolkata |
| Secondary Sponsor(s) | |
| Contact for Public Queries | Dr Santam Chakraborty |
| Contact for Scientific Queries | Dr Santam Chakraborty |
| Public Title | HOPE: Can four weeks of more precisely targeted radiation cause fewer side effects than a typical five-week treatment in women with primary cervical cancer? |
| Scientific Title | A phase II multi-center, open-label, randomized, parallel-group, superiority study to compare the acute toxicity of Hypofractionated dOse Painted External radiotherapy versus conventional external radiotherapy in patients with carcinoma cervix stage I - IIB. (HOPE) |
| Countries of Recruitment | India |
| Health Condition(s) or Problem(s) Studied | Squamous cell carcinoma Cervix stages I - IIB (FIGO 2018) |
| Intervention(s) | **Experimental arm**: Dose painted interval compressed concurrent chemoradiation will be delivered using external beam radiation therapy with volumetric modulated arc therapy to a dose of 42.5 Gy in 20 fractions over 4 weeks to the primary clinical target volume |

| | |
|---|---|
| | (comprising of the uterus, cervix, paracervical tissue and vagina) and 36 Gy in 20 fractions over 4 weeks to the elective pelvic nodal volumes (external iliac, internal iliac, presacral and common iliac). Concurrent chemotherapy will be delivered using weekly Cisplatin 50 mg/$m^2$. All patients will also receive image-guided brachytherapy to ensure that the high-risk clinical target volume receives a total equivalent dose of at least 80$Gy_{10}$.<br><br>**Control Arm:** Conventional concurrent chemoradiation will be delivered using external-beam radiation therapy with volumetric modulated arc therapy to a total dose of 45 Gy in 25 fractions over 5 weeks to the primary clinical target volume and the elective nodal volumes, as above. Concurrent chemotherapy will be delivered using weekly Cisplatin 40 mg/$m^2$. All patients will also receive image-guided brachytherapy to ensure that the high-risk clinical target volume receives a total equivalent dose of at least 80$Gy_{10}$. |
| Key Inclusion and Exclusion Criteria | Inclusion criteria:<br>1. Age 18 - 70<br>2. Performance stage ECOG 0 - 2<br>3. FIGO 2018 Stage (confirmed using FDG PET/CT or CT Pelvis + MRI Pelvis) I - IIB.<br>4. Squamous cell carcinoma<br>5. Human papilloma virus positive disease as evaluated using p16 Immunohistochemistry<br>6. Absence of concomitant comorbidities which prevent administration of concurrent chemotherapy<br>Exclusion criteria:<br>1. Non-squamous histology including adenocarcinoma or adenosquamous carcinoma<br>2. Absence of p16 positive disease<br>3. Extensive intratuterine involvement or presence of gross pyometra<br>4. Indeterminate or pathological pelvic lymph nodes on imaging. Any visualized lymph node should have ALL of the following criteria to be considered non-pathological:<br>a. Short axis diameter: < 10 mm<br>b. Homogenous Texture<br>c. Smooth Border<br>d. Any shape with fatty hilum OR bean like / oval shape without fatty hilum.<br>All patients must provide written informed consent before trial inclusion. |
| Study Type | Phase II, open-label, parallel group, randomized controlled trial |

| | |
|---|---|
| Date of First Enrollment | July 2026 |
| Target Sample Size | Total 300 (equally allocated in each arm) |
| Recruitment Status | To be started |
| Primary Outcome(s) | 1. Proportion of grade 2 or higher gastrointestinal toxicity (diarrhea, vomiting and nausea) defined using CTCAE 6.0. |
| Key Secondary Outcomes | 1. Three year disease free survival<br>2. Quality of life<br>3. Late toxicity |
| Translational Objectives | 1. Circulating HPV DNA kinetics<br>2. Lymphocyte recovery kinetics |

### Protocol Version

| Version | Date | Change |
|---|---|---|
| 1.0 | 12/02/2026 | Initial version presented to IRB |
| 2.0 | 03/03/2026 | Version after initial IRB review. Added detailed justification of rationale for choosing the dose fractionation regimen as well as the choice of concurrent chemotherapy regimen.<br>SAE reporting extended to include reporting till 3 months post radiotherapy. Additionally reporting timelines clarified to be adherent to institutional timelines. |
| 2.1 | 30/03/82026 | Additional sample size calculations for pelvic nodal failure estimation and power to determine non-inferiority in disease free survival. |
| 2.2 | 30/6/2026 | Clarifications added after ICMR review for trial funding.<br>1. Rationale for choice of concurrent chemotherapy regimen was clarified in detail as well as a discussion on why alternate regimens were not chosen<br>2. Details of trial specific quality assurance that will be performed have been provided.<br>3. TCP modelling for potential loss of efficacy due to dose de-escalation has been added |

### Protocol Summary

The HOPE trial is a Phase II, multi-center, open-label, randomized, parallel-group superiority study comparing acute toxicity profiles of hypofractionated dose-painted external radiotherapy versus conventional external radiotherapy in patients with carcinoma cervix stages I-IIB (FIGO 2018). The trial will be conducted in India with enrollment commencing July 2026.

A total of 300 patients will be randomized in a 1:1 ratio to experimental or control arms. The experimental arm employs dose-painted interval-compressed concurrent chemoradiation utilizing volumetric modulated arc therapy (VMAT), delivering 42.5 Gy in 20 fractions over 4 weeks to the primary clinical target volume and 36 Gy in 20 fractions over 4 weeks to elective pelvic nodal volumes (external iliac, internal iliac, presacral, and common iliac). Concurrent chemotherapy consists of weekly cisplatin 50 $mg/m^2$. The control arm delivers conventional concurrent chemoradiation with VMAT at 45 Gy in 25 fractions over 5 weeks to both primary and elective volumes, with weekly cisplatin 40 $mg/m^2$. Both arms receive image-guided brachytherapy to achieve a total equivalent dose of at least 80 $Gy_{10}$ to the high-risk clinical target volume.

Eligible participants are women aged 18-70 years with ECOG performance status 0-2, histologically confirmed squamous cell carcinoma of the cervix, FIGO 2018 stage I-IIB disease (confirmed by FDG PET/CT or CT pelvis plus MRI pelvis), and HPV-positive disease determined by p16 immunohistochemistry. Exclusion criteria include non-squamous histology, p16-negative disease, extensive intrauterine involvement, gross pyometra, and pathological pelvic lymph nodes meeting specified imaging criteria.

The primary endpoint is the proportion of patients experiencing grade ≥2 gastrointestinal toxicity (diarrhea, vomiting, nausea) per CTCAE 6.0 criteria. Secondary endpoints include three-year disease-free survival, quality of life, and late toxicity. Translational objectives assess circulating HPV DNA kinetics and lymphocyte recovery kinetics. This study evaluates whether interval compression with dose escalation to the primary tumor and simultaneous integrated boost technique can reduce acute toxicity while maintaining therapeutic efficacy in early-stage cervical cancer.

#### Funding

### Roles and Responsibilities

#### Protocol Contributors

The principal investigators have designed and developed the trial protocol.

#### Trial Sponsors

Extramural trial funding will be sought from the various governmental and nongovernmental organizations for conduct of the trial.

#### Role of study sponsor and funders

The study sponsor and funders will have no role in study design, data collection, analysis, interpretation and writing of the final report.

#### Coordinating Centre

Tata Medical Center will serve as the coordinating center for the trial.

#### Committees

A trial management committee composed of the principal investigators from each of the centres will be responsible for the design, conduct and analysis of the trial. This committee will also oversee the trial related quality assurance. The trial management committee will be responsible for implementation of any protocol modifications recommended by the independent data safety and monitoring committee.

The independent data safety and monitoring committee will be made up of members with no direct competing interests.

### Access to Trial Protocol

The final clinical trial protocol will be published in an open access journal and also be made available as a preprint in a globally accessible preprint server.

### Data Sharing

A limited set of deidentified clinical and radiological data consisting of CT scans, MRI, RT plans and structure set objects will be made available globally in the CHAVI repository (https://chavi.ai). Additional data requests will be scrutinized by the trial management committee and shared after obtaining appropriate institutional approval. Members of the trial management committee will have access to the cleaned data which will be hosted on the central REDCap installation at Tata Medical center.

### Background

#### Epidemiology of Node-Negative Cervical Cancer

Cervical carcinoma is a major public health concern and is the second most common cancer affecting women in India, with an estimated annual incidence rate of 11.4 per 100,000 [1]. For patients with locally advanced disease, the standard treatment is external-beam radiotherapy (EBRT) to a dose of 40-50 Gy in 1.8-2 Gy fractions over 23-25 fractions, along with concurrent chemotherapy (CCRT) with Cisplatin (40 mg/m2 weekly for 5 cycles) [2,3]. This is followed by intracavitary brachytherapy with a targeted cumulative high-risk clinical target volume (HRCTV) dose of 80-90 Gy [4]. Recently, with the publication of the INTERLACE trial, neoadjuvant chemotherapy (NACT) with weekly Paclitaxel and Carboplatin before the aforementioned concurrent chemoradiation has become an alternative standard of care [4,5].

Currently, 40%-50% of patients treated with the above protocols present with node-negative disease. For example, in the INTERLACE trial, 51% of patients were node-negative on cross-sectional imaging [5]. Results from a prospective cohort study conducted at the Tata Medical Center showed that 47% of patients were node-negative on FDG PET/CT staging [6]. Similarly, in the multicenter EMBRACE-I cohort study, 47.8% were node-negative (Abdominal CT + Pelvic MRI) [2].

#### Pelvic Node Failure in N0

The incidence of pelvic nodal failure (PNF) in patients with pelvic node-negative disease at presentation is low. In a study from Japan by Kobayashi et al., the pelvic nodal relapse rate after treatment in patients without pelvic lymph nodes at diagnosis (n = 89) was 1 recurrence in the pelvic node at the first relapse [7]. Of 48 patients with pelvic node-positive disease, 6 developed pelvic nodal relapse. However, in this cohort, only 12 patients received EBRT boost to the pelvic lymph nodes. Among the patients with pelvic node-positive disease, all patients had recurrence in an initially involved node [7]. In a report of pelvic nodal relapse from the EMBRACE I study, among the 641 patients with initially pelvic node-negative disease, a total of 23 patients (3.58%) had pelvic nodal relapse [8]. Among the patients with initial pelvic or para-aortic node-positive disease (n = 697), pelvic nodal relapse was observed in 61 patients (8.75%). Isolated pelvic nodal failure in the absence of other sites of failure was observed in 2% of the patients with pelvic node-negative disease and 3% of patients with node-positive disease [8]. The authors note that the upper border of the pelvic radiotherapy field was not strictly defined, implying that the common iliac chain may not have been adequately covered. A subsequent analysis by the same group identified the following key variables, which indicated a higher risk of nodal failure (both pelvic and para-aortic): initial node positivity, tumor width, and hemoglobin nadir [9]. The presence of local failure was also associated with a higher risk of nodal recurrence. In the cohort study conducted at Tata Medical Center, among patients staged with FDG PET/CT, the actuarial cumulative incidence of pelvic nodal failure was 3% at 5 years [6]. We have also audited the outcomes of patients with cervical carcinoma treated between 2019 and 2022 at Tata Medical Center (unpublished data). In this audit, a total of 97 patients with stage I-II squamous cell carcinomas were identified. Among these patients, one pelvic failure (cumulative incidence : 1.1%) was identified at a median follow-up of 45 months.

Beadle et al analyzed outcomes of 198 patients with carcinoma cervix treated at MD Anderson Cancer Center who had a regional recurrence without a local recurrence [10]. During this period, a total of 1894 patients were treated, indicating an incidence of isolated regional nodal failure (pelvic, para-aortic, and inguinal) of 10.45% [10]. However, this cohort of patients was primarily treated with definitive radiotherapy delivered with conventional portals, without concurrent chemotherapy (10.6%) [10]. Of the 180 patients with documented relationships between field borders and regional recurrence, 75 (42%) had marginal recurrence without in-field recurrence. Overall, 59 patients experienced a single in-field recurrence, and 46 experienced both in-field and marginal recurrence. This publication also indicates that true in-field pelvic relapse is an uncommon event (5.5%).

#### Elective Regional Nodal Dose Deescalation

Given the low incidence of pelvic nodal relapse in patients with node-negative cervical cancer, dose-deescalation to the elective lymph node basins can potentially reduce toxicities. While there is no direct evidence for this approach in cervical cancer, increasing evidence is emerging from the treatment of other Human Papillomaviruses (HPV) associated squamous cell cancers like anal cancer and head and neck cancers.

##### Head Neck Cancers

For Head and neck cancers, the following table summarizes the key evidence underlying the hypothesis that dose-deescalation to the elective nodal volume may be safe

| Autho r (Year) | N | Trial Type | HPV+ ve | Elective Dose | EQD2 (α/β 3) | EQD2 (α/β 10) | Outcomes |
|---|---|---|---|---|---|---|---|
| Bosch et al (2025) [11] | 300 | RCT | 59.8% (117 OPX) | 43 Gy (33 - 34#) | 37.1 Gy | 40.5 Gy | 2 year Regional recurrence 4.9% vs 4.3% (Exp vs Control) |
| Nuyts et al (2013) [12,13 ] | 200 | RCT | 20% (83 OPX) | NA | NA | 40 Gy | 2-year Regional RFS: 13% vs 5% (Exp vs Control) |

It is noteworthy that in both randomized trials, patients with node-positive disease were included, and not all patients were HPV positive. In the study by Nuyts et al., 11 patients experienced regional nodal recurrence in the experimental arm, of whom 2 had recurrence within the elective nodal volume. In the same trial, of the 6 patients with nodal recurrence in the control arm, 1 had a recurrence in the elective nodal volume [12,13]. Furthermore, Sher et al. reported the results of a single-arm phase II study enrolling 67 patients with head and neck cancers (49 oropharynx), in which no elective nodal radiation was delivered[14]. At a median follow-up of 33 months, no solitary nodal recurrences in the elective node were observed. For HPV positive disease, Tsai et al reported on the outcomes of a cohort of HPV+ve oropharyngeal cancer (n = 276) where an elective nodal EBRT dose of 30 Gy (in 15 fractions, EQD2 = 30 Gy) was used. At a median follow-up of 26 months, 8 patients experienced locoregional recurrence, all at the site of primary or nodal gross disease [15]. This translated into a 2-year locoregional control of 97%.

##### Anal Canal Cancer

In the case of Anal cancer, another classical malignancy associated with HPV infection, an elective nodal radiation dose between 36 - 40 Gy is standard of care [16]. Lepinoy et al presented outcomes of 142 patients with anal canal cancer treated with an elective inguinal and pelvic nodal radiotherapy dose of 36 Gy followed by a boost to involved nodes and gross disease [17]. These patients had an inguinal control rate of 98.5% and a nodal control of 96%. The PLATO ACT4 randomized controlled trial is investigating a low-dose radiotherapy regimen for patients with localized anal canal cancer [18]. The experimental arm doses have been de-escalated to 34.5 Gy in 23 fractions to the elective nodal volume and 41.4 Gy in 23 fractions to the gross disease. Early data are encouraging, with a clinical complete response rate of 84% in the control and 85% in the experimental arm. Lower-dose radiotherapy resulted in improved sexual function at 6 months. Treatment compliance and acute toxicities were also reduced.

#### Radiobiological Basis of Radiosensitivity

HPV is a non-enveloped, double-stranded, DNA virus that infects epithelial cells. In a subset of patients infected with high-risk types, HPV DNA can integrate into the host genome [19]. The viral E6 and E7 genes subsequently interact with the TP53 and Retinoblastoma pathways and function as oncogenes, promoting proliferation, defective DNA damage repair, accumulation of downstream mutations, and cellular immortalization [19,20]. In the following sections, we evaluate the biological and mechanistic basis underlying increased radiosensitivity in HPV-positive cervical cancers.

##### Clinical outcomes of HPV Negative cervical cancer

While the vast majority of cervical cancers are etiologically linked to HPV infection, a small subset of patients are HPV independent [21]. False HPV negativity may arise due to assay limitations and biological mechanisms like infection by non-high-risk types, loss of HPV DNA fragments during integration, and latent HPV infection [22]. Results from some observational studies have demonstrated that these HPV independent cancers are associated with a high risk of lymphatic metastases and poorer overall survival and disease-free survival [23–26]. Indeed, as demonstrated by Mkrtchian et al., HPV negativity is an independent prognostic factor, even after adjustment for stage and age [26].

#### Mechanism of Radiosensitivity in HPV positive cancers

Several mechanisms have been evaluated that may explain the higher radiation sensitivity and radiocurability of HPV-positive cancers [27]. Kimple et al demonstrated that HPV+ve head neck cancers have higher intrinsic radiation sensitivity with SF2 values of 0.22 as compared to HPV-ve cancers with SF2 values of 0.59 ($p < 0.001$) [28]. HPV+ve cells tended to have a prolonged G2-M phase arrest and had increased apoptosis following exposure to ionizing radiation as compared to HPV-ve cell lines. Additionally, Rietbergen et al. reported a higher percentage of cells expressing CD98 and CD44 (indicative of a cancer stem cell lineage) in tumor biopsies from HPV-negative head and neck cancers than in HPV-positive cancers [29]. Additionally, some studies have demonstrated differences in the immune regulatory landscape of the tumor microenvironment in HPV+ve cancers compared to HPV-ve cancers [30].

#### Differences between head and  neck and cervical cancers

Although it is tempting to believe that HPV mediated mechanisms of oncogenesis are similar between cervical and oropharyngeal cancers, there are several areas of difference in the biology. These were reviewed by Martinelli et al., and are summarised below [31]:

| Feature | HPV+ve Oropharyngeal Cancer | HPV+ve Cervical Cancer |
|---|---|---|
| Anatomical origin | Primarily originates from the reticular epithelium of tonsillar crypts | Arises in the transformation zone between the squamous epithelium (ectocervix) and the columnar epithelium (endocervix) |
| Cancer stem cell markers | Infected progenitor cells consistently express CD44 and ALDH1 | Infected reserve cells show variable expression of these markers, preventing definitive identification |
| Unique mutations | Involved in chromatin remodelling, immune regulation, and DNA repair, like RB1, FGFR2, FGFR3, MLL2, MLL3, ASXL1, NOTCH1, ATM, BRCA1, NF1, FLG, BRCA2, LRP1B, HRAS, TRAF3, DDX3X, TPRX1, CYLD, RIPK4, UBR5 | Involved in cell signaling, growth, and differentiation, like EGFR, SMAD4, ERBB2, ERBB3, ELF3, TGFBR2, CREBBP, MAPK1c, CBFB, ARID1A, NFE2L2, CASP8, STK11, SHKBP1, LKB1, NOL7 |
| Specific | Involved in immune signaling and | Involved in focal-adhesion and |

| | | |
|---|---|---|
| Amplications | self-renewal. 3q26.33 (SOX2), 3q27.1 (KLHL6), 3q27.3 (BCL6), 5p13.1 (RICTOR), 8q24.21 (MYC), 11q13.3 (FGF19, FGF3, FGF4), 14q32.33 (AKT1) | growth factor signalling. 3q28 (TP63, LPP), 3q24.1 (TGFBR2), 18q21.2 (SMAD4), 7p11.2 (EGFR) |
| Specific deletions | 4q31.3 (FBXW7), 13q14.2 (RB1), 14q32.32 (TRAF3), Xp11.3 (KDM6A) | 3p14.1 (FOXP1) |
| HPV Integration sites | RAD51B, MACROD2, NR4A2, KLF5, KLF12, MYC, TP63, 9p24.1 region (PDL1, PDL2, PLGRKT) | POU5F1B, FHIT, KLF12, KLF5, LRP1B, LEPREL1, HMGA2, DLG2, SEMA3D |
| E6 mutation rates | 18.5% of tumors have E6 mutations | 2.0% of tumors have E6 mutations |

*Table 1: Biological differences between HPV mediated cervical squamous cell carcinoma versus oropharyngeal cancers. Adapted from Martinelli et al. [31]*

#### Impact of pelvic radiation dose on adverse effects

##### Acute Gastrointestinal adverse effects

Concurrent chemoradiation is associated with a significant burden of acute and late morbidity. However, dose reduction is associated with reduced acute and late toxicity. This is immediately apparent when we evaluate trials comparing intensity-modulated radiotherapy with three-dimensional radiotherapy in cervical cancer. The acute GI toxicity outcomes of the recently reported randomized controlled trials conducted in patients treated with definitive radiotherapy in India are presented in Table 2.

| Author (Year) | N | Dosimetric Parameter | Technique | | Toxicity Outcome (Grade 2+) | Technique | |
|---|---|---|---|---|---|---|---|
| | | | 3DCRT | IMRT | | 3DCRT | IMRT |
| Rai et al (2024)[32] | 200 | Bowel V45 | 426 | 81.59 | Lower GI | 26.9% | 21.45% |
| | | | | | Vomiting | 28% | 20% |
| Kapoor et al (2022)[33] | 80 | BowelV40 | 251 | 187 | Lower GI | 90% | 42.5% |
| Naik et al (2016)[34] | 40 | BowelV45 | 227 | 132 | Lower GI | 45% | 20% |
| | | | | | Vomiting | 35% | 15% |
| Gandhi et al | 44 | BowelV45 | 417 | 199 | Lower GI | 63.6% | 31.8% |

| | | | | | | | |
|---|---|---|---|---|---|---|---|
| (2013)[35] | | | | | Vomiting | 36.4% | 9.1% |

*Table 2: Summary of dosimetric reduction and the impact on the toxicity reported in randomized trials of IMRT in the definitive treatment of carcinoma cervix. V45 = Absolute volume receiving a minimum dose of 45 Gy*

As shown in the table, reducing the dose to the bowel bag reduces lower GI toxicity and vomiting. Specifically, across these four trials, the weighted-average reduction in the absolute volume of bowel receiving a minimum dose of 45 Gy is 60.9%, and this translates into a weighted-average relative risk of lower GI toxicity of 0.65 and vomiting of 0.60.

#### Acute Haematological Toxicity

Acute haematological toxicity arises from the large volume of pelvic bone marrow irradiated during concurrent chemoradiation. Multiple studies have evaluated the impact of dose reduction using intensity modulated radiotherapy (IMRT) [36–40]. A summary of the key dosimetric and haematological toxicity outcomes is summarised below in Table 3 from the two large randomized trials evaluating pelvic bone marrow sparing IMRT in patients treated with definitive chemoradiation.

| Author (Year) | N | Bone marrow delineation | Anemia (Grade 2+) | | Neutropenia(Grade 2+) | | Thrombocytopenia(Grade 2+) | |
|---|---|---|---|---|---|---|---|---|
| | | | STD | PBMS | STD | PBMS | STD | PBMS |
| Li et al (2024) [37] | 242 | Whole bone | 30.6% | 17.6% | 66.7% | 37.2% | 33.1% | 20.7% |
| Huang et al (2020) [38] | 116 | Low density region | 13.4% | 4.9% | 43.9% | 13.4% | 0% | 4.8% |

*Table 3: Summary of randomized trials evaluating pelvic bone marrow sparing Intensity modulated radiotherapy in cervical cancer patients. STD = Standard Intensity modulated radiotherapy, PBMS = Pelvic bone marrow sparing Intensity modulated radiotherapy.*

It is noteworthy that in the trial reported by Li et al., where three doses were allowed (45 Gy, 50 Gy and 50.4Gy), the incidence of grade 3 haematological toxicity excluding lymphopenia were 60%, 68.6% and 68% in the control groups (without pelvic bone marrow sparing) and 7%, 17% and 38% in the experimental arm (with pelvic bone marrow sparing) [37]. In both studies, pelvic bone marrow doses were reduced with the use of pelvic bone marrow sparing IMRT.

#### Fraction size sensitivity in cervical cancer cell lines

Traditionally, cervical cancer is considered to have an alpha:beta ratio (signifying sensitivity to fraction size) values between 10 - 12 and these have been traditionally used for radiobiological dose calculations. However a recent review compiled the results of alpha:beta ratio values for 98 cervical cancer cell lines from 37 papers [41]. They found that the published results were best fitted by a log-normal distribution with the most probable value of 4.3 Gy. The average value for cervical cancer was 8.05 Gy. Both of these values indicate a higher fraction size sensitivity, which may indicate a role for evaluation of hypofractionated radiotherapy.

#### Randomized trials on Hypofractionation in Cervix

Safaei et al reported a randomized controlled trial (n = 59) comparing the efficacy of hypofractionated radiotherapy to conventionally fractionated radiotherapy (45 Gy / 25 fractions / 5 weeks) [42]. All patients underwent three dimensional conformal radiotherapy. In the hypofractionated radiotherapy arm, a dose of 40 Gy in 15 fractions (3 weeks) with weekly Cisplatin to a dose 40 $mg/m^2$ (3 cycles) was delivered. This non-inferiority trial compared the response (defined on MRI) at 3 months and in an interim analysis demonstrated that complete response was 72% in the experimental arm as compared to 74.1% in the control arm ($p = 0.13$). Grade 3 acute toxicity was higher in the patients receiving hypofractionated radiotherapy (44.8% vs 30%, $p= 0.032$) [42].

Dankulchai et al. reported the results of the HYPOCx-iRex Trial (n = 41) which compared a hypofractionated radiotherapy regimen of 44 Gy in 20 fractions (4 weeks) to a conventional regimen (45 Gy in 25 fractions) [43]. Concurrent chemotherapy was administered at a dose of 40 $mg/m^2$ for 5 cycles in both arms. Both arms received treatment with image guided IMRT. Grade 3 acute toxicity (CTCAE) was higher in the hypofractionated radiotherapy (43%) as compared to the conventional fractionation (32%). Similar results were observed when patient reported outcomes were compared (29% vs 11%). This difference was primarily driven by excess GI toxicity. The trial was also designed as a non-inferiority trial focussing on late toxicity and the early results suggested no difference in tumor outcomes though para-aortic nodal control was better in the hypofractionated radiotherapy. Remaining endpoints like local control and disease free survival were numerically better in the hypofractionated radiotherapy arm.

Summarizing, both trials demonstrated an increased rate of acute toxicity when hypofractionated radiotherapy delivered particularly in the form of higher grades of GI toxicity even when radiotherapy has been delivered using modern conformal technique.

#### Rationale for the current study

As summarised above, the incidence of pelvic nodal failure in patients with node negative carcinoma cervix at presentation is quite low. Despite the use of IMRT, a substantial portion of patients continue to experience acute gastrointestinal and haematological toxicities. Majority of the dose to the pelvic bone marrow and the bowel results from the regional nodal irradiation performed. Radiobiological and clinical data exists from other HPV associated cancers that dose reduction to elective nodal volume does not compromise outcomes. Hypofractionated radiotherapy has been evaluated and has shown encouraging outcomes though at the cost of increased acute toxicity.

Given the low incidence of regional nodal failure and potential efficacy of reducing the overall treatment time demonstrated in the HYPOCx-iREX trial, this study will evaluate a dose painting approach incorporating a dose reduction for the elective nodal volume (36 Gy in 20 fractions at 1.8 Gy per fraction) whilst simultaneously delivering a moderately hypofractionated dose to the primary disease (42.5 Gy in 20 fractions at 2.12 Gy per fraction) to reduce the overall treatment time. The primary dose is chosen to be isoeffective to the standard 45 Gy in 25 fraction In order to ensure that the cumulative dose of 200 mg/m$^2$ cisplatin is delivered in both arms, a dose of 50 mg/m$^2$ weekly cisplatin will be administered during radiation per cycle. Additionally, this regimen allows a dose of 1.8 Gy to be delivered to the elective nodal volume. A lower dose per fraction would be required if a five week regimen is adopted which may be detrimental. In essence an experimental arm with dose deescalation over 5 weeks would involve deescalation of both the total dose and and the dose per fraction. The current regimen is a deescalation only for the total dose and not the dose per fraction. It is noteworthy that for most squamous cell carcinomas, the dose response curve between the dose per fraction region of 1.4 - 1.8 Gy shows a very steep reduction in the response probability below 1.8 [44].

The dose of 50 mg/m$^2$ weekly has been used in several studies in the past [45–48]. We hypothesize that the dose-painted hypofractionated treatment regimen will reduce the acute toxicity allowing this modest dose escalation and be non-inferior in terms of disease free survival. The cumulative dose of Cisplatin is kept equivalent in both arms which is the considered as the primary determinant of outcomes [49]. Majority of the chemotherapy dose interruptions occur after 4 cycles of chemotherapy have been administered, hence this regimen may allow us to deliver a higher cumulative dose of cisplatin also. The investigators considered alternative regimens like delivering 40 mg/m2 weekly Cisplatin for four cycles concurrent with the hypofractionated arm and delivering one fraction after external beam radiotherapy or omitting it. The former choice was not deemed to be optimal as it did not serve the purpose of radiosensitization and the latter choice was not accepted as the cumulative dose of cisplatin would be lower and lead to a potential loss of efficacy. The investigators appreciate that this would necessarily be comparing two different schedules of concurrent chemotherapy. However the concurrent chemoradiation regimen in this study should be considered as an integrated regimen with radiotherapy rather than in isolation as in practice we would not treat patients without concurrent chemotherapy.

#### Tumor Control Probability (TCP) Modelling

When ever dose deescalation is contemplated for elective nodal volumes, a potential loss in the tumor control probability exists. In the current study, the elective nodal dose is being de-escalated by about 19%. While this is lower than what has been done in Head Neck cancers, a formal modelling of the TCP loss has been conducted. For elective nodal volumes the TCP is not a direct function of the dose as a significant proportion of patients do not have disease in the elective nodal region and therefore the TCP by definition is 1. A special formalism has been proposed by Moos et al to allow TCP computation in elective nodal volumes (which do not harbour gross nodal disease) [50]. The TCP for a elective nodal volume receiving a homogenous dose of D is given by the formula:

$TCP(D, Q) = Q * TCP^{+}(D \mid D_{50}\text{^}LNL, m_{50}\text{^}LNL) + (1 - Q) * 1$

Where,

*Q = Probability of occult metastases*

*$TCP+(D|D_{50}\text{^}LNL, m_{50}\text{^}LNL)$ = Conditional probability of controlling an involved LNL*

*D50 = Dose which results in 50% control*

*m50 = Slope of the TCP at D50*

The parameters of D50 and m50 have been taken from Okunieff et al who defined these parameters for squamous cell carcinomas [51]. Using this formula we can compute the predicted loss of TCP for different risks of elective nodal failure as compared to conventional dose fractionation schedules. This loss of TCP is shown is plotted below.

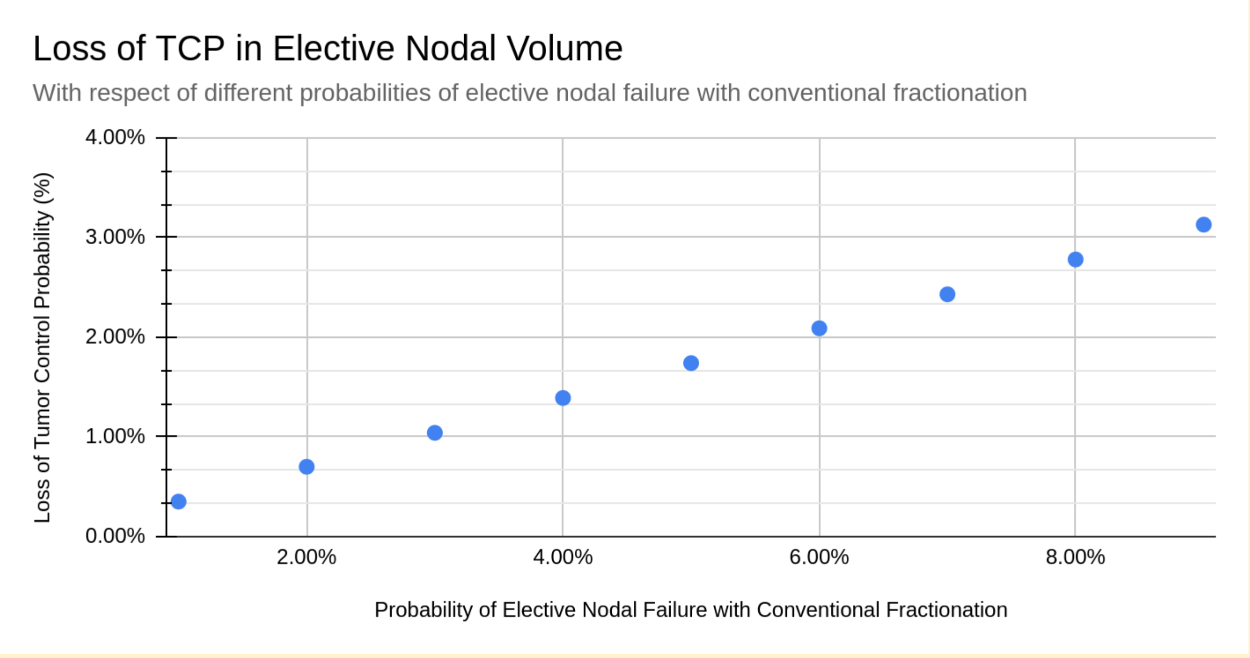


As as can seen the predicted loss of TCP with the proposed dose de-escalation is 3.13 % only when the probability of elective nodal failure is 9% with the standard regimen. However as shown in the literature review this rate is realistically less than 5% and the predicted loss of TCP is 1.74% or less when this is the case.

##### Estimands

| **Objective** | **Population** | **Treatment Groups** | **Endpoint** | **Summary Measure** | **Intercurrent Event** |
|---|---|---|---|---|---|
| Disease Free Survival | Full randomized population | Patients will be compared as randomized | Interval of time between the randomization to any disease recurrence, death due to any cause or development of a second primary | Hazard ratio with 95% confidence intervals | Treatment Policy |
| Quality of Life | Full randomized population | Patients will be compared as randomized | Mean global health related quality of score and symptom experience scores | Difference in means with 95% confidence intervals | Treatment policy |
| Late Toxicity | Full randomized population | Patients will be compared as randomized | Cumulative incidence of CTCA 6.0 defined late toxicity outcomes like fatigue, abdominal pain, proctitis, cystitis, vaginal toxicity, bowel toxicity and bone fractures | Hazard ratio with 95% confidence intervals | Treatment policy |

Treatment policy handling of intercurrent events implies that the occurrence of intercurrent events which result in treatment discontinuation or interruptions will not result in exclusion of the patients from the analysis of the endpoint. As late toxicity related events occur after completion of the treatment hence they will be handled as per treatment policy strategy.

#### Translational Objectives

Two translational studies have been planned in tandem which will synergise with the clinical outcomes.

1. **Circulating HPV DNA kinetics**: To compare the Human papilloma virus (HPV) circulating cell free DNA levels between patients receiving HDRT vs CRT to determine if the use of HDRT results in differential kinetics of HPV cfDNA clearance post

treatment. This is particularly pertinent given the dose deescalation being performed in the HDRT arm.

2. **Lymphocyte Recovery Kinetics**: Dose deescalation as employed in the HDRT arm may translate into better and faster lymphocyte recovery post treatment. The duration of persistence of lymphopenia has been shown to be associated with poorer outcomes in several studies [52–58]. Comparison of the lymphocyte recovery kinetics in between the two arms will be performed to determine if HDRT results in better lymphocyte recovery post treatment. The key endpoint of interest will be the nadir lymphocyte count defined as the lowest level of lymphocyte count observed in a given patient as well as the time spent with grade 2 or higher lymphocytopenia (< 800/mm$^3$).

#### Trial Design

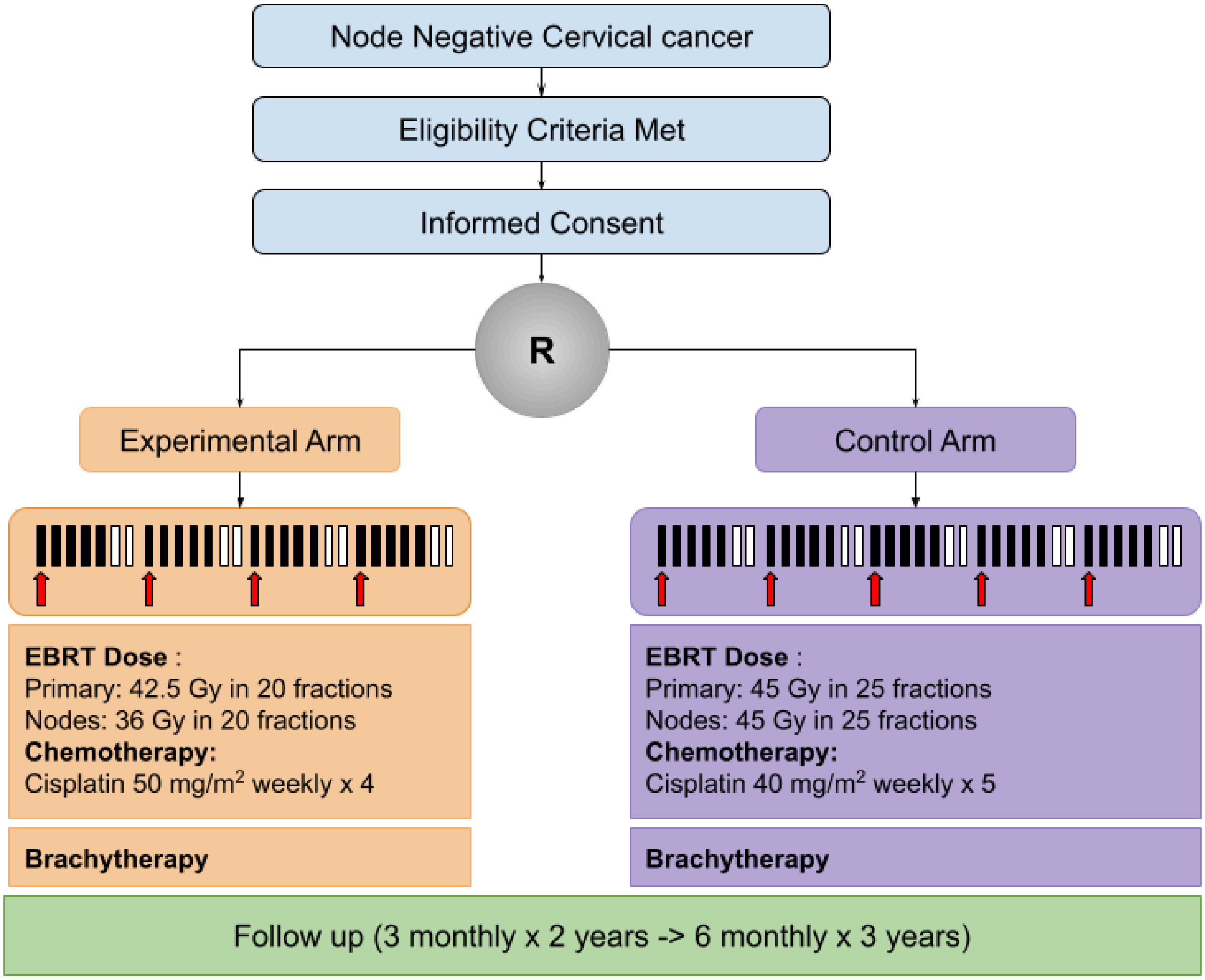


*Figure 1: Graphical representation of the study schema*

The trial design is an open label, parallel group, two arm, randomized controlled trial. An initial trial run-in phase will be included in the design to establish that HDRT can be delivered safely in

these patients.

#### Registration

Subjects must meet all inclusion criteria and none of the exclusion criteria to be eligible for this trial/study. There will be no exceptions to these eligibility requirements at the time of registration. All enquiries about eligibility should be addressed by contacting the Clinical Trials Unit (CTU) at Tata Medical Centre prior to registration.

Subjects must be registered before starting study treatment. Requests for registration will only be accepted from authorized investigators and their designates at sites that have received ethics approval. Treatment should be planned to start within 28 days after registration. Registration should be done only after all screening assessments have been performed and the responsible investigator has both verified the subject's eligibility, and signed the completed registration form. Once the registration process has been completed, the subject will be assigned a subject study number. Written registration confirmation will be provided to the site.
Individuals may only be registered once in this trial.

#### Withdrawal Criteria

Participation in this study is voluntary. Patients will be able to withdraw anytime. A patient may withdraw, or be withdrawn from study treatment for the following reasons:
Intercurrent illness which prevents further follow up
Withdrawal of consent for treatment by patient
Any alterations in the patient's condition which justifies the discontinuation in the investigator's opinion
If a patient or investigator decides to stop the study treatment then the patient's health status will be periodically reviewed via continued study visits or phone contact, or from their general practitioner or medical records to allow collection of outcomes data.
Follow-up assessments including completion of the quality of life questionnaires should still be completed if the patient is willing.
In case a patient withdraws from the study entirely, the effective date of the notification will be the date on which their withdrawal is received by the study team. No information about the patient will be collected from that point in time onwards but any information collected prior to that date can be used and forms part of this study.

#### Patient Transfer

As far as possible the patient should remain under the care of the treating team. For patients moving from the area during follow up, every effort should be made for the patient to be followed up at another participating trial centre and for this trial centre to take over responsibility for the patient. A copy of the patient Case Record Forms (CRF) will need to be provided to the new site after appropriate patient consent. Until the new centre agrees (in writing) to take over responsibility, the patient remains the responsibility of the original centre.

#### Trial Center Requirements

Given the pragmatic design of this study, trial centers should be high volume centers treating at least 30 cervix cancer patients annually. They should have facilities for surgery, radiotherapy, chemotherapy and working pathology and radiology services. The trial centres must fulfil the trial quality assurance requirements as outlined in the Radiotherapy Quality Assurance (RTQA) document.

#### Intervention and Comparator

##### Summary

Patients will be randomized to HDRT and CRT in a 1:1 ratio. Patients in both arms will be treated with image based intracavitary brachytherapy and then be reviewed for follow up. The first follow up will be performed at 6 weeks after treatment completion. Subsequent followups will be conducted three monthly for the first couple of years and six monthly thereafter till five years.

##### Interventions

1. **Control Arm (CRT)** : Patients will undergo radiotherapy to the whole pelvis (primary and elective clinical target volumes) to a dose of 45 Gy in 25 fractions (1.8 Gy per fraction) delivered over a period of five weeks along with concurrent chemotherapy using Cisplatin at a dose of 40 mg/m2 weekly (capped at a maximum of 70 mg per cycle).
2. **Experimental Arm (HDRT)** : Patients will under radiotherapy to a dose of 42.5 Gy in 20 fractions (2.12 Gy per fraction) to the primary clinical target volume and a dose of 36 Gy in 20 fractions (1.8 Gy per fraction) to the elective nodal clinical target volume delivered over a period of 4 weeks. Concurrent cisplatin will be administered at a dose of 50 mg/m2 weekly (capped at a maximum of 80 mg per cycle).

After completion of concurrent chemoradiation, patients in both arms will undergo brachytherapy using intracavitary or combined intracavitary-interstitial brachytherapy. The linear quadratic equivalent dose (LQED2) at 2 Gy to the high-risk clinical target volume (HR CTV) D90 should be more than $\geq$ 80 Gy depending on HRCTV volume. Alpha beta ratio for calculation of LQED2 for the high risk CTV would be 10.

Patients in both arms will undergo image guidance in the form of volumetric imaging using onboard cone-beam or megavoltage fan beam CT scans. Daily image guidance is recommended but at least 3 fractions per week should have volumetric image guidance. Similarly use of image guided brachytherapy is recommended with pre-brachytherapy MRI or MRI done with an applicator in situ. At a minimum CT guidance should be available for brachytherapy planning.

##### Pre-treatment Evaluation

All patients should have the following pretreatment evaluation for the study:

1. Clinical assessment including medical history and patient examination. The primary tumor size, vaginal involvement and parametrial extension should be noted in the records.
2. Biopsy should be obtained from the cervix for documentation of invasive cancer. Biopsy report with carcinoma in situ is not sufficient for the study. Biopsy should be obtained or reviewed at the trial center.

3. Complete hemogram: Hemoglobin, total leucocyte count, platelet count and differential leucocyte count.
4. Renal function test : Serum Urea and Creatinine
5. Electrolytes : Serum Sodium, Potassium, Magnesium and Calcium
6. Liver function test
7. HIV, HBV and HCV status
8. CEMRI of the Pelvis with standard MRI protocol
9. Contrast enhanced CT scan of Thorax and Abdomen or FDG PET CT

These investigations are performed as a standard for all new patients presenting with carcinoma cervix at the participating centers. Hence these investigations will not be funded.

#### Neoadjuvant Chemotherapy

Use of neoadjuvant chemotherapy as used in the INTERLACE trial protocol is allowed for eligible patients in both arms [5]. In case this is used the same should be specified in the process document for the institute. The regimen allowed for this will be weekly Paclitaxel (80 mg / m2) with weekly Carboplatin (2 AUC) for a maximum of six cycles. If neoadjuvant chemotherapy is used, patients should be started on radiotherapy 1 week after the last cycle of chemotherapy. Hence treatment planning should be done in the 2nd to 3rd week of chemotherapy. In all such cases, the pre-chemotherapy target volume should be used for treatment planning (specially in case of good response to vaginal or uterine disease). Response imaging is not needed after neoadjuvant chemotherapy. However a repeat planning CT may be considered at the discretion of the investigator to determine the need for adaptive replanning. Concurrent chemoradiation protocol will remain consistent. It is important that randomization is done before neoadjuvant chemotherapy is completed.

It is important to emphasize that the nodal stage should be assessed on the pre-chemotherapy imaging. Nodal status should be confirmed on pre-external beam radiotherapy imaging also.

#### External Beam Radiotherapy

##### Timing

Radiotherapy should be started within 3 weeks of randomization if neoadjuvant chemotherapy is not used.  In patients receiving neoadjuvant chemotherapy, radiotherapy should be started within 7 - 10 weeks post randomization. Concurrent chemotherapy should be initiated within 3 days of starting radiotherapy.

##### Equipment

Megavoltage equipment capable of delivering static intensity modulation with a multileaf collimator or dynamic intensity modulation (using a multileaf collimator or tomotherapy) is required. For all patients the allowed energies are 6 - 15 MV photons. For isocentric linear accelerators mounted on a C-arm gantry the Source to Source Distance (SSD) should be 100 cm.

#### Localization, Simulation and positioning

Patient preparation procedures will remain uniform and will not be dependent on the arm allocated. All patients will undergo CT simulation. Use of an immobilizing device is allowed if pre-specified in the institutional protocol. Patients can be positioned supine or prone depending on the institutional protocol, but the same positioning should be used for all patients. Patients will be scanned in the helical mode with minimum slice thickness of 2.5 mm. CT scans will be obtained from diaphragm to the mid thigh. A uniform bladder filling protocol is required to be pre-specified. Additionally centers may pre-specify any protocol used for ensuring reproducible rectal filling. Centers may choose either of the methods to account for the uterine motion:

1. A margin based approach with a uniform or differential margin is given around the uterus to account for the motion and positioning changes with bladder or rectal filling. The margin may be anisotropic but should be pre-specified in the institutional protocol.
2. An approach based on generating an ITV based on the summation of the extent of uterine motion on the full and empty bladder scans. In such a case, the empty bladder scan should be obtained after the patient has passed urine. The full bladder scan will be obtained with contrast after the patient has been instructed to drink 500 ml of water and wait for 45 minutes.

#### Target volume delineation

If the institute opts for a ITV based approach, then the full bladder and empty bladder scans will be rigidly registered with the volume of interest encompassing the pelvis. The result of the rigid registration would be visually verified to ensure that bones in the pelvis are appropriately matched. If the institute opts for a margin based approach then this step is not required.

**Primary CTV (CTVp)**: Primary CTV will encompass the gross disease and the entire cervix and uterus. The vaginal extent of the disease will determine the length of vagina to be included in the primary CTV. Typically at least 2 cm of the normal vagina beyond the clinically or radiologically demonstrated disease should be included in the CTV. CTV primary should include parametrium which will extend superiorly up to the peritoneal reflection (guided by the presence of the round ligament or the sigmoid colon). Anteriorly the boundary will be the posterior wall of the bladder or the posterior border of the external iliac vessel. Laterally it will extend to the medial edge of the internal iliac vessels. Inferiorly the volume will be bounded by the urogenital diaphragm. For institutes opting for a ITV based approach, the primary CTV will be delineated in full and empty bladder CT scans and added together to generate a composite ITV. Delineation of the primary GTV is not required for the study. Image registration with other modalities like

PET/CT and MRI to assist target volume delineation is allowed and should be followed as per the institutional process document.

**Elective Nodal CTV (CTVn_Pelvic)**:

Pelvic vessels will be delineated (both artery and vein) including the common iliac, external iliac and the visceral branches of the internal iliac vessels. The following expansions will be given around the vessels to generate the nodal CTV using the following steps:

1. 7 mm isotropic margin would be given first around the vessels.
2. The generated volume will be edited such that it is edited away from the muscles, bone, bowel and bladder.

The following table shows the CTV volume boundaries

| | |
|---|---|
| Common iliac | Superiorly: Upto the level of aortic bifurcation<br>Inferiorly: Continuous with the external and internal iliac vessels<br>Laterally: Extends to the iliopsoas laterally.<br>Posteriorly: Vertebral body. Extend contour between the psoas major and the vertebral body upto the posterior border of the vertebral body<br>Anteriorly: Trim from any bowel loops. |
| External iliac | Superiorly: Merges with the common iliac volume<br>Inferiorly : Upto the deep circumflex vessel<br>Anteriorly : Upto 10 mm from the external iliac vessel<br>Posteriorly: Extends to the obturator nodal CTV |
| Internal Iliac | Superiorly: Merges with the common iliac volume<br>Inferiorly: Upto where the vessels turn laterally to leave the pelvis.<br>Anteriorly : Merges with the obturator nodal CTV<br>Posteriorly: 7 mm from the vessel |
| Presacral | This will be drawn as a 1 cm strip in front of the vertebral body joining the internal iliac nodal CTV. Inferiorly drawn upto the slice where the piriformis muscle is visible. Sacral foramina should not be included. |
| Obturator | Drawn as a 18 mm strip joining the external and internal iliac CTV. Anterior border should extend upto the posterior border of the external iliac vein.<br>The obturator CTV should extend inferiorly upto where the obturator vessels leave the pelvis through obturator foramen. |

*Table 4: Boundaries for the different target volumes*

**Planning Target Volume**: Will be generated based on the institutional protocol. Separate PTVs will be generated for the CTVp and CTVn_Pelvic by expanding the respective CTVs by 5 mm and labelled as PTVp and PTVn_Pelvic respectively. In the HDRT arm, a planning PTV for the low dose volume can be generated by subtracting the PTVn_Pelvic from the PTVp. The planner may label the PTVn_Pelvic as Plan_PTVn_36 and Plan_PTVp_42.5 for convenience. In case of the CRT arm, these two PTV will receive the same dose. The PTV will not be trimmed from any structure unless it extends beyond the body contour in which case it will be trimmed back by 5 mm from the body surface to allow correct dose calculation.

**Critical Structures**:

The following will be delineated for all patients:

1. Femoral heads: Will be delineated separately for the left and right side. Will include the femoral head, neck, greater trochanter and the portion of femur up until the lower border of the lesser trochanter.
2. Bowel Bag: Delineated as a single structure that includes the entire external contour of the bowel.
3. Bone marrow : Pelvic, sacrum and lumbar vertebra will be delineated as a single structure. Femoral heads will not be included in the bone marrow contour.
4. Rectum: The rectum will be delineated as a single structure from the anal verge to the sigmoid colon (where the rectum turns anteriorly)
5. Bladder: Will be drawn as a single structure to include the entire bladder.

Additional organs at risk and supporting structures may be delineated as per institutional practice but the doses to the above structures are to be mandatorily reported.

#### Dose fractionation

1. Control arm (CRT) : Both the primary and elective nodal volume PTV will receive a dose of 45 Gy in 25 fractions over 5 weeks.
2. Experimental arm (HDRT): The primary PTV (PTVp) will receive a dose of 42.5 Gy in 20 fractions over 4 weeks. Simultaneously, the PTVn_Pelvis will receive a dose of 36 Gy in 20 fractions over the same period of time.

Twice daily treatments are not allowed unless mandated to compensate for unplanned gaps in treatment. Breaks or planned gaps in treatment are not allowed except to allow for interdigitating brachytherapy applications.

#### Dose Specification

No point based dose prescription is needed though a reference point may be generated for the planning process. No plan normalization is allowed and dose should be prescribed to the 100% isodose. The following will be the dose specification for the volumes:

| | Control (CRT) | Experimental (HDRT) |
|---|---|---|
| PTVp | 45 Gy in 25 fractions (1.8 Gy)<br>EQD2 = 44.25 $Gy_{10}$ / 43.2 $Gy_3$ | 42.5 Gy in 20 fractions (2.125 Gy)<br>EQD2 = 42.94 $Gy_{10}$ / 43.56 $Gy_3$ |
| PTVn_Pelvis | 45 Gy in 25 fractions (1.8 Gy) | 36 Gy in 20 fractions (1.8 Gy) |

| | EQD2 = 44.25 $Gy_{10}$ / 43.2 $Gy_3$ | EQD2 = 34.56 $Gy_{10}$ / 35.4 $Gy_3$ |
|---|---|---|

*Table 5: Dose prescription and corresponding linear quadratic equivalent doses for the different volumes.*

The dose schedule has been explicitly chosen to ensure that the LQED2 to the primary clinical target volume (CTVp) is isoeffective. The minimal difference in the EQD2 can be made using brachytherapy. For the elective pelvic nodal clinical target volume the LQED2 represents a 20% dose reduction in the HDRT arm as compared to the dose in the CRT arm.

#### Treatment Planning

As inverse planned intensity modulated radiotherapy is mandated in this protocol, the field arrangements, collimator alignment and other beam parameters will be determined by the planning approach used in each institute. Use of non-coplanar fields is allowed. However if field junctions are needed for adequate coverage the junctional dose should be measured in at least 5 patients as a part of the quality assurance process. Both volumetric arc and fixed field approaches for intensity modulation are allowed. However, each institute should specify the planning technique to be adopted in the process document for the study. Irrespective of the planning technique used, the following dose constraints should be met as planning objectives. Physicists are free to create dummy structures to support plan optimization. Additionally, MU objectives and Normal Tissue Avoidance objectives available in the treatment planning systems can be used as long as the planning strategy is pre-specified. Institutes will be encouraged to designate machines for which treatment planning can be done for this trial. As this will be a volumetric prescription, the ICRU 84 recommendations for dose reporting should be followed.

| **Volume** | **Criteria** | **Control (CRT)** | | **Experimental (HDRT)** | | |
|---|---|---|---|---|---|---|
| | | Mandatory | Optimal | **Criteria** | Mandatory | Optimal |
| PTVp | **D98** | ≥ 40.5 | ≥ 42.75 | **D98** | ≥ 38.25 | ≥ 40.38 |
| | **D95** | **≥ 42.75** | **≥ 44.1** | **D95** | **≥ 40.38** | **≥ 41.65** |
| | **D2** | ≤ 48.15 | ≤ 47.25 | **D2** | ≤ 45.46 | ≤ 44.62 |
| PTVn_Pelvis | **D98** | ≥ 40.5 | ≥ 42.75 | **D98** | ≥ 32.4 | ≥ 34.2 |
| | **D95** | ≥ 42.75 | ≥ 44.1 | **D95** | ≥ 34.2 | ≥ 35.28 |
| | **D2** | ≤ 48.15 | ≤ 47.25 | **D2*** | ≤ 38.5 | ≤ 37.8 |
| Bladder | **Dmax** | ≤ 47.25 | ≤ 45 | **Dmax** | ≤ 44.62 | ≤ 43.78 |
| | **V40** | NA | ≤ 70% | **V37** | NA | ≤ 70% |
| Rectum | **Dmax** | ≤ 47.25 | ≤ 45 | **Dmax** | ≤ 44.62 | ≤ 43.78 |

| | | | | | | |
|---|---|---|---|---|---|---|
| | **V40** | NA | ≤ 85% | **V37** | NA | ≤ 85% |
| Femoral Head | **D2** | ≤ 47.25 | ≤ 45 | **D2** | ≤ 44.62 | ≤ 43.78 |
| | **V30** | ≤ 15% | ≤ 10% | **V24** | ≤ 15% | ≤ 10% |
| Bowel Bag | **V40** | NA | ≤ 100 | **V32** | NA | ≤ 100 |
| | **V30** | NA | ≤ 350 | **V24** | NA | ≤ 350 |
| | **Dmax** | ≤ 47.25 | ≤ 45 | **Dmax** | ≤ 44.62 | ≤ 43.78 |
| Duodenum | **D2cc** | ≤ 45 | NA | **D2cc** | ≤ 38.25 | NA |
| Spinal Canal | **Dmax** | ≤ 45 | NA | **Dmax** | ≤ 38.25 | NA |
| Kidney | **V12** | ≤ 55% | ≤ 40% | **V11** | ≤ 55% | ≤ 40% |
| | **V20** | ≤ 10% | ≤ 8% | **V18** | ≤ 10% | ≤ 8% |
| | **V23** | ≤ 5% | ≤ 3% | **V21** | ≤ 5% | ≤ 3% |
| | **V28** | ≤ 3% | ≤ 1% | **V26** | ≤ 3% | ≤ 1% |
| PRV Ovary* | **Dmean** | ≤ 5 | ≤ 3 | **Dmean** | ≤ 5 | ≤ 3 |
| | **V5.5** | ≤ 29% | ≤ 10% | **V5** | ≤ 29% | ≤ 10% |
| | **V10** | ≤ 20% | ≤ 5% | **V9** | ≤ 20% | ≤ 5% |

*Table 6: Showing the dose constraints to be followed for the trial. Mandatory constraints should be met in all cases. * D2 for PTVn_Pelvis for the HDRT arm to be evaluated for a planning volume which is not overlapping the PTVp with an additional margin of 3 mm. ** PRV Ovaries are required only if oophoropexy has been performed. Vx =Volume receiving x dose in Gy, Dx = Minimum dose to x volume. For CRT arm plan evaluation for the PTVp and PTVn_Pelvis can be done as a singular structure.*

#### Beam Energy

Most patients will be planned with 6 MV photons. Use of flattening filter free 6 MV and 10 MV photons is allowed if adequately commissioned for use with IMRT.

#### Plan Quality Assurance

As a trial which mandates Intensity Modulated Radiotherapy, per patient plan quality assurance is necessary. For all patients, a verification plan will be created on a phantom. Point dose verification will be done at isocenter with the help of an ion chamber. Variation within 3% of the prescribed dose will be considered acceptable. Additional fluence verification with the help of additional ion chambers at other points, ion chamber / diode arrays and films are allowed. The plan quality assurance parameters including the type of phantom and dose measurement system should be detailed in the process document. In addition, the parameters followed for generating the plan for quality assurance should be specified. Every effort must be made to replicate the delivery conditions in the phantom plan.

##### Treatment Delivery

Radiotherapy will be delivered using megavoltage equipment on 5 days a week treating one fraction per day. All fields should be treated on all days.

##### Verification Imaging

Verification imaging before daily treatment will be obtained  with daily volumetric cone beam KV CT or MV fan beam CT. In case of a fault in the volumetric imaging system, EPID based matching is allowed if the patient cannot be shifted to another unit with a functional imaging system. Ideally no more than two fractions in a week should be done using planar imaging.

##### On treatment review

On treatment, reviews will be performed weekly for all patients. During each review visit, patients would be examined clinically to document any toxicities especially cutaneous and mucosal toxicities. Toxicity grading will be done using the Common Terminology Criteria of Adverse Events (CTCAE) version 6.0. More frequent reviews may be done in patients who have grade 3 or higher toxicities.

##### Treatment scheduling and gaps

Treatments can start Monday through Wednesday. However it is preferred to start treatment on Monday or Tuesday. Gaps in treatment should be avoided. In case the institute has beam matched machines, the patient should be shifted to the other machine to avoid treatment interruptions. In case beam matched machines are not available or not serviceable, efforts should be made to treat the patient in another machine without interruptions. It is allowed to use 3DCRT for this purpose to avoid unnecessary treatment interruptions. However, efforts should be made to get an IMRT plan ready for this machine within 3 working days. Any such plan should also undergo quality assurance before delivery.

#### Brachytherapy

All patients included in the study will receive intracavitary brachytherapy or interstitial -intracavitary implant after completion of external radiation depending upon the response to concurrent chemoradiation. All brachytherapy will be executed using CT-based planning. The preferred dose regime for intracavitary brachytherapy will be 7Gy x 4 sessions prescribed to the HRCTV. However, institutes are free to individualize the schedule of brachytherapy to achieve the planned dose targets..The minimum dose to the high-risk clinical target volume(HR CTV) D90 should be 80-90 Gy depending on HRCTV volume. Brachytherapy dose constraints will remain same in both arms and outlined in the table 7 below.

| Volume | Paramete r | Optimal | Mandatory |
|---|---|---|---|
| High Risk Clinical Target Volume (HRCTV) | D90 | $\geq$ 85 $Gy_{10}$ | $\geq$ 80 $Gy_{10}$ |
| | D98 | $\geq$ 80 $Gy_{10}$ | $\geq$ 75 $Gy_{10}$ |
| Rectum | D2cc | $\leq$ 65 $Gy_3$ | $\leq$ 70 $Gy_3$ |
| Sigmoid | D2cc | $\leq$ 70 $Gy_3$ | $\leq$ 75 $Gy_3$ |
| Bladder | D2cc | $\leq$ 80 $Gy_3$ | $\leq$ 85$Gy_3$ |
| Bowel loops | D2cc | $\leq$ 70 $Gy_3$ | $\leq$ 75 $Gy_3$ |

*Table 7: Cumulative dose constraints for Brachytherapy to be followed in both arms expressed in EQD2. Note that sigmoid and bowel loop cumulative doses to be calculated with respect to the full external beam radiotherapy dose even though the actual dose may be lower. Alpha beta ratio of 10 to be considered to target volume and 3 for organs at risk. D90 = Minimum dose to 90% volume, D98 = Minimum dose to 98% volume, D2cc = Minimum dose to most exposed 2 cc volume*

#### Concurrent Chemotherapy

For those with creatinine clearance >50 ml/min, cisplatin will be administered at a dose of 40 mg/m$^2$ weekly during external beam radiation in the control arm (CRT) and 50 mg/m$^2$ in the experimental arm (HDRT).The maximum dose in a week for the control and experimental arms are capped at 70 mg and 80 mg respectively. In case of creatinine clearance between 40 and 50 ml/min, a 25% dose reduction will be applied. Patients with creatinine clearance below 40 will be treated with radiation alone. Assessment of calculated creatinine clearance will be done prior to each cycle of chemotherapy. Substitution of Cisplatin with carboplatin is not allowed in this trial.

#### Overall treatment time

Overall treatment time will be calculated from the start of EBRT till the last fraction of brachytherapy. This calculation will only apply to patients who have completed their treatment. For patients where the treatment was incomplete or where brachytherapy had to be replaced with supplementary external pelvic radiotherapy, the overall treatment time will be calculated and reported separately. The ideal overall treatment time for the control and experimental arms are 56 and 49 days respectively.

#### Treatment therapy Adverse events

#### Expected

Nearly all participants in the study will experience expected adverse events.

1. Tiredness
2. Fatigue
3. Loss of appetite
4. Permanent amenorrhea if pre-menopausal

#### Common

Common adverse events will be experienced by nearly 10 - 50% of the participants in the study

1. Bowel Symptoms:
    a. Nausea and vomiting
    b. Diarrhea
    c. Frequency of motions
    d. Urgency of motions
    e. Urge incontinence of stools
    f. Pain in abdomen
    g. Bleeding in stools
    h. Passage of mucus in stools
    i. Pain during passage of stools
    j. Anal stenosis
2. Urinary Symptoms:
    a. Frequency of urination
    b. Urgency of urination
    c. Blood in urine (hematuria)
3. Redness or blistering in the skin
4. Hematological side effects:
    a. Anemia
    b. Leucopenia
    c. Thrombocytopenia
    d. Neutropenia
5. Electrolyte imbalances:
    a. Hyponatremia
    b. Hypomagnesemia
    c. Hypocalcemia
6. Endocrine and sexual
    a. Hot flushes
    b. Vaginal stenosis and shortening
    c. Vaginal dryness
    d. Dyspareunia

##### Less Common

Less common side effects will be experienced by 10% or less participants in the study:

1. Hematological:
   a. Febrile neutropenia
2. Vagina:
   a. Vaginal necrosis
3. Bowel:
   a. Obstruction
   b. Perforation
   c. Fistula formation
4. Bladder:
   a. Bladder contracture
   b. Fistula formation
   c. Urethral stricture
5. Bone:
   a. Avascular necrosis of femur
   b. Pelvic insufficiency fractures
   c. Osteoporosis
6. Renal:
   a. Renal injury
7. Hearing
   a. Sensorineural hearing loss
8. Neurological
   a. Sensory neuropathy
   b. Motor neuropathy

#### Treatment Modifications

##### External Beam Radiotherapy

Treatment should not be interrupted unless the patient has particularly severe Grade IV toxicity. In particular external beam radiotherapy treatment should not be interrupted for thrombocytopenia unless counts go below 20,000. In such cases a daily manual platelet count may be mandated before treatment. Use of platelet transfusions is allowed to maintain the radiation dose intensity. Similarly radiation should not be held for neutropenia and use of growth factor support is allowed. Radiation can be interrupted for severe diarrhea, vomiting and other life threatening toxicities but should be resumed as soon as it is clinically safe to do so. In the event radiation is interrupted, external beam radiotherapy should not be used for gap correction. Instead dose escalation should be done using brachytherapy if required.

#### Concurrent Chemotherapy

Carboplatin should not be substituted for Cisplatin in the event of toxicity. The following modifications are recommended for concurrent cisplatin.

**Haematological Toxicity:**

| Parameter & Value | Cisplatin |
|---|---|
| ANC < 1000 | Withhold Cisplatin. Repeat ANC after 1 week.<br>If ANC not recovered in 2 weeks then omit cisplatin<br>Else 80% dose. |
| ANC < 500 or Febrile Neutropenia | Withhold Cisplatin. If ANC does not recover > 500 stop Cisplatin.<br>If febrile neutropenia Cisplatin should be restarted only if ANC > 1000 at 80% dose reduction |
| Platelet < 75,000 (manual) | Omit treatment. Future treatment at 80% dose. |

Discontinue chemotherapy if there is a repeat occurrence of febrile neutropenia or chemotherapy omission low ANC and / or platelet count. Proceed with radiation only

**Renal Toxicity:**

| Parameter & Value | Cisplatin |
|---|---|
| - Creatinine clearance 60 - 50 ml/min | 80% of the dose |
| - < 50 ml/min | Withhold cisplatin |

Stop Cisplatin if it has to be omitted for 2 cycles. Repeat Creatinine clearance

**Gastrointestinal Toxicity:**

Patients with Grade 3 or 4 nausea / vomiting / diarrhoea / anorexia / constipation should have chemotherapy held till toxicity settles to Grade I or less. Subsequent cycles can be administered after discussing with a consultant. Dose of Cisplatin to be modified to 80% Discontinue chemotherapy if there are more than one such episodes or dose reduction needed for an additional toxicity.

**Special situations:**

In the event that the patient has multiple toxicities which merit a dose reduction or dose modification, the lowest dose should be used. If chemotherapy omission is required for multiple toxicities , all toxicities should resolve to Grade 1 or less before chemotherapy is restarted. If chemotherapy cannot be given for two consecutive cycles then it should be stopped and radiation continued. All such cases should be discussed with the consultant. Stop concurrent chemotherapy if there is hypersensitivity.

#### Brachytherapy

Brachytherapy should be administered as soon as possible after the conclusion of external beam radiotherapy. Interdigitation of brachytherapy is allowed but if adopted then the patient should not receive external beam radiotherapy or concurrent chemotherapy on the same day.
In the unlikely event that the patient is unfit for brachytherapy or refuses to undergo brachytherapy, the preferred alternative treatment is surgery (total hysterectomy). If surgery is not feasible then additional external beam radiotherapy boost can be considered at the investigator's discretion. In case this modality is chosen the total dose delivered should be restricted to the primary disease only and not the entire pelvis. Hypofractionated radiotherapy should not be used for the external beam radiotherapy boost. Consideration should be given to daily image guided volumetric modulated arc therapy to reduce organ at risk dose.
All cases where brachytherapy is not possible would be considered as protocol violations.

#### Concomitant Therapy

Concomitant medications like antiemetics, growth factors, analgesics, antidiarrheals and other medications can be given as per standard practice during the treatment. The use of investigational drugs is not allowed unless being done as a part of a dedicated clinical trial. The patient must be told to notify the treating physician about any new medications that he/she takes after the start of treatment. Any drug used concomitantly should be assessed for any potential interactions with the chemotherapy by the investigator team.

### Outcome

#### Trial Run In

For the run in phase of the trial key outcome of interest is completion of the treatment in the HDRT arm which will be considered to be protocol compliant if all of the following four criteria are met:

1. Mandatory dose constraints are achieved.
2. EBRT duration is within 31 days (from start of EBRT to end of EBRT). The extra 3 days is to account for logistical issues stemming from machine breakdown and does not include breaks due to toxicity.
3. Brachytherapy delivered such that an overall treatment time of 49 days is maintained.
4. The D90 to HRCTV is over 80 Gy.
5. At least 3 cycles of concurrent chemotherapy were delivered.

As defined before, the trial protocol would be considered deliverable if 16 of the 20 patients complete the treatment as defined above.

#### Primary Outcome

The primary outcome of interest is the incidence of acute toxicity defined using the CTCAE 6.0 grading system. Acute toxicity is defined as an adverse effect occurring during the course of radiotherapy and upto 6 weeks after completion of treatment. The maximum grade of each toxicity endpoint the patient experiences will be evaluated. The following the key gastrointestinal physician defined toxicity endpoints will be evaluable for this outcome (See appendix II for the definition of grades): diarrhea, vomiting, nausea and abdominal pain. Proportion of patients experiencing any grade 2 or higher gastrointestinal will be compared between the two arms.

#### Secondary Outcomes

1. **Disease free survival**: This will be defined as the interval between the date of randomization to the documented date of death, any recurrence or a second malignancy (whichever event is detected earliest). Patients who are alive without disease will be censored at the last date of follow up. The actuarial estimate of disease free survival in both arms will be estimated along with 95% confidence intervals.
2. **Patient reported outcome**: Patient reported outcomes will be evaluated using the EORTC QLQ C30 and CX24 questionnaires which will be administered at baseline and subsequently at each week during radiotherapy and at each scheduled follow up visit. The three scales of interest for this endpoint are the global quality of life, C30 summary score (defined by Glessinger et al) and the symptom experience score. These will be calculated from responses using guidance provided by the EORTC group. Mean scores and 95% confidence intervals of mean will be compared between groups at 6 months, 12 months, 24 months and 36 months.
3. **Late Toxicity**: The cumulative incidence and 95% confidence intervals of each of the defined late toxicity endpoints (see Appendix II) will be compared between the two arms.

#### Translational Outcomes

1. **Circulating cell free DNA**: Compare the circulating human papilloma virus circulating cell free DNA (cfDNA) levels between the two arms. A quantitative estimate obtained from the test mean copies per microlitre will be reported. The cfDNA levels will be quantified using digital droplet polymerase chain reaction at the following time points:
    a. Before start of radiotherapy (Baseline)
    b. Completion of EBRT (end of CtRT)
    c. At 3 months post treatment (Post treatment)
    d. At 6 months post treatment (Post treatment)
    e. At 9 months post treatment (Post treatment)
2. **Lymphocyte Counts**: Absolute lymphocyte counts will be obtained at different time points coinciding with the specimen acquisition for cfDNA estimation in addition to the time points at which complete hemogram is routinely performed during treatment. The mean values at each of these time points will be compared to compare the nadir values of the lymphocyte counts between each arm, and the total time required for lymphocyte count recovery to CTCAE 6.0 Grade I or better ( > 800 /$mm^3$). Total time for recovery

will be computed using actuarial method and overall duration of time with 95% confidence intervals for each arm will be reported.

#### Participant Timeline

The following table shows the participant timelines at randomization. Baseline screening and eligibility evaluation will be performed before starting chemoradiation.

| Assessment | Arm | Rand | EBRT (week) | | | | | BRT | Followup (month) | | | | | | | | | | | | | |
|---|---|---|---|---|---|---|---|---|---|---|---|---|---|---|---|---|---|---|---|---|---|---|
| | | | 1 | 2 | 3 | 4 | 5 | | 3 | 6 | 9 | 12 | 15 | 18 | 21 | 24 | 30 | 36 | 42 | 48 | 54 | 60 |
| History | Both | X | | | | | | | X | X | X | X | X | X | X | X | X | X | X | X | X | X |
| Physical Exam | Both | X | | | | | | | X | X | X | X | X | X | X | X | X | X | X | X | X | X |
| EBRT | CRT | | X | X | X | X | X | | | | | | | | | | | | | | | |
| | HDRT | | X | X | X | X | | | | | | | | | | | | | | | | |
| Chemotherapy | CRT | | X | X | X | X | X | | | | | | | | | | | | | | | |
| | HDRT | | X | X | X | X | | | | | | | | | | | | | | | | |
| Brachytherapy | Both | | | | | | | X | | | | | | | | | | | | | | |
| Toxicity | CRT | X | X | X | X | X | X | | X | X | X | X | X | X | X | X | X | X | X | X | X | X |
| | HDRT | X | X | X | X | X | | | X | X | X | X | X | X | X | X | X | X | X | X | X | X |
| Quality of Life (C30 & CX24) | CRT | X | X | X | X | X | X | | X | X | X | X | X | X | X | X | X | X | X | X | X | X |
| | HDRT | X | X | X | X | X | | | X | X | X | X | X | X | X | X | X | X | X | X | X | X |
| CBC + DIFF | CRT | X | X | X | X | X | X | | X | X | X | X | | | | | | | | | | |
| | HDRT | X | X | X | X | X | | | X | X | X | X | | | | | | | | | | |
| Elec/Renal | CRT | X | X | X | X | X | X | | | | | | | | | | | | | | | |
| | HDRT | X | X | X | X | X | | | | | | | | | | | | | | | | |
| Plasma Biospecimen* | CRT | X | | | | | X | | X | X | X | X | | | | | | | | | | |
| | HDRT | X | | | | X | | | X | X | X | X | | | | | | | | | | |
| Tissue Biopsy* | CRT | X | | | | | | X | | | | | | | | | | | | | | |
| | HDRT | X | | | | | | X | | | | | | | | | | | | | | |

*Table 8: Schedule of Assessments in each arm. * Plasma and tissue biospecimen collection in patients who consent for biobanking only. CRT = Conventional Radiotherapy, HDRT = Hypofractionated Dose Painted Radiotherapy, EBRT = External Beam Radiotherapy, C30 = EORTC QLQ C30 , CX24 = EORTC CX24, CBC+DIFF = Complete hemogram with differential counts, Elec/Renal = Electrolytes and renal panel (Serum sodium, potassium, calcium, magnesium, creatinine), BRT = Brachytherapy. Note that chemotherapy will be delivered with concurrent cisplatin at a dose 40 mg/m2 weekly in the CRT arm and 50 mg/m2 in the HDRT arm.*

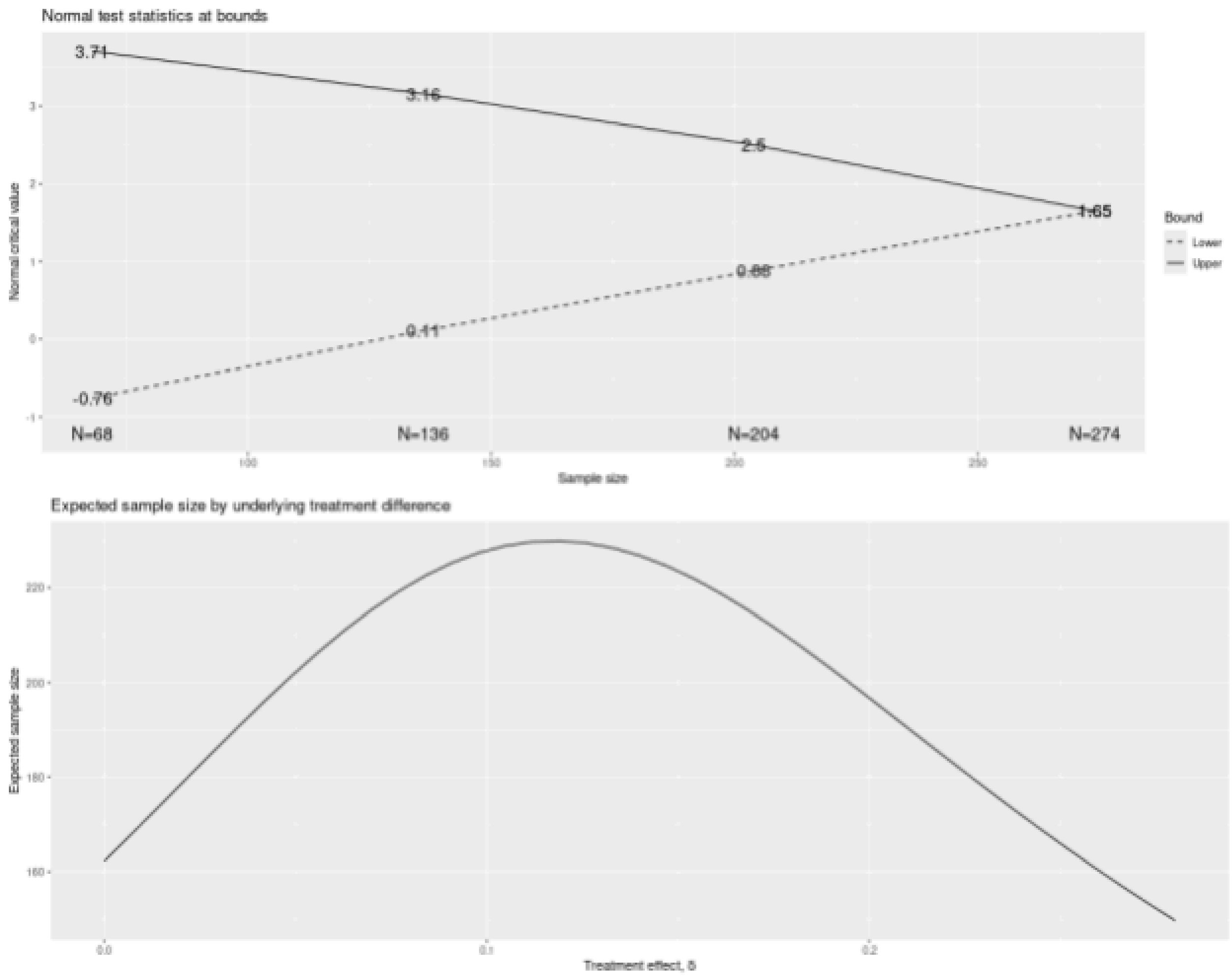


*Figure 2: Plots for Normal test statistics at bounds at each interim analyses (top) and the expected sample size for different treatment effects (absolute difference in toxicity).*

The tabular summary of the interim analyses with Z values for efficacy and futility bounds for each interim analysis are described in the table below. The final sample size is rounded to 300 patients to account for dropouts.

| Analysis | Value | Efficacy | Futility |
|---|---|---|---|
| IA 1: 25% | Z | 3.7057 | -0.7603 |
| N: 68 | p (1-sided) | 0.0001 | 0.7765 |
| | ~risk difference at bound | 0.4019 | -0.0825 |
| | P(Cross) if risk difference=0 | 0.0001 | 0.2235 |
| | P(Cross) if risk difference=0.14 | 0.0079 | 0.0201 |

| Analysis | Value | Efficacy | Futility |
|---|---|---|---|
| IA 2: 50% | Z | 3.1560 | 0.1085 |
| N: 136 | p (1-sided) | 0.0008 | 0.4568 |
| | ~risk difference at bound | 0.2420 | 0.0083 |
| | P(Cross) if risk difference=0 | 0.0009 | 0.5626 |
| | P(Cross) if risk difference=0.14 | 0.0933 | 0.0532 |
| IA 3: 74% | Z | 2.5033 | 0.8820 |
| N: 204 | p (1-sided) | 0.0062 | 0.1889 |
| | ~risk difference at bound | 0.1567 | 0.0552 |
| | P(Cross) if risk difference=0 | 0.0064 | 0.8255 |
| | P(Cross) if risk difference=0.14 | 0.3973 | 0.1075 |
| Final | Z | 1.6508 | 1.6508 |
| N: 274 | p (1-sided) | 0.0494 | 0.0494 |
| | ~risk difference at bound | 0.0892 | 0.0892 |
| | P(Cross) if risk difference=0 | 0.0454 | 0.9546 |
| | P(Cross) if risk difference=0.14 | 0.8020 | 0.1980 |

*Table 8: IA is the percent of statistical information (planned sample size) at interim analysis. N = rounded sample size. The nominal p-values indicative of superiority and futility at each time point are shown in the table. The p(Cross) refers to the cumulative probability of cross a bound at or before the given analysis under the null hypothesis*

The total number of cervical cancer patients registered in the three participating centers annually is 400. Assuming that 50% of the patients will be eligible, and about 50% will give consent, we estimate that 100 patients will be randomized each year. The total duration of accrual will therefore be 3 years. Assuming that the initial accrual with quality assurance will be slower we expect that the first interim analysis will be conducted at the end of year one. Subsequent interim analyses will be planned at the 2nd, 3rd and 4th year (spaced approximately 8 - 9 months apart, depending on the accrual speed).

#### Secondary Outcome

The key secondary outcome of interest is disease free survival. Given the low incidence of pelvic nodal relapse in patients with node negative cervical cancer, a clinically meaningful threshold of non-inferiority is difficult to test. In the UPGRADE-RT trial reported by Bosch et al, the primary outcome measure was normalcy in the diet score while the secondary outcome measure was 2-year recurrence in the elective nodal volume [11]. The authors expected a rate of 5% in the control arm and chose an upper bound of acceptability of 9% with the justification that ENI can be avoided when the risk of nodal failure is less than 10 - 15% in head neck cancers. In the trial by Nuyts et al, the primary outcome of interest was dysphagia at 6 months [11,12]. No sample size calculations were performed for determination of the tumor control.

If we assume that the disease free survival in the control arm is 70% at 3 years and assume a non-inferiority margin corresponding to a hazard ratio of 1.5, a sample size of 300 participants provides us with a power of 0.73 to determine a non-inferiority with a one sided type I error of 5%. Additionally, assuming that the proportion of patients with a failure in the elective nodal region is 5%, sample size of 150 participants in the experimental arm will allow us to determine this proportion of patients with a failure in the elective nodal region with an absolute margin of error of 3.46% with 95% confidence.

### Recruitment

Patients will be recruited from the outpatients presenting in the disease management group outpatient departments of the recruiting institute. Patients will be randomized after evaluation. No advertising is permitted for recruitment and no inducements will be given for recruitment of patients in the study. Local principal investigators are however free to use methods as required to improve accrual to the current study.

### Randomization

#### Sequence Generation

The randomization sequence will be generated by an independent statistician who will then put the sequence in the REDCap database. Once added the sequence cannot be changed or modified. The randomisation schedule will be created using computer-generated random numbers before the first participant has been recruited, in a one-to-one ratio. Randomization sequence will be generated and stored in the REDCap randomization module for random patient assignment.

#### Screening

All diagnostic tests should have been done within 28 days of the screening for the study. These include:

1. History and physical examination
2. Complete hemogram, renal function
3. Serum electrolytes including calcium and magnesium
4. Viral serology
5. Liver function test
6. MRI Pelvis
7. FDG PET CT

These tests are done routinely for all patients in the participating centers, and will not be funded through the study.

#### Baseline Assessments

All patients will undergo routine baseline investigations including appropriate clinical review, and baseline pathology review as outlined subsequently. No additional imaging investigation is required for the patients participating in the trial unless the investigator feels it is clinically indicated. In consenting patients, (optional) baseline biopsy samples and a 10 mL sample of whole blood and plasma will be stored at TMC Biobank for future translational studies.Clinical examination is needed to document the performance status and the local disease status before radiotherapy.

#### On treatment Assessment

Patients will be followed weekly during adjuvant radiation  with routine clinical review and toxicity grading (CTCAE 6.0).

#### Quality of Life Assessment

Quality of life assessments will be performed for patients enrolled in the quality of life substudy using the European Organization for Research and Treatment of Cancer (EORTC) quality of life questionnaire 30 (QLQ C30) and the Cervix 24 (EORTC CX 24) questionnaire. Regional language translations will be made available so that patients can self-report quality of life assessments.  Assessments will be performed at baseline and subsequently weekly during external beam radiotherapy. After radiotherapy completion, quality of life assessments will be performed at each follow up visit. Additional assessments at interim follow up visits may also be conducted. Paper and electronic administration is allowed. Additionally, quality of life data can be collected through telephonic interviews if the patient follow-up cannot be maintained physically.

##### Follow up Assessment

Patients will then be reviewed 3 monthly for 2 years, and then 6 monthly till 5 years. After 5 years an annual follow up will be recommended. Each patient is planned to complete the study treatment unless disease progression occurs or toxicity (CTCAE 6.0) prohibits further therapy. At each follow up patient will undergo a clinical evaluation to document the disease status and document late toxicities. Imaging is not required in these patients but symptom directed imaging can be ordered as required. The patient follow up schedule will be maintained in the REDCap database and patients will be contacted at regular intervals to ensure adherence to follow up visits.In case the patient is unable to come for a physical follow up telemedicine follow-ups are allowed.

##### End of Study Visit

The end of study visit will be completed in case of patient death, withdrawal of consent or at 3 years after the last patient is enrolled in the trial.

##### Post Study Closure

The study will be closed after the required number of events have been observed. Collection of long-term outcomes, for example, survival data, will continue after the main study is closed, using a simple follow-up CRF, monitoring of central registries, contact with treating clinicians, etc

##### Consent withdrawal

If the patient desires to withdraw consent then the same should be documented clearly in the REDCap database. After the withdrawal of the consent, no further data collection is permitted for the patient. However if the patient has been randomized prior to withdrawal of the consent, then data collected till that time will be retained.

#### Data Management

Access to the data will be available to the principal investigator of the study as well as to the co-investigators. The data quality and integrity will be checked at quarterly intervals using the data quality checking system available in Redcap. Patient identifiers will be noted as such in the Redcap database and the same will be used for ensuring that any data exports contain de-identified data only. Institutes will have access to the data of their own institute during the trial phase. Once the trial is completed the main database will be checked for data consistency and quality and then closed for analysis.

Source documents pertaining to the trial must be maintained by investigational sites. Source documents may include a subject's medical records, hospital charts, clinic charts, the investigator's subject study files, as well as the results of diagnostic tests such as X-rays, laboratory tests, and electrocardiograms. The investigator's copy of the case report forms serves as part of the investigator's record of a subject's study-related data. All study-related documentation will be maintained for 10 years following completion of the study or according to existing regulatory requirements.

The following information should be entered into the subject's medical record:

1. Subject’s name, contact information and protocol identification.
2. The date that the subject entered the study, and the subject number.
3. A statement that informed consent was obtained (including the date).
4. Relevant medical history
5. Dates of all subject visits and results of key trial parameters.
6. Occurrence and status of any adverse events.
7. The date the subject exited the study, and a notation as to whether the subject completed the study or reason for discontinuation

##### Additional Analyses

Additional subgroup analyses for the primary and secondary endpoints have not been planned as a part of this study. However if any are planned these will be included as a future amendments to the statistical analysis plan.

#### Data Monitoring

An independent data safety and monitoring committee (DSMC) will be set up. Members of this DSMC will review the overall conduct of the study. Trial safety will be assessed by evaluation of the severe acute toxicity rate and rates of treatment interruption in the first 50 patients enrolled in the trial. The trial monitoring committee will independently evaluate these parameters to determine continuation of the trial. The DSMC will be composed of radiation oncologists and physicists who will also review the accumulated dosimetry of at least 15 representative plans in the experimental arm to determine adherence to the proposed dose constraints.

### Safety Reporting

#### Assessment of Safety

##### Definitions

###### Adverse Events

An ADVERSE EVENT (AE) is any untoward medical occurrence in a patient or clinical investigational subject administered a pharmaceutical product and which does not necessarily have a causal relationship with this treatment. An AE can therefore be any unfavourable or unintended sign (including an abnormal laboratory finding), symptom, or disease temporally associated with the use of a medicinal investigational product, whether or not considered related to the medicinal product (see below).

Adverse events include the following:

1. All suspected adverse reactions from radiotherapy
2. All reactions from concurrent chemotherapy or medications given as concomitant medications with concurrent chemotherapy
3. Apparently unrelated illnesses, including the worsening (severity, frequency) of pre-existing illnesses
4. Injury or accidents.
5. Abnormalities in physiological testing or physical examination that require clinical intervention or further investigation (beyond ordering a repeat examination)
6. Laboratory abnormalities that require clinical intervention or further investigation (beyond ordering a laboratory test).
7. Any untoward event that occurs after the protocol-specified reporting period which the Investigator believes may be related to the radiotherapy

AEs must be reported as AEs even if they do not meet SAE criteria.

###### Serious Adverse Events

Any adverse experience occurring at any dose that results in any of the following outcomes:

- Death;
- A life-threatening adverse experience;
- Inpatient hospitalization or prolongation of existing hospitalization;
- A persistent or significant disability/incapacity;
- A congenital anomaly/birth defect.

Important medical events that do not result in death, are not life-threatening, or do not require hospitalization may be considered an SAE experience, when, based upon medical judgment, they may jeopardize the patient and may require medical or surgical intervention to prevent one of the outcomes listed in the definition.

SAEs (more than 3 months after last treatment) attributed to the protocol treatment (possible, probable, or definite) will also be reported.

Note: All deaths in the study require both routine and expedited reporting regardless of causality. Attribution to treatment or other causes must be provided. “On study” is defined as during or within 3 months of completing protocol treatment.

**Suspected Unexpected Serious Adverse Reaction (SUSAR)**

A SUSAR is an SAE that is related to the drug or device and is unexpected (i.e. not listed in the investigator brochure or approved Product Information; or is not listed at the specificity or severity that has been observed; or is not consistent with the risk information described in the Subject Information Sheet and Informed Consent Form or elsewhere in the protocol. (FDA, Safety Reporting Requirements for INDs and BA/BE Studies, draft guidance, September 2010)).

An event is causally related if there is a reasonable possibility that the drug [intervention] caused the AE, i.e. there is evidence to suggest a causal relationship between the drug and the event (FDA, Safety Reporting Requirements for INDs and BA/BE Studies, draft guidance, September 2010).

The following events will not be reported as SAEs:

1. Hospitalization to deliver study procedures e.g. brachytherapy or external beam radiotherapy or chemotherapy
2. Hospitalization to treat disease progression
3. Unrelated scheduled elective surgery
4. Convenience purposes e.g. transportation issues / blood transfusions.

**Reporting SAE**

Adverse Events (AEs) and Serious Adverse Events (SAEs) that meet the criteria defined above experienced by patients accrued to this protocol will be reported within 24 hours to the local institutional board as per the standard guidelines. Attribution of the relatedness will be determined by the institutional review board. All SAEs occurring at other sites will also be reported to the institutional IRB of Tata Medical Center with 24 hours of their reporting to Tata Medical Center.

#### Auditing

The study may be subject to audit and inspection by the representatives of the regulatory bodies which may include hospital level Institutional review board, stage regulatory bodies and national regulatory bodies.

#### Declaration of Interests

The investigators have no conflicts of interest to declare.

#### Access to Data

Intra study data sharing will be done with input from the Trial management committee. All site PIs will be provided with access to the cleaned dataset. All project data will be housed in the institutional REDCap installation available at Tata Medical Center. Data access groups will be used to restrict access to the institutional data to individual institutions. Minimum identifying personal information will be stored.

#### Ancillary and Post Trial Care

Patients will continue to receive post trial care as per the institutional protocols in place. Ancillary care for late toxicity related outcomes will also be provided as per the standard institutional protocol. Patients enrolled will be covered by indemnity for negligent harm through an insurance policy for the trial. Additional coverage for additional healthcare, compensation and damages will also be covered through the same insurance policy.

#### Dissemination Policy

The Trial Management Committee will appoint a Writing Committee to draft manuscript(s) based on the trial data. Manuscript(s) will be submitted to peer-reviewed journal(s). The Writing Committee will develop a publication plan, including authorship, target journals and expected dates of publication. All publications must be circulated to the trial management committee and receive approval prior to submission. Note that for the purpose of approval an email approval is recommended but not mandatory. Additionally trial results will be reported at the clinical trial registry of India as per the extant policies and regulations.

All publications and presentations will be submitted on behalf of an author group. A trial author group composed of all members of the trial management committee as well as any additional contributors will have oversight over each publication. All members in the author group will be considered to have equal authorship in publication and names in the author group will be kept in the alphabetical order using the first name and last name. One of the authors will be the designated corresponding author.

#### Clinical Study Report

A Clinical Study Report which summarises and interprets all the pertinent study data collected will be issued which may form the basis of manuscript(s) intended for publication. The Clinical Study Report or summary thereof will be provided to the local IRB of the institutes participating in the protocol. Additionally the same will be made available on the trial registration websites.

#### References


[1] National Cancer Registry Programme Investigator Group, Mathur P, Sathishkumar K, Das P, Santhappan S, Sankarapillai J, et al. Cancer incidence and mortality across 43 cancer registries in India. JAMA Netw Open 2025;8:e2527805. https://doi.org/10.1001/jamanetworkopen.2025.27805.

[2] Pötter R, Tanderup K, Schmid MP, Jürgenliemk-Schulz I, Haie-Meder C, Fokdal LU, et al. MRI-guided adaptive brachytherapy in locally advanced cervical cancer (EMBRACE-I): a multicentre prospective cohort study. Lancet Oncol 2021;22:538–47. https://doi.org/10.1016/S1470-2045(20)30753-1.

[3] De Leeuw AAC, Nout RA, Van Leeuwen RGH, Mans A, Verhoef LG, Jürgenliemk-Schulz

IM. Implementation of state-of-the-art (chemo)radiation for advanced cervix cancer in the Netherlands: A quality improvement program. Technical Innovations & Patient Support in Radiation Oncology 2019;9:1–7. https://doi.org/10.1016/j.tipsro.2018.10.001.
[4] Lindegaard JC, Kirisits C, Schmid MP, Wulff CN, Steen SG, Kristoffersen KB, et al. Impact of Patient Selection on Real-World Outcomes by Using the EMBRACE-II Treatment Protocol in Locally Advanced Cervical Cancer. Int J Radiat Oncol Biol Phys 2025;123:669–80. https://doi.org/10.1016/j.ijrobp.2025.05.068.
[5] McCormack M, Eminowicz G, Gallardo D, Diez P, Farrelly L, Kent C, et al. Induction chemotherapy followed by standard chemoradiotherapy versus standard chemoradiotherapy alone in patients with locally advanced cervical cancer (GCIG INTERLACE): an international, multicentre, randomised phase 3 trial. Lancet 2024. https://doi.org/10.1016/s0140-6736(24)01438-7.
[6] Achari RB, Chakraborty S, Ray S, Mahata A, Mandal S, Das J, et al. 18F-fluorodeoxyglucose PET-CT-guided pelvic chemoradiation therapy using helical tomotherapy for locally advanced carcinoma cervix without para-aortic nodal disease: Clinical and patient-reported outcomes from a prospective phase 2 study. J Med Imaging Radiat Oncol 2024;68:624–34. https://doi.org/10.1111/1754-9485.13667.
[7] Kobayashi R, Yamashita H, Okuma K, Ohtomo K, Nakagawa K. Details of recurrence sites after definitive radiation therapy for cervical cancer. J Gynecol Oncol 2016;27:e16. https://doi.org/10.3802/jgo.2016.27.e16.
[8] Nomden CN, Pötter R, de Leeuw AAC, Tanderup K, Lindegaard JC, Schmid MP, et al. Nodal failure after chemo-radiation and MRI guided brachytherapy in cervical cancer: Patterns of failure in the EMBRACE study cohort. Radiother Oncol 2019;134:185–90. https://doi.org/10.1016/j.radonc.2019.02.007.
[9] Peters M, de Leeuw AAC, Nomden CN, Tanderup K, Kirchheiner K, Lindegaard JC, et al. Risk factors for nodal failure after radiochemotherapy and image guided brachytherapy in locally advanced cervical cancer: An EMBRACE analysis. Radiother Oncol 2021;163:150–8. https://doi.org/10.1016/j.radonc.2021.08.020.
[10] Beadle BM, Jhingran A, Yom SS, Ramirez PT, Eifel PJ. Patterns of regional recurrence after definitive radiotherapy for cervical cancer. Int J Radiat Oncol Biol Phys 2010;76:1396–403. https://doi.org/10.1016/j.ijrobp.2009.04.009.
[11] van den Bosch S, Doornaert PAH, Hoebers FJP, Kreike B, Vergeer MR, Zwijnenburg EM, et al. Clinical benefit and safety of reduced elective dose in definitive radiotherapy for head and neck squamous cell carcinoma: The UPGRADE-RT multicenter randomized controlled trial. J Clin Oncol 2025;43:2583–94. https://doi.org/10.1200/JCO-24-02194.
[12] Nuyts S, Lambrecht M, Duprez F, Daisne J-F, Van Gestel D, Van den Weyngaert D, et al. Reduction of the dose to the elective neck in head and neck squamous cell carcinoma, a randomized clinical trial using intensity modulated radiotherapy (IMRT). Dosimetrical analysis and effect on acute toxicity. Radiother Oncol 2013;109:323–9. https://doi.org/10.1016/j.radonc.2013.06.044.
[13] Nevens D, Duprez F, Daisne J-F, Schatteman J, Van der Vorst A, De Neve W, et al. Recurrence patterns after a decreased dose of 40Gy to the elective treated neck in head and neck cancer. Radiother Oncol 2017;123:419–23. https://doi.org/10.1016/j.radonc.2017.03.003.
[14] Sher DJ, Moon DH, Vo D, Wang J, Chen L, Dohopolski M, et al. Efficacy and quality-of-life following involved nodal radiotherapy for head and neck squamous cell carcinoma: The INRT-AIR phase II clinical trial. Clin Cancer Res 2023;29:3284–91. https://doi.org/10.1158/1078-0432.CCR-23-0334.

[15] Tsai CJ, McBride SM, Riaz N, Kang JJ, Spielsinger DJ, Waldenberg T, et al. Evaluation of substantial reduction in elective radiotherapy dose and field in patients with human Papillomavirus-associated oropharyngeal carcinoma treated with definitive chemoradiotherapy. JAMA Oncol 2022;8:364–72. https://doi.org/10.1001/jamaoncol.2021.6416.
[16] Feng M, Hallemeier CL, Almada C, Aranha O, Dorth J, Felder S, et al. Radiation therapy for anal squamous cell carcinoma: An ASTRO clinical practice guideline. Pract Radiat Oncol 2025;15:367–86. https://doi.org/10.1016/j.prro.2025.02.001.
[17] Lépinoy A, Lescut N, Puyraveau M, Caubet M, Boustani J, Lakkis Z, et al. Evaluation of a 36 Gy elective node irradiation dose in anal cancer. Radiother Oncol 2015;116:197–201. https://doi.org/10.1016/j.radonc.2015.07.050.
[18] Gilbert A, Adams R, Webster J, Gilbert DC, Abbott NL, Berkman L, et al. Standard versus reduced-dose chemoradiotherapy in anal cancer (PLATO-ACT4): short-term results of a phase 2 randomised controlled trial. Lancet Oncol 2025;26:707–18. https://doi.org/10.1016/S1470-2045(25)00213-X.
[19] Muñoz N, Bosch FX, de Sanjosé S, Herrero R, Castellsagué X, Shah KV, et al. Epidemiologic classification of human papillomavirus types associated with cervical cancer. N Engl J Med 2003;348:518–27. https://doi.org/10.1056/NEJMoa021641.
[20] IARC Working Group on the Evaluation of Carcinogenic Risks to Humans. Molecular Mechanisms of HPV-induced Carcinogenesis. Human Papillomaviruses, International Agency for Research on Cancer; 2007.
[21] Pirog EC. Cervical adenocarcinoma: Diagnosis of human Papillomavirus-positive and human Papillomavirus-negative tumors. Arch Pathol Lab Med 2017;141:1653–67. https://doi.org/10.5858/arpa.2016-0356-RA.
[22] Xing B, Guo J, Sheng Y, Wu G, Zhao Y. Human Papillomavirus-negative cervical cancer: A comprehensive review. Front Oncol 2020;10:606335. https://doi.org/10.3389/fonc.2020.606335.
[23] Nicolás I, Marimon L, Barnadas E, Saco A, Rodríguez-Carunchio L, Fusté P, et al. HPV-negative tumors of the uterine cervix. Mod Pathol 2019;32:1189–96. https://doi.org/10.1038/s41379-019-0249-1.
[24] van der Marel J, van Baars R, Quint WGV, Berkhof J, del Pino M, Torné A, et al. The impact of human papillomavirus genotype on colposcopic appearance: a cross-sectional analysis. BJOG 2014;121:1117–26. https://doi.org/10.1111/1471-0528.12668.
[25] Rodríguez-Carunchio L, Soveral I, Steenbergen RDM, Torné A, Martinez S, Fusté P, et al. HPV-negative carcinoma of the uterine cervix: a distinct type of cervical cancer with poor prognosis. BJOG 2015;122:119–27. https://doi.org/10.1111/1471-0528.13071.
[26] Liana S. Mkrtchian, Irina A. Zamulaeva, Liudmila I. Krikunova, Kiseleva Vi, Matchuk On, Liubov Liubina, et al. HPV Status and Individual Characteristics of Human Papillomavirus Infection as Predictors for Clinical Outcome of Locally Advanced Cervical Cancer. Journal of Personalized Medicine 2021. https://doi.org/10.3390/jpm11060479.
[27] Huang Y, Zou D, Guo M, He M, He H, Li X, et al. HPV and radiosensitivity of cervical cancer: a narrative review. Ann Transl Med 2022;10:1405. https://doi.org/10.21037/atm-22-5930.
[28] Kimple RJ, Smith MA, Blitzer GC, Torres AD, Martin JA, Yang RZ, et al. Enhanced radiation sensitivity in HPV-positive head and neck cancer. Cancer Res 2013;73:4791–800. https://doi.org/10.1158/0008-5472.CAN-13-0587.
[29] Rietbergen MM, Martens-de Kemp SR, Bloemena E, Witte BI, Brink A, Baatenburg de Jong RJ, et al. Cancer stem cell enrichment marker CD98: a prognostic factor for survival in

patients with human papillomavirus-positive oropharyngeal cancer. Eur J Cancer 2014;50:765–73. https://doi.org/10.1016/j.ejca.2013.11.010.
[30] Liu C, Mann D, Sinha UK, Kokot NC. The molecular mechanisms of increased radiosensitivity of HPV-positive oropharyngeal squamous cell carcinoma (OPSCC): an extensive review. J Otolaryngol Head Neck Surg 2018;47:59. https://doi.org/10.1186/s40463-018-0302-y.
[31] Martinelli C, Ercoli A, Parisi S, Iatì G, Pergolizzi S, Alfano L, et al. Molecular mechanisms and clinical divergences in HPV-positive cervical vs. Oropharyngeal cancers: A critical narrative review. BMC Med 2025;23:405. https://doi.org/10.1186/s12916-025-04247-z.
[32] Rai B, Dey T, Ballari N, Singh M, Miryala R, Srinivasa GY, et al. Three-dimensional conformal radiotherapy versus image-guided intensity modulated external beam radiotherapy in locally advanced cervical cancer: A phase III randomized control study. Clin Oncol (R Coll Radiol) 2024;36:728–37. https://doi.org/10.1016/j.clon.2024.08.004.
[33] Kapoor AR, Bhalavat RL, Chandra M, Pareek V, Moosa Z, Markana S, et al. A randomized study for dosimetric assessment and clinical impact of bone marrow sparing intensity-modulated radiation therapy versus 3-dimensional conformal radiation therapy on hematological and gastrointestinal toxicities in cervical cancer. J Cancer Res Ther 2022;18:1490–7. https://doi.org/10.4103/jcrt.jcrt_1242_20.
[34] Naik A, Gurjar OP, Gupta KL, Singh K, Nag P, Bhandari V. Comparison of dosimetric parameters and acute toxicity of intensity-modulated and three-dimensional radiotherapy in patients with cervix carcinoma: A randomized prospective study. Cancer Radiother 2016;20:370–6. https://doi.org/10.1016/j.canrad.2016.05.011.
[35] Gandhi AK, Sharma DN, Rath GK, Julka PK, Subramani V, Sharma S, et al. Early clinical outcomes and toxicity of intensity modulated versus conventional pelvic radiation therapy for locally advanced cervix carcinoma: A prospective randomized study. Int J Radiat Oncol Biol Phys 2013;87:542–8. https://doi.org/10.1016/j.ijrobp.2013.06.2059.
[36] Corbeau A, Kuipers SC, de Boer SM, Horeweg N, Hoogeman MS, Godart J, et al. Correlations between bone marrow radiation dose and hematologic toxicity in locally advanced cervical cancer patients receiving chemoradiation with cisplatin: a systematic review. Radiother Oncol 2021;164:128–37. https://doi.org/10.1016/j.radonc.2021.09.009.
[37] Li W, Ma L, Li F, Li K, Zhang Y, Ren H, et al. Effects of bone marrow sparing radiotherapy on acute hematologic toxicity for patients with locoregionally advanced cervical cancer: a prospective phase II randomized controlled study. Radiat Oncol 2024;19:46. https://doi.org/10.1186/s13014-024-02432-7.
[38] Huang J, Gu F, Ji T, Zhao J, Li G. Pelvic bone marrow sparing intensity modulated radiotherapy reduces the incidence of the hematologic toxicity of patients with cervical cancer receiving concurrent chemoradiotherapy: a single-center prospective randomized controlled trial. Radiat Oncol 2020;15:180. https://doi.org/10.1186/s13014-020-01606-3.
[39] Zhou P, Zhang Y, Luo S, Zhang S. Pelvic bone marrow sparing radiotherapy for cervical cancer: A systematic review and meta-analysis. Radiother Oncol 2021;165:103–18. https://doi.org/10.1016/j.radonc.2021.10.015.
[40] Williamson CW, Sirák I, Xu R, Portelance L, Wei L, Tarnawski R, et al. Positron Emission Tomography-Guided Bone Marrow-Sparing Radiation Therapy for Locoregionally Advanced Cervix Cancer: Final Results From the INTERTECC Phase II/III Trial. Int J Radiat Oncol Biol Phys 2021. https://doi.org/10.1016/j.ijrobp.2021.08.019.
[41] Thayer-Freeman C, Washington B, Fabian D, Cheek D, Clair WS, Bernard M, et al. In vitro α/β ratio variations in cervical cancer, with consequent effects on equivalent dose in 2 Gy fraction in high-dose-rate brachytherapy. Adv Radiat Oncol 2025;10:101725.

https://doi.org/10.1016/j.adro.2025.101725.
[42] Maddah Safaei A, Esmati E, Gomar M, Akhavan S, Sheikh Hasani S, Malekzadeh Moghani M, et al. Hypofractionated versus standard chemoradiotherapy in the definitive treatment of uterine cervix cancer: interim results of a randomized controlled clinical trial. J Cancer Res Clin Oncol 2024;150:20. https://doi.org/10.1007/s00432-023-05563-8.
[43] Dankulchai P, Prasartseree T, Sittiwong W, Thephamongkhol K, Nakkrasae P. Early results of hypofractionated chemoradiation in cervical cancer with 44 Gy/ 20 F vs 45 Gy/ 25 F: A phase II, open-label, randomised controlled trial (HYPOCx-iRex trial). Clin Oncol (R Coll Radiol) 2025;46:103907. https://doi.org/10.1016/j.clon.2025.103907.
[44] Herring DF. The degree of precision required in the radiation dose delivered in cancer radiotherapy. Brit J Radiol 1971:51–8.
[45] Gundog M, Basaran H, Bozkurt O, Eroglu C. A comparison of cisplatin cumulative dose and cisplatin schedule in patients treated with concurrent chemo-radiotherapy in nasopharyngeal carcinoma. Braz J Otorhinolaryngol 2020;86:676–86. https://doi.org/10.1016/j.bjorl.2019.04.008.
[46] Helfenstein S, Riesterer O, Meier UR, Papachristofilou A, Kasenda B, Pless M, et al. 3-weekly or weekly cisplatin concurrently with radiotherapy for patients with squamous cell carcinoma of the head and neck - a multicentre, retrospective analysis. Radiat Oncol 2019;14:32. https://doi.org/10.1186/s13014-019-1235-y.
[47] Prathipati A, Jilla S, Subramanian BV, Madala RK. Impact of Various Prognostic Factors on Overall Survival, Disease-Free Survival and Patterns of Failure in Carcinoma Cervix: A Tertiary Care Centre Experience from South India. Indian Journal of Gynecologic Oncology 2018;16:11. https://doi.org/10.1007/s40944-018-0179-8.
[48] Chen M-F, Tseng C-J, Tseng C-C, Kuo Y-C, Yu C-Y, Chen W-C. Clinical Outcome in Posthysterectomy Cervical Cancer Patients Treated With Concurrent Cisplatin and Intensity-Modulated Pelvic Radiotherapy: Comparison With Conventional Radiotherapy. International Journal of Radiation Oncology*Biology*Physics 2007;67:1438–44. https://doi.org/10.1016/j.ijrobp.2006.11.005.
[49] Akyildiz A, Gultekin M, Yigit E, Demir E, Ismayilov R, Ahmed M, et al. Efficacy of cumulative cisplatin dose on survival in patients with locally advanced cervical cancer treated with definitive chemoradiotherapy: multicenter study by Turkish Oncology Group. Int J Gynecol Cancer 2024;34:1359–65. https://doi.org/10.1136/ijgc-2024-005419.
[50] Moos K, Baldinger M, Perez Haas Y, Ludwig R, Looman E, Balermpas P, et al. A tumor control probability model for elective nodal irradiation to balance toxicity and regional tumor control in treatment plan optimization for head-and-neck squamous cell carcinoma. Phys Med Biol 2026;71:035022. https://doi.org/10.1088/1361-6560/ae4165.
[51] Okunieff P, Morgan D, Niemierko A, Suit HD. Radiation dose-response of human tumors. International Journal of Radiation Oncology, Biology, Physics 1995;32:1227–37. https://doi.org/10.1016/0360-3016(94)00475-Z.
[52] Lakomy DS, Wu J, Lombe D, Papasavvas E, Msadabwe SC, Geng Y, et al. Immune correlates of therapy outcomes in women with cervical cancer treated with chemoradiotherapy: A systematic review. Cancer Med 2021;10:4206–20. https://doi.org/10.1002/cam4.4017.
[53] Li Y, Liu A, Wang X, Guo L, Li Y, Liu D, et al. The role of lymphocyte recovery index in prognosis prediction for locally advanced cervical cancer with radiation-induced lymphopenia. Cancer Med 2025;14:e70638. https://doi.org/10.1002/cam4.70638.
[54] Pham T-N, Coupey J, Thariat J, Valable S. Impact of circulating lymphocyte kinetics following radiotherapy on patient survival: A model-based meta-analysis. Comput Biol Med

2025;186:109702. https://doi.org/10.1016/j.compbiomed.2025.109702.
[55] Yang A, Janowski E, Siebers JV, Karki A, Jin R, Romano K. Impact of post-treatment lymphopenia on clinical outcomes in locally advanced cervical cancer. Int J Radiat Oncol Biol Phys 2025;123:e400. https://doi.org/10.1016/j.ijrobp.2025.06.2408.
[56] Damen PJJ, Kroese TE, van Hillegersberg R, Schuit E, Peters M, Verhoeff JJC, et al. The influence of severe radiation-induced lymphopenia on overall survival in solid tumors: A systematic review and meta-analysis. Int J Radiat Oncol Biol Phys 2021;111:936–48. https://doi.org/10.1016/j.ijrobp.2021.07.1695.
[57] Cho O, Chun M, Chang S-J, Oh Y-T, Noh OK. Prognostic value of severe lymphopenia during pelvic concurrent chemoradiotherapy in cervical cancer. Anticancer Res 2016;36:3541–7.
[58] Yang L, Xu Z, Ma L, Liu Q, Chang ATY, Wang Q, et al. Early onset of severe lymphopenia during definitive radiotherapy correlates with mean body dose and predicts poor survival in cervical cancer. Cancer Biomark 2022;34:149–59. https://doi.org/10.3233/CBM-210292.
[59] Anderson K. gsDesign: Group Sequential Design. 2026.
[60] Harris PA, Taylor R, Thielke R, Payne J, Gonzalez N, Conde JG. Research electronic data capture (REDCap)--a metadata-driven methodology and workflow process for providing translational research informatics support. J Biomed Inform 2009;42:377–81. https://doi.org/10.1016/j.jbi.2008.08.010.
[61] Harris PA, Taylor R, Minor BL, Elliott V, Fernandez M, O'Neal L, et al. The REDCap consortium: Building an international community of software platform partners. J Biomed Inform 2019;95:103208. https://doi.org/10.1016/j.jbi.2019.103208.
[62] Zou G. A modified poisson regression approach to prospective studies with binary data. Am J Epidemiol 2004;159:702–6. https://doi.org/10.1093/aje/kwh090.
[63] Rohde MD, French B, Stewart TG, Harrell FE Jr. Bayesian transition models for ordinal longitudinal outcomes. Stat Med 2024. https://doi.org/10.1002/sim.10133.
[64] Giesinger JM, Kieffer JM, Fayers PM, Groenvold M, Petersen MA, Scott NW, et al. Replication and validation of higher order models demonstrated that a summary score for the EORTC QLQ-C30 is robust. J Clin Epidemiol 2016;69:79–88. https://doi.org/10.1016/j.jclinepi.2015.08.007.

### Appendix I: Biological Materials

#### Biobanking Methodology

Informed consent will be taken from the patient for biospecimen preservation. The following will be the schedule of biospecimen acquisition:

| Timing | Plasma | Tissue |
|---|---|---|
| Baseline | X | X |
| End of Chemoradiation / Pre Brachytherapy | X | X |
| 3 months | X | |
| 6 months | X | |
| Recurrence* | X | X |

* Specimen to be obtained if clinically feasible.

##### *Whole Blood and Plasma Banking*

7 - 10 mL of whole whole blood will be obtained after venipuncture at the schedules indicated in EDTA vacutainers. 2 - 3 ml of blood will be separated into 1 ml aliquots. Remaining blood centrifuged at 200g at 4 degree C for 10 min. 500 µl of plasma layer will be aliquoted using 1 ml pipette into 500 µl cryo-vials. Store the 1 ml aliquots and 500µl plasma aliquots in a cryo-storage box at -80°C. The samples will be labelled with a barcoded sticker with BV ID (Biobank ID) and sample ID generated by Labvantage.

##### *Tissue Banking*

Tissue specimen processing will be dependent on the time of acquisition. Baseline specimens and specimens obtained at the time of recurrence will be formalin fixed and embedded in paraffin as they will be obtained at the time of biopsy. Additionally if possible, biopsy specimens will also be preserved in RNALater. Typically a biopsy specimen size of 0.5 cm or less will be placed in 5 - 10 volumes of RNA later (500 - 700 µl). Tissues in RNALater will be stored in 4 degree Celsius. After that the RNALater supernatant will be removed and the tissue stored in cryovials at -80°C. The cryovials will be labelled with a barcoded sticker with the BV ID (Biobank ID) and sample ID generated by Labvantage.

### Appendix II: Definition of Acute Toxicity related endpoints

| Toxicity | Definition | Grade 1 | Grade 2 | Grade 3 | Grade 4 |
|---|---|---|---|---|---|
| Diarrhea | A disorder characterized by an increase in frequency and/or loose or watery bowel movements. | Change in consistency or frequency | Increase of 4 - 6 stools per day over baseline; moderate increase in ostomy output compared to baseline; limiting instrumental ADL or mild/moderate impact on age-appropriate normal daily activity (pediatric); change in consistency or frequency AND limiting instrumental ADL or mild/moderate impact on age-appropriate normal daily activity (pediatric) | Increase of >=7 stools per day over baseline; hospitalization indicated; severe increase in ostomy output compared to baseline; requires IV intervention; limiting self-care ADL or severe impact on age-appropriate normal daily activity (pediatric) | Life-threatening consequences; urgent intervention indicated |
| Constipation | A disorder characterized by irregular and infrequent or difficult evacuation of the bowels. | Occasional or intermittent symptoms; occasional use of stool softeners, laxatives, dietary modification, or enema | Persistent symptoms with regular use of laxatives or enemas; limiting instrumental ADL or mild/moderate impact on age-appropriate normal daily activity (pediatric) | Obstipation with manual evacuation indicated; limiting self-care ADL or severe impact on age-appropriate normal daily activity (pediatric) | Life-threatening consequences; urgent intervention indicated |
| Nausea | A disorder characterized by a queasy sensation and/or the urge to vomit. | Loss of appetite without alteration in eating habits | Oral intake decreased without significant weight loss, dehydration or malnutrition; IV intervention indicated | Inadequate oral caloric or fluid intake; tube feeding, TPN, or hospitalization indicated | NA |

| Toxicity | Definition | Grade 1 | Grade 2 | Grade 3 | Grade 4 |
|---|---|---|---|---|---|
| Vomiting | A disorder characterized by the reflexive act of ejecting the contents of the stomach through the mouth. | Intervention not indicated | Initiation of outpatient IV hydration; medical intervention indicated | Initiation of tube feeding, or TPN; hospitalization indicated | Life-threatening consequences |
| Proctitis | A disorder characterized by inflammation of the rectum. | Rectal discomfort, intervention not indicated | Symptomatic (e.g., rectal discomfort, passing blood or mucus); fecal urgency or stool incontinence; medical intervention indicated; limiting instrumental ADL | Severe symptoms; limiting self-care ADL | Life-threatening consequences; urgent intervention indicated |
| Abdominal Pain | A disorder characterized by a sensation of marked discomfort in the abdominal region. | Mild pain | Moderate pain; limiting instrumental ADL | Severe pain; limiting self-care ADL | NA |
| Dyspepsia | A disorder characterized by an uncomfortable, often painful feeling in the stomach, resulting from impaired digestion. Symptoms include burning stomach, bloating, heartburn, nausea and vomiting. | Mild symptoms; intervention not indicated | Moderate symptoms; medical intervention indicated | Severe symptoms; operative intervention indicated | NA |
| Malabsorption | A disorder characterized by inadequate absorption of | NA | Altered diet; oral intervention indicated | Inability to aliment adequately; TPN indicated | Life-threatening consequences; urgent intervention |

| Toxicity | Definition | Grade 1 | Grade 2 | Grade 3 | Grade 4 |
|---|---|---|---|---|---|
| | nutrients in the small intestine. Symptoms include abdominal marked discomfort, bloating and diarrhea. | | | | indicated |
| Small Intestinal Obstruction | A disorder characterized by blockage of the normal flow of the intestinal contents of the small intestine. | Asymptomatic; clinical or diagnostic observations only; intervention not indicated | Symptomatic; altered GI function; limiting instrumental ADL or mild/moderate impact on age-appropriate normal daily activity (pediatric) | Hospitalization indicated; invasive intervention indicated; limiting self care ADL or severe impact on age-appropriate normal daily activity (pediatric) | Life-threatening consequences; urgent operative intervention indicated |
| Small Intestinal Perforation | A disorder characterized by a rupture in the small intestine wall. | NA | Invasive intervention not indicated | Invasive intervention indicated | Life-threatening consequences; urgent operative intervention indicated |
| Noninfective Cystitis | A disorder characterized by inflammation of the bladder which is not caused by an infection of the urinary tract. | Microscopic hematuria; minimal increase in frequency, urgency, dysuria, or nocturia; new onset of incontinence | Moderate hematuria; moderate increase in frequency, urgency, dysuria, nocturia or incontinence; urinary catheter placement or bladder irrigation indicated; limiting instrumental ADL or mild/moderate impact on age-appropriate normal daily activity (pediatric) | Gross hematuria; transfusion, IV medications, or hospitalization indicated; elective invasive intervention indicated | Life-threatening consequences; urgent intervention indicated |
| Bladder Perforation | A disorder characterized by | NA | Invasive intervention not | Invasive intervention | Life-threatening consequences; |

| Toxicity | Definition | Grade 1 | Grade 2 | Grade 3 | Grade 4 |
|---|---|---|---|---|---|
| | a rupture in the bladder wall. | | indicated | indicated | organ failure; urgent operative intervention indicated |
| Dysuria | A disorder characterized by painful urination. | Present | NA | NA | NA |
| Urinary Fistula | A disorder characterized by an abnormal communication between any part of the urinary system and another organ or anatomic site. | NA | Symptomatic, invasive intervention not indicated | Invasive intervention indicated | Life-threatening consequences; urgent invasive intervention indicated |
| Urinary Frequency | A disorder characterized by urination at short intervals. | Present | NA | NA | NA |
| Urinary Incontinence | A disorder characterized by inability to control the flow of urine from the bladder. | Occasional (e.g., with coughing, sneezing, etc.), pads not indicated | Spontaneous; pads indicated; limiting instrumental ADL or mild/moderate impact on age-appropriate normal daily activity (pediatric) | Intervention indicated (e.g., clamp, collagen injections); operative intervention indicated; limiting self-care ADL or severe impact on age-appropriate normal daily activity (pediatric) | NA |
| Urinary Retention | A disorder characterized by accumulation of urine within the bladder because of the inability to urinate. | Urinary, suprapubic or intermittent catheter placement not indicated; able to void with some residual | Placement of urinary, suprapubic or intermittent catheter placement indicated; medication indicated | Elective invasive intervention indicated; substantial loss of affected kidney function or mass | Life-threatening consequences; organ failure; urgent operative intervention indicated |
| Urinary | A disorder | Present | Medical | NA | NA |

| Toxicity | Definition | Grade 1 | Grade 2 | Grade 3 | Grade 4 |
|---|---|---|---|---|---|
| Urgency | characterized by a sudden compelling urge to urinate. | | management indicated; limiting instrumental ADL or mild/moderate impact on age-appropriate normal daily activity (pediatric) | | |
| Anemia | A disorder characterized by a reduction in the amount of hemoglobin in 100 ml of blood. Signs and symptoms of anemia may include pallor of the skin and mucous membranes, shortness of breath, palpitations of the heart, soft systolic murmurs, lethargy, and fatigability. | Hemoglobin (Hgb) <LLN - 10.0 g/dL; <LLN - 6.2 mmol/L; <LLN - 100 g/L | Hgb <10.0 - 8.0 g/dL; <6.2 - 4.9 mmol/L; <100 - 80 g/L | Hgb <8.0 g/dL; <4.9 mmol/L; <80 g/L; transfusion indicated | Life-threatening consequences; urgent intervention indicated |
| Thrombocytopenia | A disorder characterized by a decrease in the number of platelets in a blood specimen. | <LLN - 75,000/mm3; <LLN - 75.0 x 10^9/L | <75,000 - 50,000/mm3; <75.0 - 50.0 x 10^9/L | <50,000 - 10,000/mm3; <50.0 - 10.0 x 10^9/L; transfusion indicated | <10,000/mm3; <10.0 x 10^9/L; life-threatening consequences; urgent intervention indicated |
| Febrile Neutropenia | A disorder characterized by an ANC <1000/mm3 and a single temperature of >38.3 degrees C (101 degrees F) or a sustained temperature of | NA | NA | ANC <1000/mm3 with a single temperature of >38.3 degrees C (101 degrees F) or a sustained temperature of >=38 degrees C (100.4 degrees F) for more than one | Life-threatening consequences; urgent intervention indicated |

| Toxicity | Definition | Grade 1 | Grade 2 | Grade 3 | Grade 4 |
|---|---|---|---|---|---|
| | >=38 degrees C (100.4 degrees F) for more than one hour. | | | hour | |
| Neutropenia | A finding based on laboratory test results that indicate a decrease in number of neutrophils (ANC) in a blood specimen. | <1500 - 1000/mm3; <1.5 - 1.0 x 10^9/L | <1000 - 500/mm3; <1.0 - 0.5 x 10^9/L | <500 - 100/mm3; <0.5 - 0.1 x 10^9/L | <100/mm3; <0.1 x 10^9/L |
| Hyponatremia | A disorder characterized by laboratory test results that indicate a low concentration of sodium in the blood. | <LLN - 130 mmol/L | 125 - <130 mmol/L and asymptomatic | 125 - <130 mmol/L symptomatic; 120 - <125 mmol/L regardless of symptoms | <120 mmol/L; life-threatening consequences |
| Hypokalemia | A disorder characterized by laboratory test results that indicate a low concentration of potassium in the blood. | <LLN - 3.0 mmol/L | Symptomatic with <LLN - 3.0 mmol/L; intervention indicated | <3.0 - 2.5 mmol/L; hospitalization indicated | <2.5 mmol/L; life-threatening consequences |
| Hypocalcemnia | A disorder characterized by laboratory test results that indicate a low concentration of calcium (corrected for albumin) in the blood. | Corrected serum calcium of <LLN - 8.0 mg/dL; <LLN - 2.0 mmol/L; Ionized calcium <LLN - 1.0 mmol/L | Corrected serum calcium of <8.0 - 7.0 mg/dL; <2.0 - 1.75 mmol/L; Ionized calcium <1.0 - 0.9 mmol/L; symptomatic | Corrected serum calcium of <7.0 - 6.0 mg/dL; <1.75 - 1.5 mmol/L; Ionized calcium <0.9 - 0.8 mmol/L; hospitalization indicated | Corrected serum calcium of <6.0 mg/dL; <1.5 mmol/L; Ionized calcium <0.8 mmol/L; life-threatening consequences |
| Hypomagnesemia | A disorder characterized by laboratory test results that indicate a low concentration of magnesium in | <LLN - 1.2 mg/dL; <LLN - 0.5 mmol/L | <1.2 - 0.9 mg/dL; <0.5 - 0.4 mmol/L | <0.9 - 0.7 mg/dL; <0.4 - 0.3 mmol/L | <0.7 mg/dL; <0.3 mmol/L; life-threatening consequences |

| Toxicity | Definition | Grade 1 | Grade 2 | Grade 3 | Grade 4 |
|---|---|---|---|---|---|
| | the blood. | | | | |
| Hot Flashes | A disorder characterized by an uncomfortable and temporary sensation of intense body warmth, flushing, sometimes accompanied by sweating upon cooling. | Mild symptoms; intervention not indicated | Moderate symptoms; limiting instrumental ADL or mild/moderate impact on age-appropriate normal daily activity (pediatric) | Severe symptoms; limiting self-care ADL or severe impact on age-appropriate normal daily activity (pediatric) | NA |
| Vaginal Discharge | A disorder characterized by vaginal secretions. Mucus produced by the cervical glands is discharged from the vagina naturally, especially during the childbearing years. | Mild vaginal discharge (greater than baseline for patient) | Moderate to heavy vaginal discharge; use of perineal pad or tampon indicated | NA | NA |
| Vaginal Dryness | A disorder characterized by an uncomfortable feeling of itching and burning in the vagina. | Mild vaginal dryness not interfering with sexual function | Moderate vaginal dryness interfering with sexual function or causing frequent discomfort | Severe vaginal dryness resulting in dyspareunia or severe discomfort | NA |
| Vaginal Fistula | A disorder characterized by an abnormal communication between the vagina and another organ or anatomic site. | Asymptomatic | Symptomatic, invasive intervention not indicated | Invasive intervention indicated | Life-threatening consequences; urgent intervention indicated |
| Vaginal Hemorrhage | A disorder characterized by bleeding from the vagina. | Mild symptoms; intervention not indicated | Moderate symptoms; intervention indicated | Hospitalization | Life-threatening consequences; urgent intervention |

| Toxicity | Definition | Grade 1 | Grade 2 | Grade 3 | Grade 4 |
|---|---|---|---|---|---|
| | | | | | indicated |
| Vaginal Stricture | A disorder characterized by a narrowing of the vaginal canal. | Asymptomatic; mild vaginal shortening or narrowing | Vaginal narrowing and/or shortening not interfering with physical examination | Vaginal narrowing and/or shortening interfering with the use of tampons, sexual activity or physical examination | NA |
| Anal Fissure | A disorder characterized by a tear in the lining of the anus. | Asymptomatic | Symptomatic | Invasive intervention indicated | NA |
| Anal Fistula | A disorder characterized by an abnormal communication between the opening in the anal canal to the perianal skin. | Asymptomatic | Symptomatic, invasive intervention not indicated | Invasive intervention indicated | Life-threatening consequences; urgent intervention indicated |
| Anal Stenosis | A disorder characterized by a narrowing of the lumen of the anal canal. | Asymptomatic; clinical or diagnostic observations only; intervention not indicated | Symptomatic; altered GI function | Symptomatic and severely altered GI function; non-emergent operative intervention indicated; TPN or hospitalization indicated | Life-threatening consequences; urgent operative intervention indicated |
| Hemorrhoids | A disorder characterized by the presence of dilated veins in the rectum and surrounding area. | Asymptomatic; clinical or diagnostic observations only; intervention not indicated | Symptomatic; banding or medical intervention indicated | Severe symptoms; invasive intervention indicated | NA |
| Fecal Incontinence | A disorder characterized by inability to control the escape of stool from the rectum. | Occasional use of pads required | Daily use of pads required | Severe symptoms; elective operative intervention indicated | NA |

| Toxicity | Definition | Grade 1 | Grade 2 | Grade 3 | Grade 4 |
|---|---|---|---|---|---|
| Rectal Fistula | A disorder characterized by an abnormal communication between the rectum and another organ or anatomic site. | Asymptomatic | Symptomatic, invasive intervention not indicated | Invasive intervention indicated | Life-threatening consequences; urgent intervention indicated |
| Rectal Necrosis | A disorder characterized by a necrotic process occurring in the rectal wall. | NA | NA | Tube feeding or TPN indicated; invasive intervention indicated | Life-threatening consequences; urgent operative intervention indicated |
| Rectal Obstruction | A disorder characterized by blockage of the normal flow of the intestinal contents in the rectum. | Asymptomatic; clinical or diagnostic observations only; intervention not indicated | Symptomatic; altered GI function; limiting instrumental ADL or mild/moderate impact on age-appropriate normal daily activity (pediatric) | Hospitalization indicated; invasive intervention indicated; limiting self care ADL or severe impact on age-appropriate normal daily activity (pediatric) | Life-threatening consequences; urgent operative intervention indicated |
| Rectal Perforation | A disorder characterized by a rupture in the rectal wall. | NA | Invasive intervention not indicated | Invasive intervention indicated | Life-threatening consequences; urgent operative intervention indicated |
| Rectal Stenosis | A disorder characterized by a narrowing of the lumen of the rectum. | Asymptomatic; clinical or diagnostic observations only; intervention not indicated | Symptomatic; altered GI function | Severely altered GI function; tube feeding or hospitalization indicated; elective operative intervention indicated | Life-threatening consequences; urgent operative intervention indicated |
| Fatigue | A disorder characterized by a state of generalized weakness with a pronounced inability to | Fatigue relieved by rest | Fatigue not relieved by rest; limiting instrumental ADL or mild/moderate impact on age-appropriate | Fatigue not relieved by rest, limiting self-care ADL or severe impact on age-appropriate normal daily | NA |

| Toxicity | Definition | Grade 1 | Grade 2 | Grade 3 | Grade 4 |
|---|---|---|---|---|---|
| | summon sufficient energy to accomplish daily activities. | | normal daily activity (pediatric) | activity (pediatric) | |
| Anorexia | A disorder characterized by a loss of appetite. | Loss of appetite without alteration in eating habits | Oral intake altered without significant weight loss or malnutrition; oral nutritional supplements indicated | Associated with significant weight loss or malnutrition (e.g., inadequate oral caloric and/or fluid intake); tube feeding or TPN indicated | Life-threatening consequences; urgent intervention indicated |
